# Serial Profiling of SARS-CoV-2 Antigens and Antibodies in COVID-19 Patient Plasma

**DOI:** 10.1101/2020.07.20.20156372

**Authors:** Alana F. Ogata, Adam M. Maley, Connie Wu, Tal Gilboa, Maia Norman, Roey Lazarovits, Chih-Ping Mao, Gail Newton, Matthew Chang, Katrina Nguyen, Maliwan Kamkaew, Quan Zhu, Travis E. Gibson, Edward T. Ryan, Richelle C. Charles, Wayne A. Marasco, David R. Walt

## Abstract

The severe acute respiratory syndrome coronavirus 2 (SARS-CoV-2) has infected millions of people worldwide. PCR tests are currently the gold standard for diagnosis of the current coronavirus disease (COVID-19) and serology tests are used to detect seroconversion in infected patients. However, there is a lack of quantitative and ultra-sensitive viral antigen tests for COVID-19. Here we show that Single Molecule Array (Simoa) assays can quantitatively detect SARS-CoV-2 spike, S1 subunit, and nucleocapsid antigens in the plasma of COVID-19 patients. Combined with Simoa anti-SARS-CoV-2 serological assays, we show correlation between production of antibodies and clearance of viral antigens from serial plasma samples from COVID-19 patients. Furthermore, we demonstrate the presence of viral antigens in blood correlates with disease severity in hospitalized COVID-19 patients. These data suggest that SARS-CoV-2 viral antigens in the blood could be a marker for severe COVID-19 cases.

**One Sentence Summary:** SARS-CoV-2 antigens S1, spike, and nucleocapsid and anti-SARS-Cov-2 antibodies were measured in longitudinal plasma samples from COVID-19 patients using Single Molecule Array (Simoa) assays.

## Introduction

The current pandemic of coronavirus disease (COVID-19), caused by the severe acute respiratory syndrome coronavirus 2 (SARS-CoV-2), has resulted in over 13,000,000 confirmed cases globally and over 135,000 deaths in the United States alone, as of July 16, 2020.(*1*) While RT-PCR tests remain the gold standard for diagnosing COVID-19, PCR tests do not provide adequate information on progression of the disease. Furthermore, PCR tests can give positive results for several weeks after a patient has seroconverted or recovered.(*2*) Serological tests can identify individuals who have mounted an immune response but cannot necessarily be used for monitoring disease in the early stages of infection.(*3*–*5*) SARS-CoV-2 antigen assays can be used for identifying active infection by measuring viral antigens in biofluids and can complement PCR and serological tests. Currently there are two FDA approved SARS-CoV-2 antigen tests that detect nucleocapsid in nasopharyngeal (NP) swabs.(*6, 7*) However, both tests provide qualitative results and may only detect viral antigen within the first five days of symptom onset. Quantitative and ultra-sensitive SARS-CoV-2 antigen assays could enable detection of viral antigens in blood, saliva, or NP swabs. In addition, combining quantitative antigen assays with serological assays could enable analysis of COVID-19 progression from early infection to seroconversion.

To address the need for a quantitative antigen assay, we developed ultra-sensitive Single Molecule Array (Simoa) SARS-CoV-2 antigen assays for S1, spike (S1-S2 extracellular domain), and nucleocapsid (N). The ultra-sensitivity of Simoa enables detection of SARS-CoV-2 antigens in the plasma of COVID-19 positive patients. Additionally, Simoa provides a dynamic range that allows quantification of antigens over a concentration range of four orders of magnitude. This precise quantification is advantageous for capturing the wide range of antigen concentrations in COVID-19 patient plasma throughout the course of hospitalization. We combined our Simoa SARS-CoV-2 antigen assays with previously developed Simoa serological assays to detect SARS-CoV-2 antigens and anti-SARS-CoV-2 immunoglobulins in longitudinal plasma samples of COVID-19 patients. These measurements show inverse correlation between anti-SARS-CoV-2 antibody production and viral antigen clearance from plasma, which provides a unique view of viral infection and immune response from the beginning of hospitalization through recovery or death. In addition, we show that the presence of viral antigen in plasma correlates with disease severity in these COVID-19 patients.

## Results & Discussion

The SARS-CoV-2 Simoa antigen assays detect S1, spike, and N antigens with limits of detection (LOD) of 5 pg/mL, 70 pg/mL, and 0.02 pg/mL, respectively. Three types of dye-encoded paramagnetic beads were functionalized with antibodies against each viral antigen and incubated with plasma samples for multiplexed Simoa measurements (Fig. 1a) as described previously.(*8*) Commercially available SARS-CoV-2 antibodies were used in combination with an anti-S1 antibody discovered via phage display (Supplemental Materials). All SARS-CoV-2 antigen assays were validated in commercial human plasma prior to analysis of clinical plasma samples (Fig. S1-S3, Table S3). In addition, we measured anti-SARS-CoV-2 immunoglobulins (total IgA, IgM, and IgG) using our recently established SARS-CoV-2 serological assays(*9*) to correlate viral antigens with immunoglobulin levels (Fig. 1a). Although total IgG levels correlate with seroconversion in COVID-19 patients, measurements of the IgG subclasses can reveal additional information on patient-specific immune responses. Therefore, we developed SARS-CoV-2 serological Simoa assays for IgG1, IgG2, IgG3, and IgG4 detection (Supplemental Materials Methods). The combined measurements of three SARS-CoV-2 antigens and seven anti-SARS-CoV-2 immunoglobulin isotypes against four SARS-CoV-2 antigens enables quantification of 31 biomarkers from 70 µL of a plasma sample.

**Figure 1.**
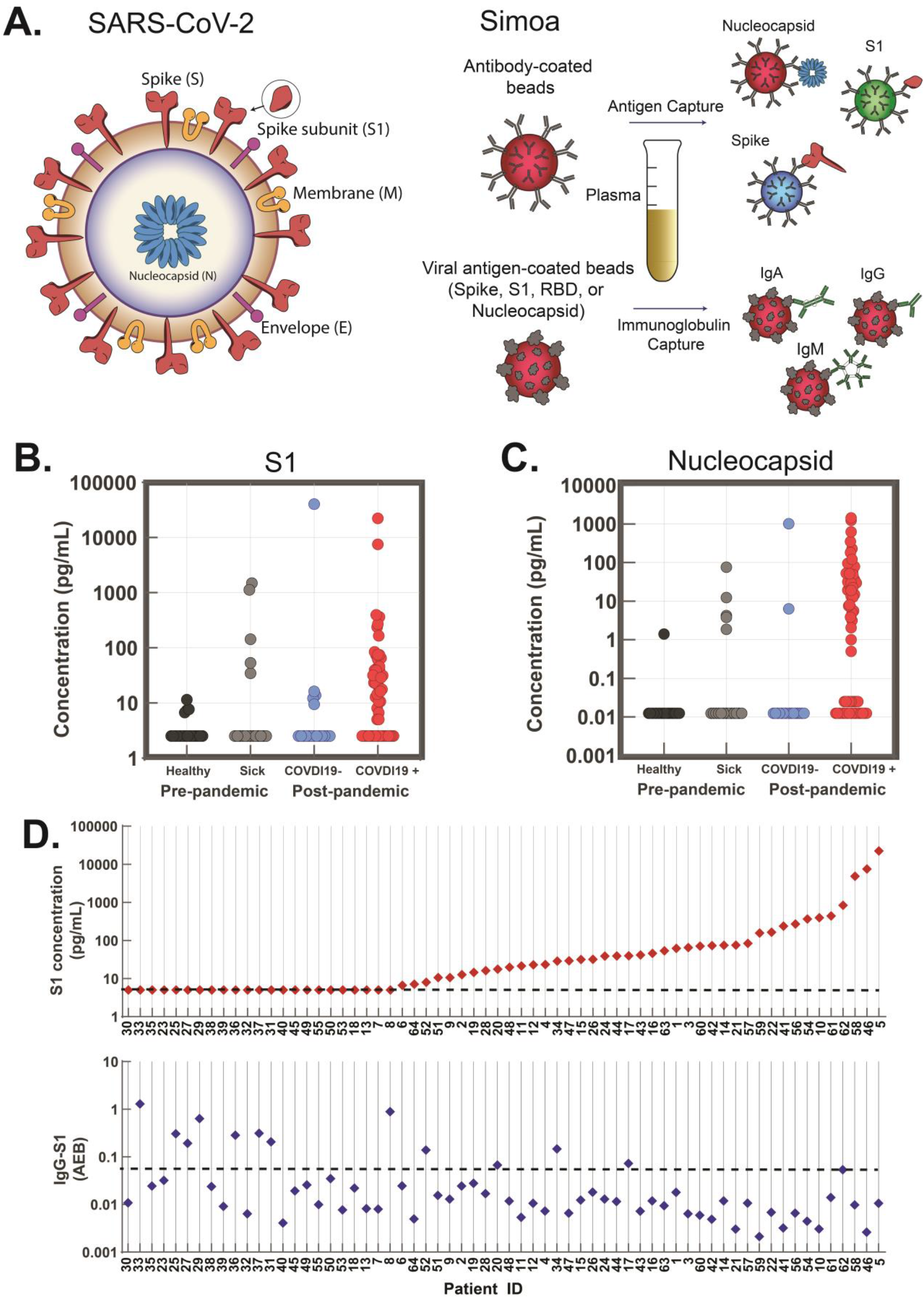
SARS-CoV-2 Antigen and anti-SARS-CoV-2 Immunoglobulin Detection in Plasma. a.) Schematic of Simoa detection of SARS-CoV-2 S1, spike, and N antigens and anti-SARS-CoV-2 immunoglobulins IgG, IgA, and IgM. Measurements for all antigens and immunoglobulins can be obtained from a single plasma sample (70 µL). b-c) Simoa SARS-CoV-2 antigen assay results for plasma samples collected from pre-pandemic healthy patients, pre-pandemic sick patients, COVID-19 negative patients, and COVID-19 positive patients. 41 of 64 COVID-19 positive patients show detectable b.) S1 and c.) N levels in plasma. Each data point represents the average of two replicate measurements. d.) Top: S1 levels plotted for individual COVID-19 positive patients, data points represent the average concentration values from two replicate measurements. Dotted lines represent the LOD for S1 assay (5 pg/mL). Bottom: IgG against S1 levels plotted for individual COVID-19 positive patients, data points represent average AEB (average enzyme per bead) value from two replicate measurements. Dotted line represents threshold (AEB >0.1) based on a cut off value that yielded 100% specificity compared to pre-pandemic healthy sample cohort (Fig. S6). Patient IDs labeled 1-64.

To probe for the presence of viral antigens in plasma, we tested samples from COVID-19 positive patients using our SARS-CoV-2 Simoa antigen assays for S1, spike, and N. These patients were determined to be COVID-19 positive bys NP RT-PCR and all plasma samples were obtained within the first ten days of the initial NP RT-PCR test. Both S1 and N were detected in 41 of 64 COVID-19 positive patients (Fig. 1b and 1c), who we identify as “viral antigen positive.” We observe S1 and N concentrations spanning four orders of magnitude in these plasma samples. Despite the presence of S1 and N in some samples, spike was only detectable in 5 of 64 COVID-19 positive patients (Fig. S4). Spike may be undetectable in some samples since the LOD is one order of magnitude higher than the LOD of the S1 assay. Additionally, in the Simoa assay for spike, the formation of a full immunocomplex depends on spike binding to the S2 subunit capture beads and the S1 subunit detection antibody. Therefore, we hypothesize that free spike antigen in plasma is likely proteolytically cleaved, releasing the S1 subunit, and the remaining spike protein fragment is undetectable by our assay.

Cross-reactivity of the SARS-CoV-2 antigen assays was assessed using samples from three control patient cohorts: (1) samples from individuals who tested negative for COVID-19 by NP RT-PCR, (2) pre-pandemic plasma samples from healthy individuals with no recorded respiratory infection in their medical history at the time of collection, collected before October 1, 2019, and (3) pre-pandemic samples collected from adults with a documented respiratory infection (including bacterial and viral pneumonia) within two months of collection, collected before October 1, 2019. Viral antigen concentrations are plotted for each individual patient in Fig. S5. In the pre-pandemic sick cohort, either S1 or N were detected in 9 of 14 individuals, indicating cross reactivity with other viruses, whereas only one sample from this cohort had both detectable S1 and N levels. Only 1 of 20 individuals from the pre-pandemic healthy cohort had detectable levels of both S1 and N. In COVID-19 negative patients, S1 or N was detectable in 6 of 18 individuals. However, analytical specificity was improved when assessing both S1 and N, resulting in only 1 of 18 COVID-19 negative patients with detectable levels of both viral antigens. We attribute detection of viral antigens in COVID-19 negative patients to either (1) assay cross-reactivity with other coronaviruses or (2) the patients with a negative NP RT-PCR test were COVID-19 positive but received a false negative result.(*10*) Currently, only two SARS-CoV-2 antigen tests are approved by the FDA and these tests qualitatively detect N antigen in NP swab samples. However, the FDA cautions that a positive result does not rule out infection with other viruses, due to cross-reactivity with other coronaviruses. Based on our preliminary results, a multiplexed approach for the detection of S1 and N protein may enhance analytical specificity of these antigen tests.

The presence of SARS-CoV-2 antigens in plasma can provide unique insight into disease progression in patients. However, it remains unclear why only a subset of patients shows detectable levels of antigen in plasma. To address this question, we tested the same set of samples using the SARS-CoV-2 serological Simoa assays for IgA, IgM, and IgG to examine correlations between antigen and immunoglobulin levels. For COVID-19 positive patients, immunoglobulin levels vary by four orders of magnitude (Fig. S6).(*9*) We observe an inverse correlation when comparing immunoglobulin levels and antigen levels for each COVID-19 positive patient (Fig. 1d). Corresponding N levels for each individual patient are plotted in Fig. S7. In several patients with S1 levels below the limit of detection, immunoglobulin levels are high. Patients with high S1 levels typically have IgG levels below the seroconversion threshold. However, some patients show detectable levels of both S1 and IgG, whereas other patients show no detectable S1 or IgG. These data highlight the diverse viral and immune responses present among COVID-19 patients.

To understand how antigen and immunoglobulin levels change with time after infection, we performed longitudinal studies in COVID-19 positive patients to monitor viral antigen and immunoglobulin levels during the course of hospitalization. This can be informative because COVID-19 patients present to the hospital at various stages of disease and show diverse progression patterns. We measured viral antigen levels (S1, spike, and N) and immunoglobulin levels (total IgA, IgM, and IgG and IgG1-4) in serial plasma samples from 39 admitted patients at Brigham and Women’s Hospital and Massachusetts General Hospital (patient IDs 1-39). All patients were diagnosed as COVID-19 positive by NP RT-PCR and serial plasma samples were collected between 0 and 30 days after the first positive NP RT-PCR test during hospitalization. SARS-CoV-2 antigen and serological Simoa results for 31 biomarkers from all longitudinal samples are presented in Fig. S8. Longitudinal antigen and immunoglobulin levels for four representative patients are plotted in Fig. 2; plots for the remaining 35 patients are shown in the Supplemental Materials. For each patient, we plot: (1) the total IgA, IgM, and IgG levels against four viral antigens: N, receptor binding domain (RBD), S1, and spike, for a total of 12 interactions, (2) N antigen levels with IgA, IgM, and IgG against N, (3) S1 antigen levels with IgA, IgM, and IgG against S1, and (4) S1 antigen levels with IgG1-4 against S1. In this patient cohort, 25 of 39 patients had detectable levels of both S1 and N antigen in their plasma, whereas only 6 of 39 patients had detectable levels of spike in their plasma during their hospitalization.

**Figure 2.**
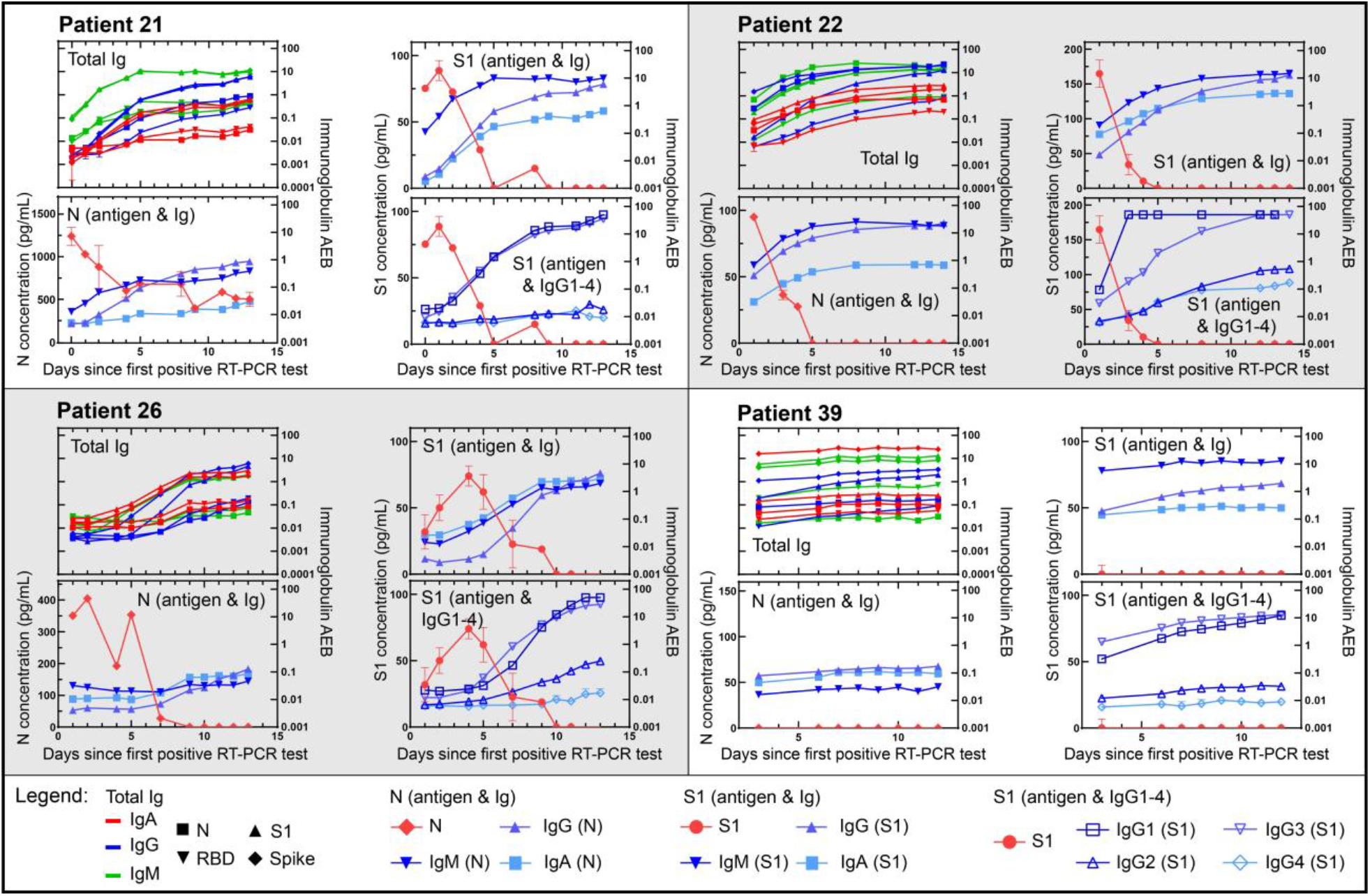
Serial data for four individual COVID-19 positive patients from admission to clinical recovery or death. Simoa antigen and serological results for serial plasma samples. (1) the total IgA, IgM, and IgG levels against four viral antigens: nucleocapsid (N), receptor binding domain (RBD), S1, and spike, for a total of 12 interactions, (2) N antigen levels with IgA, IgM, and IgG against N, (3) S1 5 antigen levels with IgA, IgM, and IgG against S1, and (4) S1 antigen levels with IgG1-4 against S1. Data points represent the average concentration or AEB from two replicate measurements and error bars represent the standard error of the mean from two replicate measurements.

The ultra-sensitivity of Simoa assays provides quantitative resolution of viral antigen and antibody levels for a detailed view of patient responses over time. For example, in Fig. 2, Patient 21 originally had high concentrations of N (1,240 pg/mL) and S1 (75.3 pg/mL) and low concentrations of anti-SARS-CoV-2 immunoglobulins in their plasma on Day 0. As patient 21 mounted an antibody response against the virus, indicated by the increase in immunoglobulin levels over time, we observe a decrease in viral antigen levels in plasma. The ultra-sensitivity of these SARS-CoV-2 Simoa assays enables us to measure even the earliest stages of seroconversion. Seroconversion was determined as previously described (See Methods for description)(*9*). This trend is observed for a majority of viral-antigen positive patients (Patient 21, 22, and 26 in Fig. 2 and Patients in Figs. S10, S12-14, S19-20, S23, S26, S31, S34, S36). As shown in Fig. 2, among Patients 21, 22, and 26, once a patient has seroconverted and their plasma immunoglobulin levels reach a steady state, there are typically no detectable levels of viral antigen. All patients seroconvert an average of 7 ± 1 days (95% CI) after the first NP RT-PCR positive test, in agreement with previous serological studies.(*11*–*14*) We defined viral antigen clearance as the first day that both S1 and N were undetectable in patient’s plasma. For viral antigen positive patients, full antigen clearance in plasma is observed 5 ± 1 days (95% CI) after seroconversion.

Furthermore, we observe a subset of patients with no detectable viral antigen levels in their plasma throughout their hospitalization. Notably, 13 of 15 patients with undetectable viral antigen levels in plasma were already seroconverted at their first NP RT-PCR test. For example, Patient 39 in Fig. 2 showed no detectable N or S1 in plasma over ten days during hospitalization. Patient 39 had already seroconverted when the first plasma sample was collected, an observation consistent for a majority of viral antigen negative patients (Fig. S11, S15-17, S18, S24,S27-28, S30-33, S35, S7-44).We propose three possibilities for the lack of detectable antigen in some patient plasma: (1) patients are presenting to the hospital after seroconversion, and therefore most viral antigens have been cleared from the plasma, (2) patients do not progress to more severe forms of the disease and therefore do not have viral antigen leakage into the blood, or (3) viral antigens are present but are bound to an immunoglobulin and the immunoglobulin blocks a binding epitope of the capture or detection antibody, resulting in an antigen-immunoglobulin complex that is undetectable by our Simoa assays. However, even with no detectable levels of viral antigen, 12 of 16 patients were admitted or transferred to the ICU during hospitalization and intubated, indicating that these patients were severe cases. Therefore, we propose a third possibility where patients have seroconverted and despite reaching viral clearance, suffer from severe respiratory damage that inhibits recovery.

To further analyze the 31 biomarkers measured in these longitudinal samples, we performed a cluster map analysis to determine if antigen and immunoglobulin levels in COVID-19 positive patients could differentiate different stages of infection. We used a Ward variance minimization algorithm (Supplemental Materials Methods) to perform hierarchical clustering. The cluster map shows three dominant branches (Fig. 3) representing three clusters of samples that are grouped based on similarity between the 31 biomarker values. The first branch is observed for early time points where we observe an inverse relationship between high levels of viral antigens and low levels of antibodies. The second branch shows a transition in patient samples where antigen levels begin to decrease as total IgM, IgA, and IgG (as well as IgG subclasses IgG1 and IgG3) begin to increase. In the third branch, we see an even further reduction in antigen levels with small increases in IgG subclasses IgG2 and IgG4 (relative to their average values across samples). The third branch is observed for later time points where we propose that patients have cleared viral antigens from the plasma and antibodies have reached their steady-state levels. For all 39 patients in this cohort, we observe that IgG1 and IgG3 show large increases in plasma concentrations over time. In contrast, IgG2 and IgG4 show little to no changes above background levels over time (note that cluster map data is standardized so all values are relative for each biomarker). Both IgG1 and IgG3 are subclasses of IgG that predominantly respond to viral antigens and mediate neutralization.(*15*–*19*) Based on these measurements, the IgG response that is mounted by the immune system is primarily mediated by IgG1 and IgG3 for SARS-CoV-2, in agreement with previous serological studies.(*4, 5*) The neutralization effects of IgG1 and IgG3 on viral antigens should be explored in future studies that include correlative neutralization titer assays.

**Figure 3.**
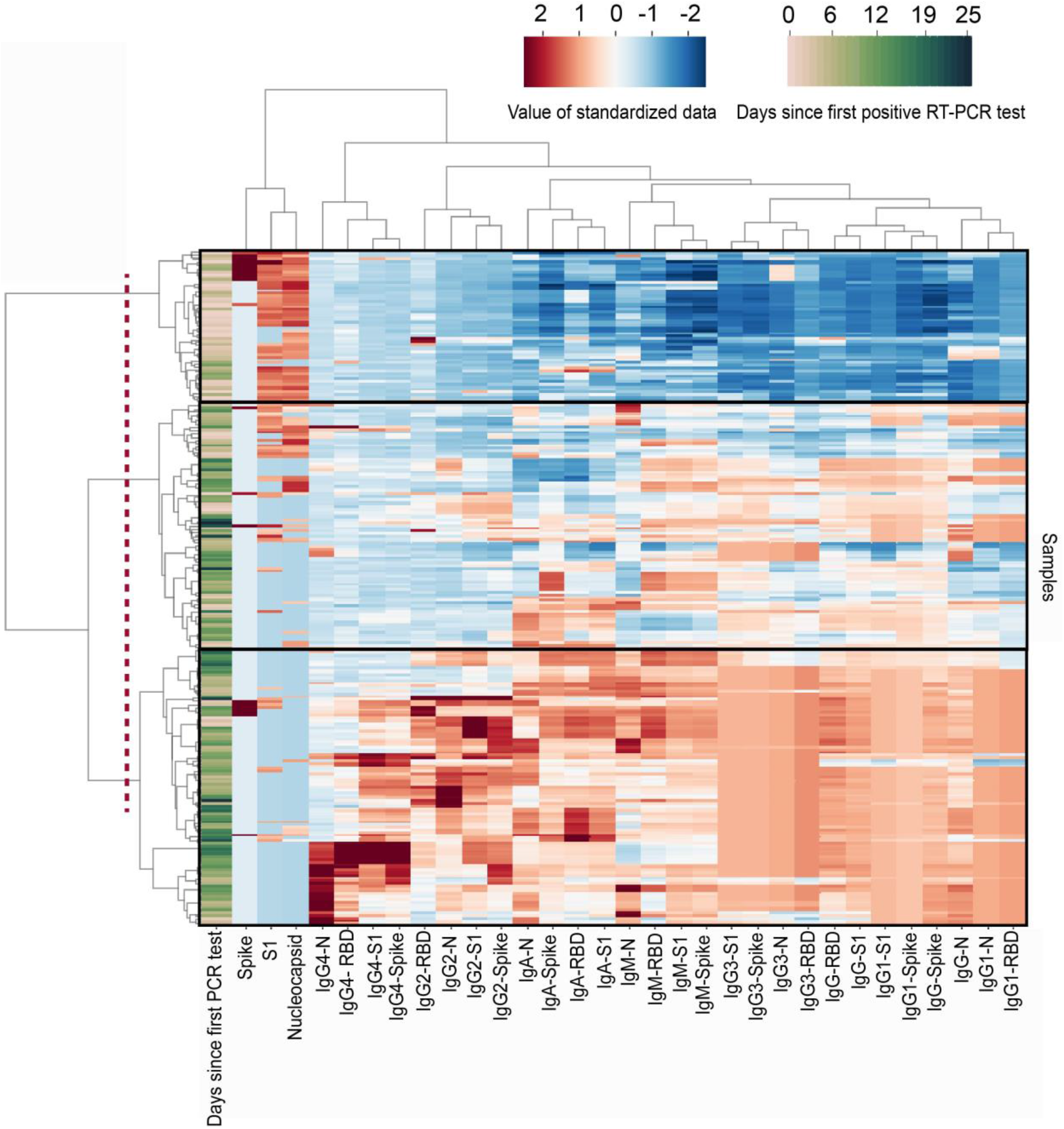
Clustergram of Simoa antigen and immunoglobulin results from 252 samples from 39 patients. Cluster map produce by Ward Variance Minimization algorithm. 252 samples from 39 patients were analyzed as described in the Supplemental Materials Methods section. Data were standardized after a non-linear transformation and each row represents a single sample. Days since first RT-PCR test for each 5 sample are presented at the far left of the clustergram. Longitudinal samples are clustered by dendrograms to the left of the clustergram. SARS-CoV-2 antigen and immunoglobulin markers are clustered by dendrograms at the top of the clustergram. The dotted line represents the cutoff that results in three branches: branch 1 (top), branch 2 (middle), and branch 3 (bottom). RBD: Receptor binding domain. N: Nucleocapsid.

In addition to showing the inverse relationship between antigen levels and seroconversion in this patient cohort, we explored how antigen clearance compared with longitudinal RT-PCR tests and clinical outcomes. The measured SARS-CoV-2 antigen and immunoglobulin levels in plasma, NP RT-PCR tests, and select clinical outcomes for each of these patients are displayed in Fig. 4 and patient features are summarized in Table S4. For each patient in Fig. 4, the date that each plasma sample was collected is indicated by an X. Plasma samples with significant detectable levels of antigen (concentrations two times above the LOD) are indicated with a black X and plasma samples with no significant detectable levels of antigen are indicated with a blue X. In this patient cohort, NP RT-PCR tests reported positive results for an average of 15 ± 5 days after viral antigen clearance and 18 ± 4 days after seroconversion. Furthermore, several patients never received a negative RT-PCR result before being discharged from the hospital. These observations are in agreement with recent virological studies that confirmed persistence of viral RNA and showed how seroconversion was not immediately followed by a decrease in NP viral RNA in hospitalized patients.(*20*) Because RNA shedding can occur for several weeks after infection and recovery, measurements of viral antigens and immunoglobulins may provide more accurate and timely indicators of disease progression and recovery compared to viral RNA.

**Figure 4.**
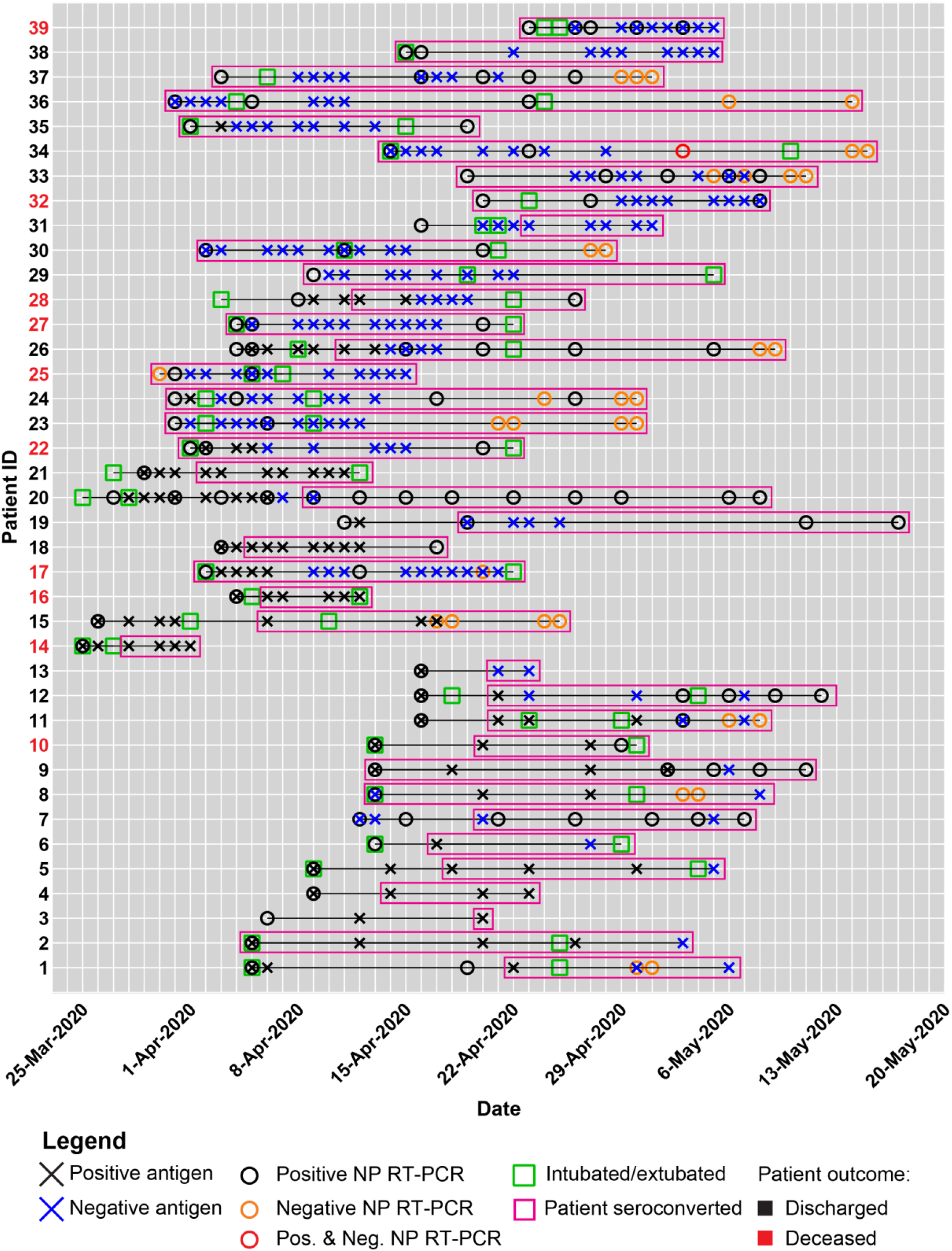
Summary of clinical data and Simoa SARS-CoV-2 antigen and immunoglobulin assays for individual patients in the longitudinal analysis. A black X indicates a viral antigen positive test, whereas a blue X indicates a viral antigen negative test. A black circle indicates a positive NP RT-PCR test, whereas an orange circle indicates a negative NP RT-PCR test. For example, on May 3, 2020, Patient 34 received 5 both a positive and negative NP RT-PCR result, indicated by the red circle. The intubation and extubation dates of each patient are indicated by green squares. Patient IDs are coded black for recovered patients who were discharged from the hospital, whereas patient IDs are coded red for deceased patients. Pink boxes represent the date of seroconversion for each patient.

Finally, we demonstrate that the presence of viral antigens in plasma correlates with certain indicators of disease severity in COVID-19 patients. To understand the correlation between SARS-CoV-2 antigen levels and disease severity, clinical features for the 64 COVID-19 positive patients (Fig. 5, patient IDs 1-64) were compared with viral antigen levels in plasma. It is important to note that all samples were obtained from patients admitted to the hospital, resulting in a cohort of primarily severe cases. For patients with serial samples, we analyzed the first plasma sample collected after the initial positive NP RT-PCR test. Viral-antigen-positive patients show detectable levels of both S1 and N in plasma; however, S1 shows higher correlation with clinical severity (Table S5-8, Fig. S45). Therefore, patients were grouped into three categories of S1 concentrations: (1) 23 patients with undetectable S1 levels (below the limit of detection), (2) 23 patients with low levels of S1 (6-50 pg/mL), and (3) 18 patients with high levels of S1 (>50 pg/mL). The cutoff between groups 2 and 3 (50 pg/mL) was chosen as five standard deviations above the LOD. There is a significant difference in rates of ICU admission upon presentation for patients with varying levels of S1 in plasma (p value: 0.0107). Patients with zero, low, and high levels of S1 were admitted to the ICU upon presentation to the hospital at rates of 30%, 52%, and 77%, respectively (Fig. 5a). However, 13 of 19 patients initially admitted to non-ICU dispositions were later transferred to the ICU during hospitalization, as summarized in Table S4. Among all COVID-19 positive patients, over 60% were intubated during hospitalization with no statistically significant difference in intubation rates among groups (Fig. 5b). All patients with high levels of S1 were intubated within one day of hospitalization, whereas patients with zero and low S1 levels were intubated within a range of zero to ten days post-admission (Fig. 5c). The difference in mean times to intubation between patients with high levels of S1 and patients with no detectable S1 is significant (p-value: 0.0050). These results suggest that high S1 levels in plasma upon presentation to the hospital correlate with severe cases of COVID-19 that can result in respiratory failure and require immediate intubation. Among all patients, a broad range of intubation periods, up to 32 days, was observed (Fig. 5d). Once intubated, there is no statistically significant difference in mean intubation periods among patient groups. No statistically significant difference in death rate was observed for patients with zero, low, and high levels of S1, where 5 of 23, 5 of 23, and 8 of 18 of patients died, respectively (p-value: 0.1921). Although six patients showed detectable levels of spike protein, there was no correlation between spike concentrations and ICU admission, intubation rate, or death rate. These results show a high level of diversity and complexity in the responses and disease trajectories in each patient. There are potentially more features in these data that will lead to further correlations, but a large cohort of patients will need to be tested to elucidate other patterns.

**Figure 5.**
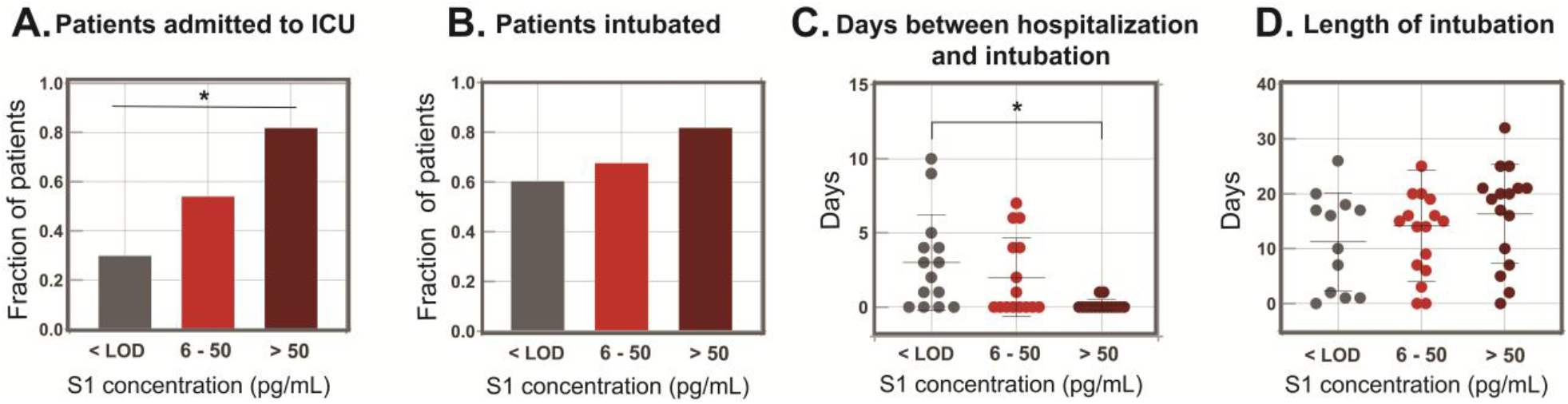
Indicators of disease severity based on S1 levels in plasma for 64 COVID-19 positive patients. a.) Fraction of COVID-19 positive patients who were immediately admitted to the ICU upon presentation to the hospital. b.) Fraction of COVID-19 positive patients who were intubated during hospitalization. c.) Days between date of presentation to the hospital and intubation date for intubated 5 COVID-19 positive patients. d.) The length of intubation for intubated COVID-19 positive patients. For all plots, significance indicated by the asterisks (p value <0.05).

Compared to RT-PCR tests, immunoassay-based antigen and serological tests have the advantage of speed and simplicity when integrated into point-of-care platforms. Plasma is a readily available biofluid from hospitalized patients but is more difficult to obtain in non-clinical settings. Saliva is a non-invasive biofluid that is easier to use for wide-scale testing. Therefore, we explored the presence of SARS-CoV-2 antigens and anti-SARS-CoV-2 immunoglobulins in saliva (Fig. S47, S48, S49). We tested 19 saliva samples from patients presenting to the Emergency Department and show that 8 of 19 patients have detectable levels of S1. Similar to our observations of antigen and immunoglobulin levels in plasma, saliva S1 and anti-SARS-CoV-2 antibody levels show negative correlation. These initial results indicate the presence of SARS-CoV-2 antigens and anti-SARS-CoV-2 immunoglobulins in saliva and highlight the potential for adapting our assays to a diagnostic test for COVID-19. Future studies on saliva will include longitudinal sample analysis for mild and severe cases of COVID-19 patients and will explore the potential for developing a saliva-based COVID-19 antigen screening tool.

Using SARS-CoV-2 Simoa assays, we demonstrate quantitative detection of SARS-CoV-2 antigens and anti-SARS-CoV-2 immunoglobulins in plasma of COVID-19 patients. While detection of N in NP swabs has been cited,(*6*) we present the first report of SARS-CoV-2 spike, S1, and N detection in plasma. The presence of S1 and N in plasma suggest that fragments of virus are entering the bloodstream, potentially due to tissue damage. Although spike is undetectable in most COVID-19 patients, possibly due to proteolytic cleavage, six patients show high levels of spike in plasma. We hypothesize that some spike is not proteolytically cleaved, especially in cases where there are high levels of spike in plasma. No evidence has been reported yet for full viral particles in blood, though we cannot rule out this possibility.(*21*) Nonetheless, severe COVID-19 cases with acute respiratory distress syndrome can result in damage to endothelial cells and vascular leakage(*22*–*25*) and we propose that this damage can lead to discharge of viral antigens into the blood. Patients with lung damage can suffer from respiratory failure and require intubation or mechanical ventilation. We also report significant correlation between high S1 levels in plasma and time between hospital admission and intubation. In situations where ICUs have limited capacity, there is urgent need to prioritize severe COVID-19 patients. Our initial results suggest that detecting viral antigen levels in plasma could aid in patient stratification and may indicate when patients require immediate medical intervention, such as intubation. Although we hypothesize that asymptomatic or mild cases will likely not show viral antigen levels in plasma, future studies that include mild cases will be used to probe SARS-CoV-2 antigen and antibody levels over time in comparison to the severe cases presented here.

## Data Availability

All Data is available within the manuscript and Supporting Information.

## Acknowledgements

The authors acknowledge Dr. Sanjat Kanjilal for collecting saliva samples.

## Author Contributions

A.F.O, A.M.M., C.W., T.G., M.N., and D.R.W. conceived the approach. A.F.O, A.M.M., C.W., T.G., M.N., and G.N. performed the experiments, A.F.O., A.M.M., C.W., T.G., M.N., R.L., C.P.M., and T.E.G. analyzed the data, M.C., K.N., M.K., Q.Z., and W.A.M. produced and purified the anti-S1 antibody, R.C.C. and E.T.R. collected the samples and performed chart review for patients from Massachusetts General Hospital, C.P.M. performed chart review for patients from Brigham and Women’s Hospital. A.F.O., A.M.M., and D.R.W. co-wrote the paper. All authors were involved in designing experiments, reviewing and discussing data, and commented on the manuscript.

## Competing Interests

David Walt has a financial interest in Quanterix Corporation, a company that develops an ultra-sensitive digital immunoassay platform. He is an inventor of the Simoa technology, a founder of the company and also serves on its Board of Directors. Dr. Walt’s interests were reviewed and are managed by BWH and Partners HealthCare in accordance with their conflict of interest policies.

The anti-SARS-CoV-2 Simoa assays in this publication have been licensed by Brigham and Women’s Hospital to Quanterix Corporation.

## Supplementary Materials

### I. Methods

#### Plasma Samples

Clinical samples were obtained from adult patients presenting to Brigham and Women’s Hospital through the Partner’s biobank and patients from Massachusetts General Hospital. 34 samples termed pre-pandemic, defined by a collection date before October 1, include 20 samples from healthy patients and 14 samples from sick patients with upper respiratory infections, bacterial pneumonia, viral pneumonia, or unspecified virus positive. 17 samples were from patients who tested negative for SARS-CoV-2 using nasopharyngeal RT-PCR. 64 samples were from patients who tested positive for SARS-CoV-2 using nasopharyngeal RT-PCR.

Serial timepoint clinical samples were obtained from patients admitted to Massachusetts General Hospital (n= 67 samples from 19 individual patients) and Brigham and Women’s Hospital (n= 205 samples from 24 patients) diagnosed as SARS-CoV-2 positive by a nasopharyngeal RT-PCR. Several timepoint samples were taken 0-30 days after the first SARS-CoV-2 positive nasopharyngeal RT-PCR test per patient.

All plasma samples were not heat inactivated and diluted in Homebrew Sample Diluent (Quanterix) to their final dilution factor. All samples were collected under approval of the Partners Institutional Review Board for Human Subjects Research.

#### Anti-S1 antibody discovery and expression

The Mehta I/II 2.7×10^10^ member non-immunized scFv-phage library was used to perform three rounds of panning against SARS-CoV-2 S1 protein (Sino Biologicals, Wayne, PA). Nunc MaxiSorp Immuno tubes (Thermo Fisher Scientific, Waltham, MA) were coated with SARS-CoV-2 S1 protein overnight in PBS and the SARS-CoV-2 S1 coating concentration was decreased in each round to increase the affinity of the enriched antibodies. Screening of the enriched library was performed by picking single bacterial colonies from the 3^rd^ round of panning. Small scale rescue was performed and the phage supernatant was used to screen via SARS-CoV-2 RBD coated ELISA plates. The recovered antibody was converted to hIgG1 and expressed transiently by transfection into Expi293F cells via ExpiFectamine (Thermo Fisher Scientific, Waltham, MA). The cells were cultured for four days before the supernatant was harvested and cells pelleted. Clarified supernatant was mixed with Protein A Sepharose 4B beads (Thermo Fisher Scientific, Waltham, MA) overnight before purification via gravity flow columns. Purified protein was buffer exchanged into PBS via Amicon Ultra-15 Centrifugal Filter Units (MilliporeSigma, Burlington, MA).

#### Preparation of Viral Antigen Simoa Reagents

##### Antibody conjugation to beads

Antibodies against S1, S2, and Nucleocapsid were conjugated to carboxylated 2.7 µm paramagnetic Homebrew beads, 700 nm dye-encoded beads, and 750 nm dye-encoded beads (Quanterix), respectively. For each bead conjugation, 2.27×10^8^ beads were washed three times with 200 µL of Bead Wash Buffer (Quanterix) and then three times with 200 µL of Bead Conjugation Buffer (Quanterix). The beads were resuspended in 297 µL Bead Conjugation Buffer and 3 µL of 1-ethyl-3-(3-dimethylaminopropyl)carbodiimide hydrochloride (EDC), freshly dissolved to 10 mg/mL in Bead Conjugation Buffer, was added. The beads were shaken for 30 minutes at 4 °C and then washed once with 300 µL Bead Conjugation Buffer before resuspending in 250 µL Bead Conjugation Buffer and adding 50 µL capture antibody (1 mg/mL). The beads were shaken for two hours at 4 °C for antibody conjugation, washed twice with 200 µL Bead Wash Buffer, and blocked with 200 µL Bead Blocking Buffer (Quanterix) for 30 minutes at room temperature. After blocking, the beads were washed once with 200 µL Bead Wash Buffer and then 200 µL Bead Diluent (Quanterix), before resuspending in 200 µL Bead Diluent. The beads were counted using a Beckman Coulter Z1 Particle Counter and stored at 4 °C for subsequent use.

##### Biotinylation of detector antibodies

Each detector antibody was conjugated to EZ-Link NHS-PEG4-Biotin (Thermo Fisher Scientific) by adding 3 µL NHS-PEG4-Biotin (2 mg freshly dissolved in 383 µL deionized water) to 100 µL antibody (1 mg/mL in PBS). The mixture was incubated for 30 minutes at room temperature and then purified using a 50 kDa 0.5 mL Amicon filter (MilliporeSigma), with five washes in Biotinylation Reaction Buffer (Quanterix) at 14000xg for five minutes. The filter was then inverted and centrifuged for two minutes at 1000xg, and washed with 50 µL Biotinylation Reaction Buffer before centrifuging again for two minutes at 1000xg. The biotinylated antibody concentration was measured with a NanoDrop spectrophotometer.

#### Simoa assays

##### Antigen Assays

Simoa assays were performed on an HD-X Analyzer (Quanterix) in an automated three-step assay format according to the manufacturer’s instructions and as previously described. (*24*) Plasma samples were diluted 8-fold in Homebrew Detector/Sample Diluent (Quanterix Corp.) with Halt Protease Inhibitor Cocktail (Thermo Fischer Scientific) and EDTA. Detector antibodies were diluted in Homebrew Detector/Sample Diluent to 0.3 µg/mL, and streptavidin-β-galactosidase (SβG) concentrate (Quanterix) was diluted to 150 pM in SβG Diluent (Quanterix). Antibody-conjugated capture beads were diluted in Bead Diluent, with a total of 500,000 beads per reaction (125,000 S1 beads, 125,000 S2 beads, and 250,000 647 nm dye-encoded helper beads for the S1/S2 multiplex assay, and 125,000 nucleocapsid beads and 375,000 647 nm dye-encoded helper beads for the nucleocapsid assay). All reagents were diluted in plastic bottles that were loaded into the HD-X Analyzer, and all assay steps were performed in an automated manner on the instrument. In each assay, capture beads were incubated with the sample for 15 minutes, detector antibody for 5 minutes, and SβG for 5 minutes, with washing steps in between. The beads were then resuspended in 25 µL resorufin-β-galactopyranoside and loaded into the microwell array for imaging. Average enzyme per bead (AEB) and sample concentration values were calculated by the HD-X Analyzer software. All samples were measured in duplicates.

##### Immunoglobulin Assays

SARs-CoV-2 serological Simoa assays were prepared and preformed as previously described.(*9*) Plasma samples were diluted 4000-fold in Homebrew Detector/Sample Diluent (Quanterix Corp.) SARs-CoV-2 serological Simoa assays for IgG1, IgG2, IgG3, and IgG4 were developed against 4 viral antigens S1, N, spike, and RBD as previously described. The detection antibodies from commercial sources are listed below. Simoa assays were performed on an HD-X Analyzer (Quanterix) in an automated three-step assay format according to the manufacturer’s instructions and as previously described.(*8*) Plasma samples were diluted 500-fold in Homebrew Detector/Sample Diluent (Quanterix Corp.)

**Table S1.** Manufacturer catalog information for commercial IgG subclass antibodies.

| Antibody Target | Manufacturer | Catalog Number |
| --- | --- | --- |
| IgG1 | Biolegend | 409904 |
| IgG2 | Biolegend | 411102 |
| IgG3 | R&D Systems | 10122 |
| IgG4 | Biolegend | 411202 |

#### Data Analysis

##### Seroconversion Thresholding

Seroconversion classification based upon the early stage classification model trained using an independent panel of 142 positive samples by RT-PCR SARS-Cov-2 and 200 negative pre-pandemic controls. The markers for this model were chosen using a cross-validation step. This cross-validation yielded four markers (IgA S1, IgA Nucleocapsid, IgG Nucleocapsid, and IgG Spike) and exhibited the best performance in the training set. The threshold for a positive test result for the unknown samples was determined based on the cutoff that yielded 100% specificity in the training set. (*9*)

##### Standardizing the biomarkers for visualization and downstream analysis

This standardization was performed so that all the biomarkers can be placed on the same scale. Throughout the text when we refer to standardized data, the following procedure was performed. First, the minimum nonzero value for each biomarker was stored, then the resulting vector was divided by 100 and added to every samples corresponding biomarker value, which is necessary for log transformation. The data was log transformed and then a second transformations step was performed such that each biomarker across the samples has an empirical mean of zero and variance of one. This is achieved by computing the mean and variance for each biomarker, subtracting the mean, and then dividing by the variance.

##### Cluster Map Analysis

The cluster map analysis was performed on the standardized data. Samples were removed if they had any missing values (reduced the number of samples from 272 to 252). Specifically the cluster map is a hierarchical clustering based on the Ward variance minimization algorithm, (https://docs.scipy.org/doc/scipy/reference/generated/scipy.cluster.hierarchy.linkage.html).

### II. Antigen Assay Development and Validation

**Table S2.** Manufacturer catalog information for commercial viral antigen antibodies and recombinant protein standards.

| SARS-CoV-2 Recombinant Protein | SARS-CoV-2 Antibody | Manufacturer | Catalog Number |
| --- | --- | --- | --- |
| Nucleocapsid | - | RayBiotech | 230-30164 |
| S1 | - | Sino Biological | V0591-V08H |
| - | Nucleocapsid (capture Ab) | Sino Biological | 40143-R004 |
| - | Nucleocapsid (detection Ab) | Sino Biological | 40143-R040 |
| - | S1 (capture Ab) | Sino Biological | 40150-D001 |
| - | S2 (capture Ab) | Sino Biological | 40590-T62 |

#### Antigen assay calibration curves

**Figure S1.**
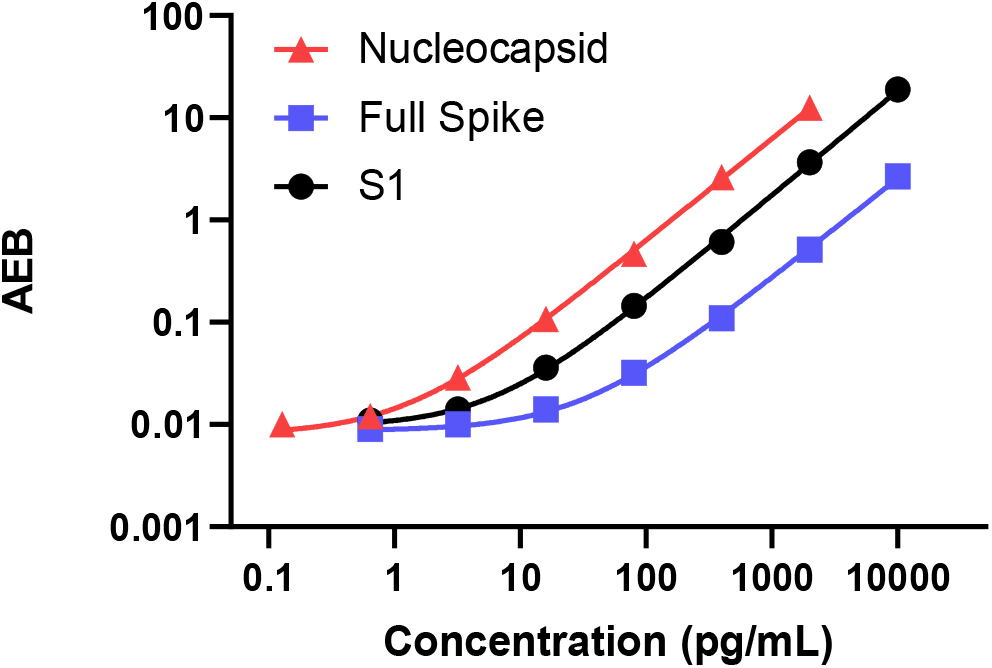
Calibration curves for the two SARS-CoV-2 antigen assays: (1) single-plex nucleocapsid assay and (2) two-plex Spike (S1/ Spike) assay. Error bars represent the standard 10 deviations of duplicate measurements. Some errors bars are narrow and encompassed within the data point.

#### Cross-reactivity validation of two-plex Spike (S1/ Spike) assay

**Figure S2.**
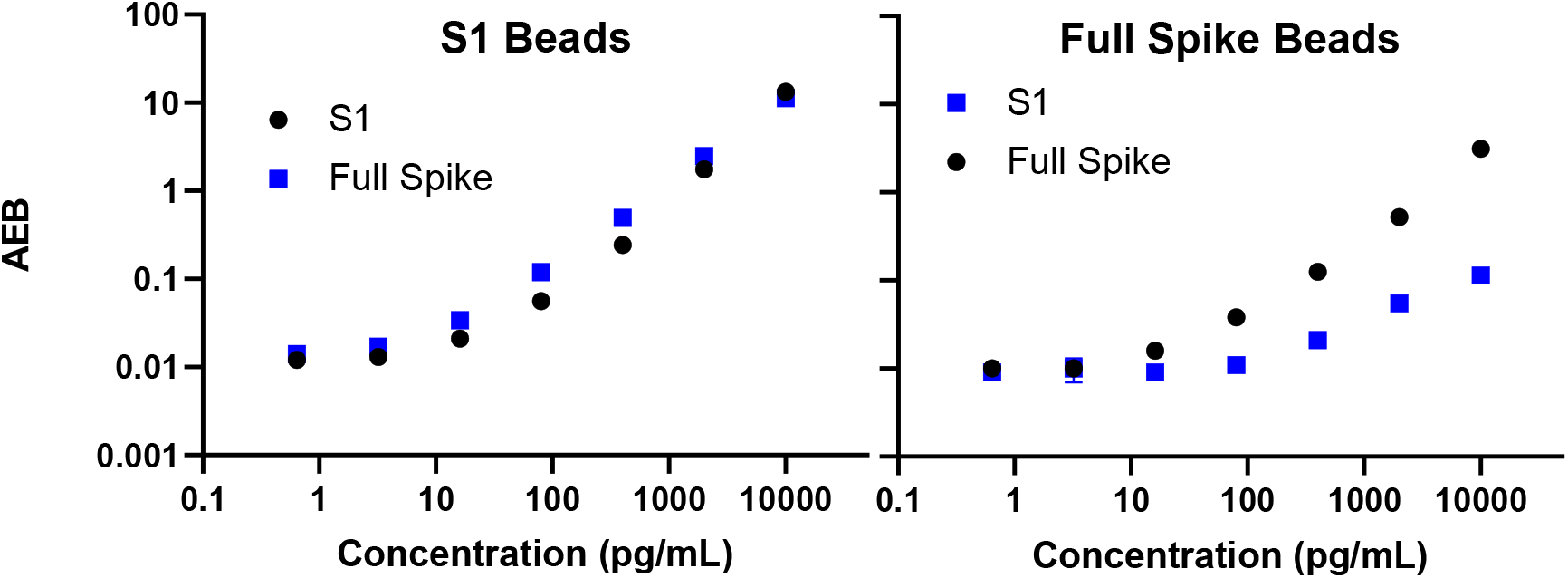
Cross-reactivity validation of two-plex Spike (S1/ Spike) assay. Left: S1 assay (S1 capture and S1 detector) captures both S1 and Spike. Right: Spike assay (S2 capture and S1 detector) captures Spike. There is cross-reactivity in the Spike assay at high concentrations 5 (>400 pg/mL) of S1. All samples measured in this study had S1 plasma concentrations below 400 pg/mL. Error bars represent the standard deviations of duplicate measurements. Some errors bars are narrow and encompassed within the data point.

#### Dilution Linearity and Spike & Recovery

Healthy K2EDTA human plasma (pooled gender) was purchased from BioIVT for assay validation experiments (Spike & Recovery and Dilution Linearity). Three samples (three different sets of pools) were used for all validation experiments. For all experiments, plasma samples were diluted with Homebrew Detector/Sample Diluent (Quanterix) with 1X Halt Protease Inhibitor Cocktail (ThermoFisher Scientific #78438). For Dilution Linearity experiments, plasma samples were diluted to 4X and spiked with a final concentration of either (1) 2,000 pg/mL nucleocapsid or (2) 2,000 pg/mL S1 and 2,000 pg/mL Spike. Samples were then serially diluted to 8X, 16X, and 32X. Antigen concentrations are plotted as a function of 1/Dilution Factor below in Figure S3. Data for each sample were fit to a linear regression and R^2^ values were calculated. R^2^ values for all three plasma samples were >0.98 for S1, >0.95 for Spike, and >0.98 for Nucleocapsid. Error bars represent the standard deviations of duplicate measurements. Some errors bars are narrow and encompassed within the data point.

**Figure S3.**
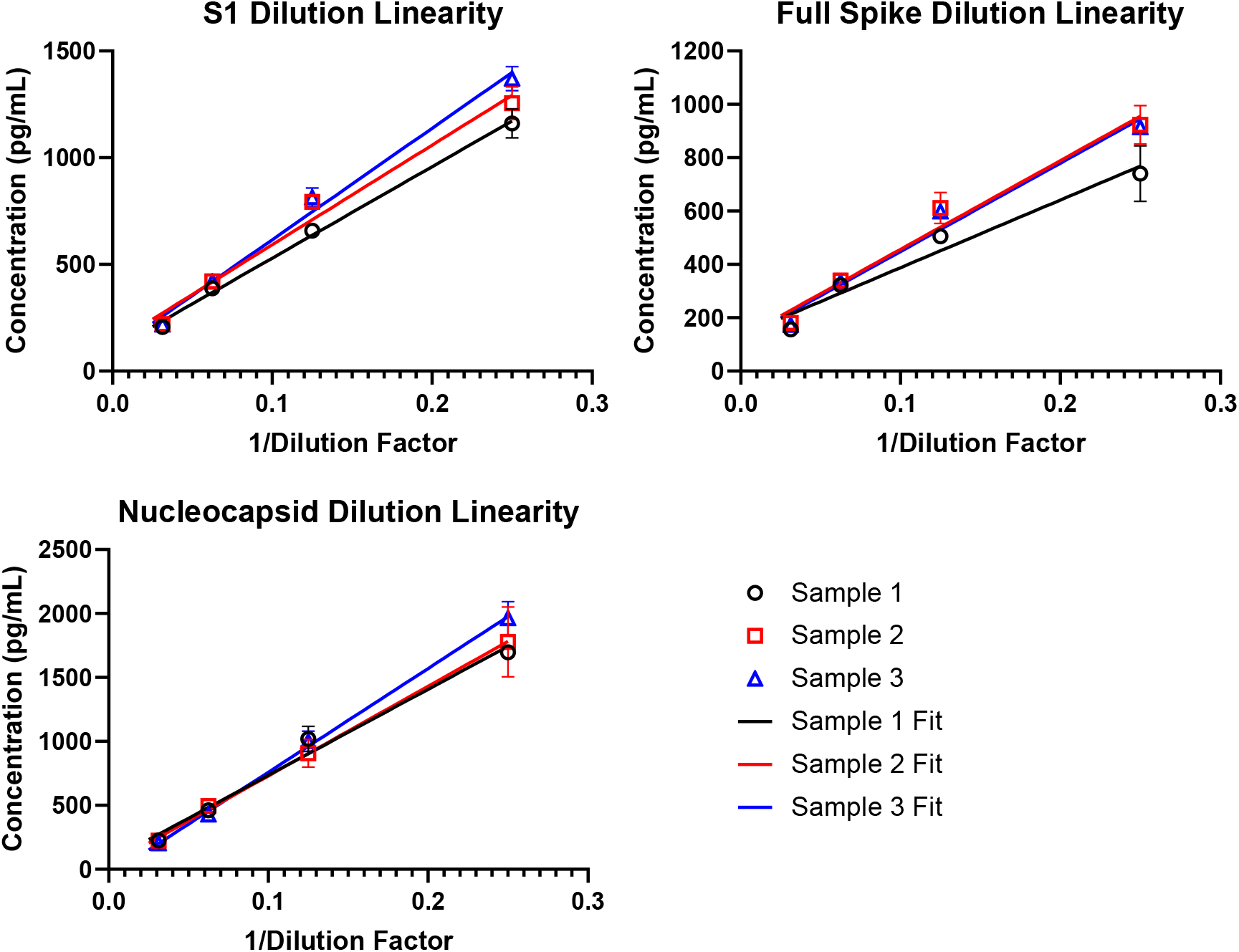
Antigen assay dilution linearity curves.

For spike and recovery experiments, plasma samples were diluted to 8X and spiked with a final concentration of (1) 2,000 pg/mL, 400 pg/mL, or 80 pg/mL nucleocapsid for the nucleocapsid 5 assay, or (2) 2,000 pg/mL, 400 pg/mL, or 80 pg/mL S1 and Spike for the S1/ Spike assay. Plasma samples at 8X dilution with no spiked recombinant protein (blank sample) were also analyzed. For each spiked sample, the concentration of antigen in each blank sample (0 pg/mL recombinant protein) was subtracted from the measured concentration before calculating percent recovery. Percent recoveries for each sample and assay are displayed below in Table S2. 10

**Table S3.** Antigen Assay Spike and Recovery.

| **Sample**<br>**(Nucleocapsid**<br>**)** | **Spiked**<br>**Concentration**<br>**(pg/mL)** | **Measured**<br>**Concentration**<br>**(pg/mL)** | **St. Dev.**<br>**(pg/mL)** | **Blank Corrected**<br>**Conc. (pg/mL)** | **N Percent**<br>**Recovery** |
| --- | --- | --- | --- | --- | --- |
| Sample 1 | 2000 | 1720 | 117 | 1720 | 86.1 |
|  | 400 | 359 | 37.7 | 359 | 89.8 |
|  | 80 | 80.9 | 0.54 | 80.8 | 101 |
|  | 0 | 0.12 | 0.00 |  |  |
| Sample 2 | 2000 | 2040 | 64.9 | 2040 | 102 |
|  | 400 | 447 | 11.8 | 446 | 112 |
|  | 80 | 78.0 | 1.46 | 77.7 | 97.1 |
|  | 0 | 0.35 | 0.05 |  |  |
| Sample 3 | 2000 | 1770 | 77.0 | 1770 | 88.4 |
|  | 400 | 403 | 22.1 | 403 | 100. |
|  | 80 | 76.3 | 10.7 | 76.0 | 94.9 |
|  | 0 | 0.30 | 0.40 |  |  |
| **Sample (S1)** | **Spiked**<br>**Concentration**<br>**(pg/mL)** | **Measured**<br>**Concentration**<br>**(pg/mL)** | **St. Dev.**<br>**(pg/mL)** | **Blank Corrected**<br>**Conc. (pg/mL)** | **S1 Percent**<br>**Recovery** |
| Sample 1 | 2000 | 1560 | 169 | 1560 | 78.1 |
|  | 400 | 268 | 5.51 | 268 | 67.0 |
|  | 80 | 51.6 | 3.92 | 51.5 | 64.4 |
|  | 0 | 0.15 | 0.00 |  |  |
| Sample 2 | 2000 | 1570 | 13.7 | 1570 | 78.2 |
|  | 400 | 307 | 1.32 | 305 | 76.3 |
|  | 80 | 67.4 | 1.14 | 66.0 | 82.5 |
|  | 0 | 1.35 | 0.86 |  |  |
| Sample 3 | 2000 | 1780 | 72.6 | 1775 | 88.7 |
|  | 400 | 315 | 4.66 | 314 | 78.4 |
|  | 80 | 63.3 | 2.75 | 61.9 | 77.4q |
|  | 0 | 1.37 | 1.26 |  |  |
| **Sample**<br>**(Spike)** | **Spiked**<br>**Concentration**<br>**(pg/mL)** | **Measured**<br>**Concentration**<br>**(pg/mL)** | **St. Dev.**<br>**(pg/mL)** | **Blank Corrected**<br>**Conc. (pg/mL)** | **Spike**<br>**Percent**<br>**Recovery** |
| Sample 1 | 2000 | 1010 | 41.5 | 1010 | 50.70 |
|  | 400 | 216 | 17.1 | 216 | 54.12 |
|  | 80 | 28.6 | 4.20 | 28.6 | 35.73 |
|  | 0 | 0.00 | 0.00 |  |  |
| Sample 2 | 2000 | 1150 | 17.2 | 1150 | 57.38 |
|  | 400 | 244 | 2.69 | 244 | 61.10 |
|  | 80 | 49.0 | 1.90 | 49.0 | 61.24 |
|  | 0 | 0.00 | 0.00 |  |  |
| Sample 3 | 2000 | 1250 | 23.1 | 1250 | 62.3 |
|  | 400 | 258 | 24.6 | 258 | 64.1 |
|  | 80 | 55.9 | 2.56 | 54.4 | 68.0 |
|  | 0 | 1.49 | 0.00 |  |  |

### III. Antigen and Immunoglobulin Data for All Patient Samples

**Figure S4.**
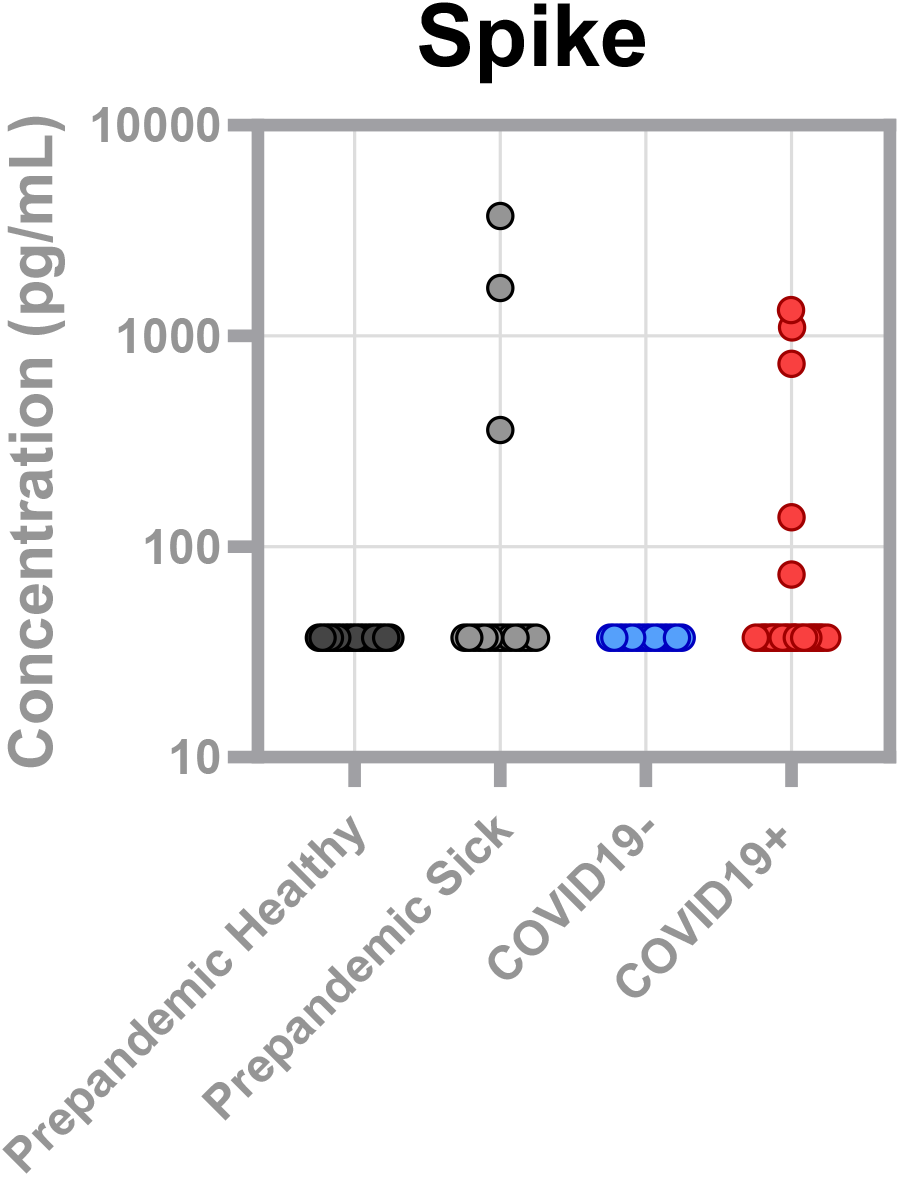
Concentration of SARS-CoV-2 spike in plasma samples from pre-pandemic healthy, pre-pandemic sick patients, COVID-19 negative patients, and COVID-19 positive patients.

**Figure S5.**
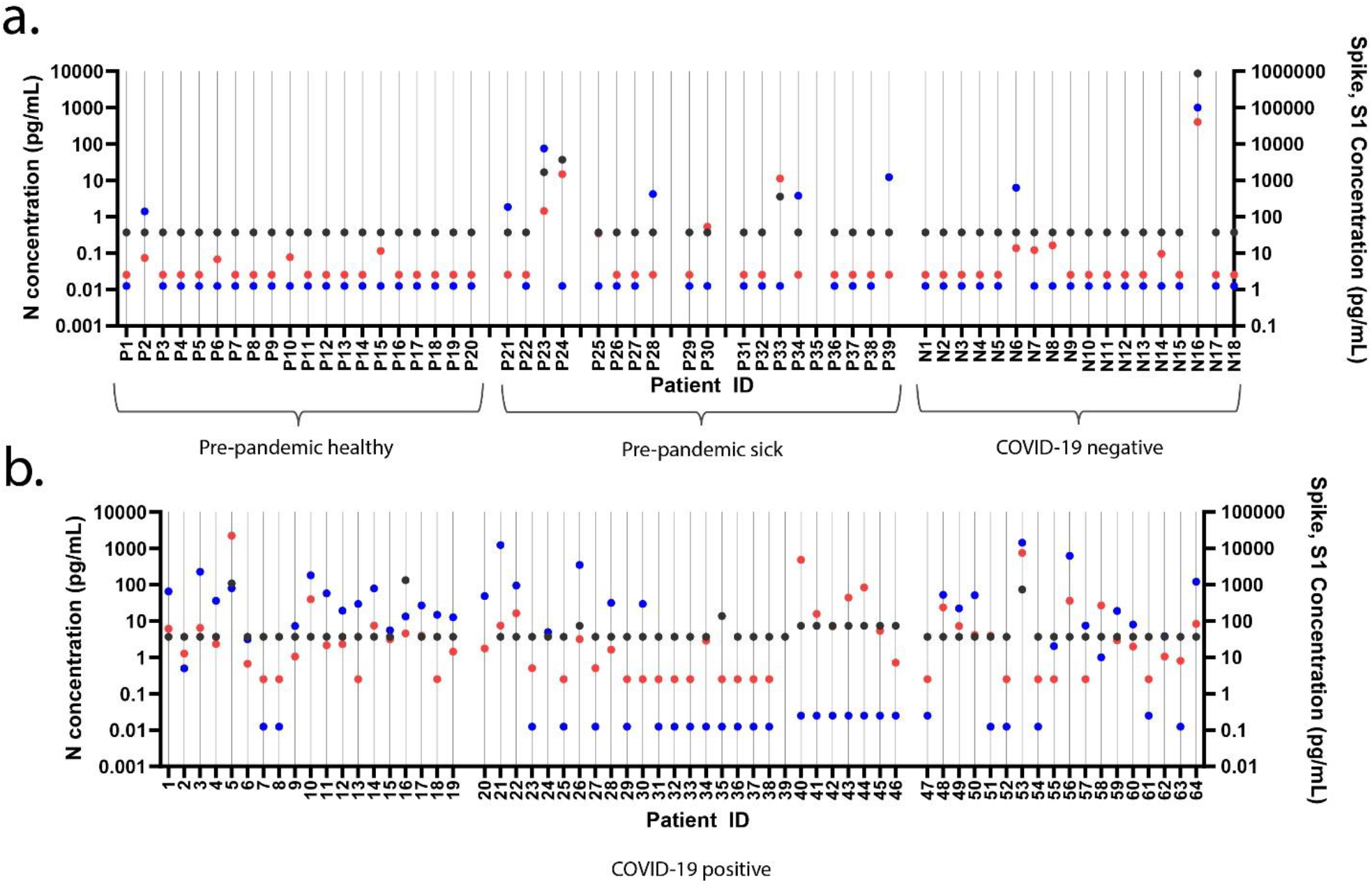
Viral antigen concentrations for each patient. a.) S1 (red), spike (black), and nucleocapsid (blue)concentrations plotted per patient from three control cohorts: pre-pandemic healthy, pre-pandemic sick, and patients who tested COVID-19 negative by NP RT-PCR. b.) S1 (red), spike (black), and nucleocapsid (blue)concentrations plotted per patient from patients who tested COVID-19 positive by NP RT-PCR.

**Figure S6.**
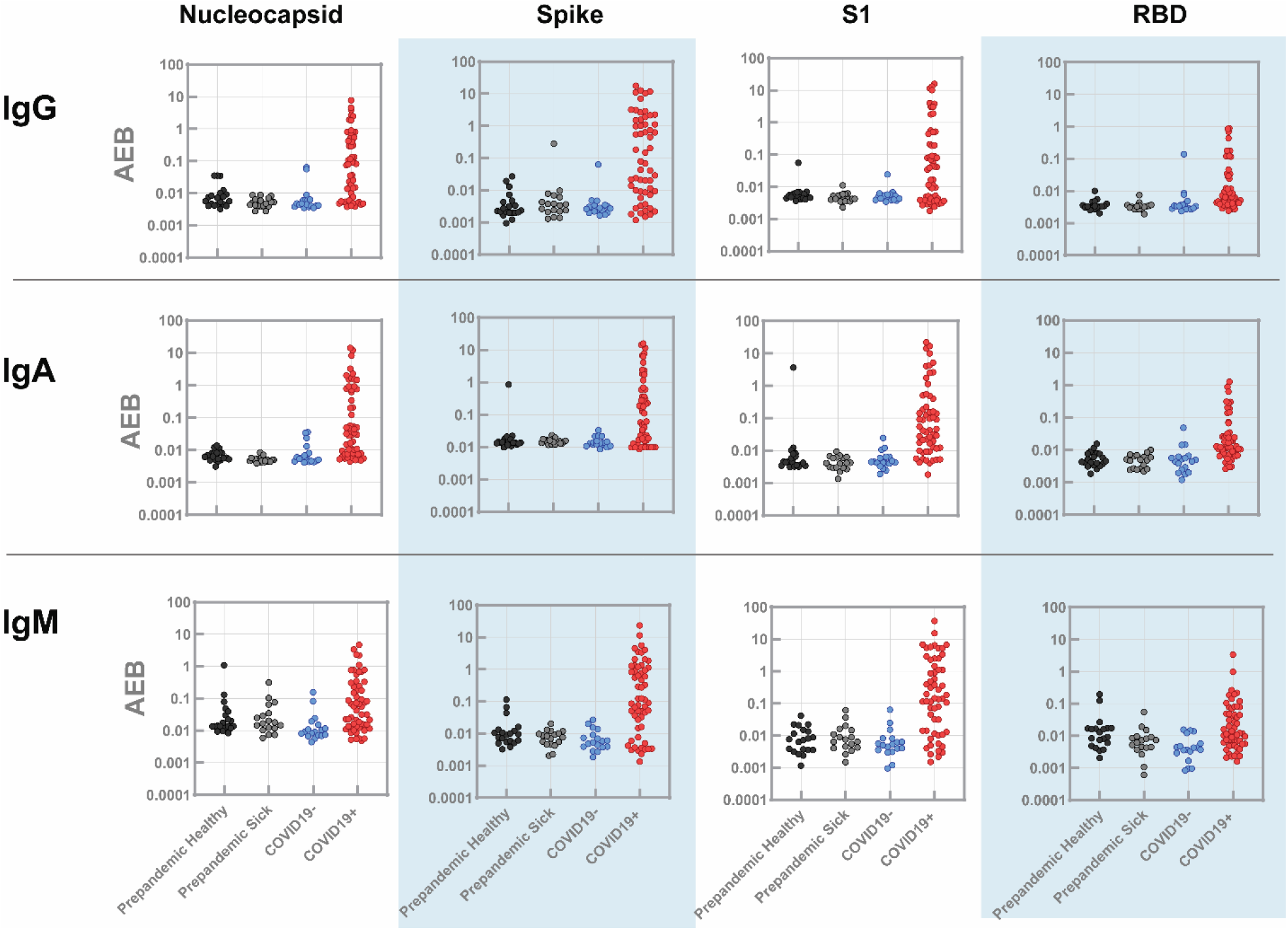
Simoa results for IgG, IgM, and IgG against four viral antigens: for samples collected from pre-pandemic healthy patients, pre-pandemic sick patients, COVID-19 negative patients, and COVID-19 positive patients. Each marker is the average AEB (average enzyme per bead) value from two replicate measurements.

**Figure S7.**
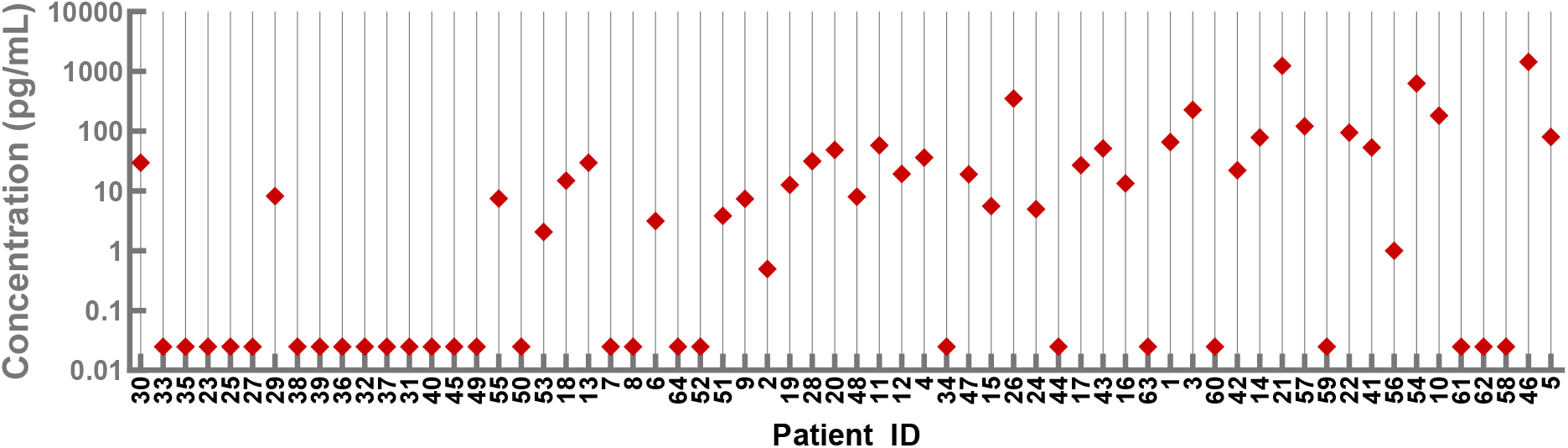
Nucleocapsid concentrations plotted by individual COVID-19 positive patients.

**Figure S8.**
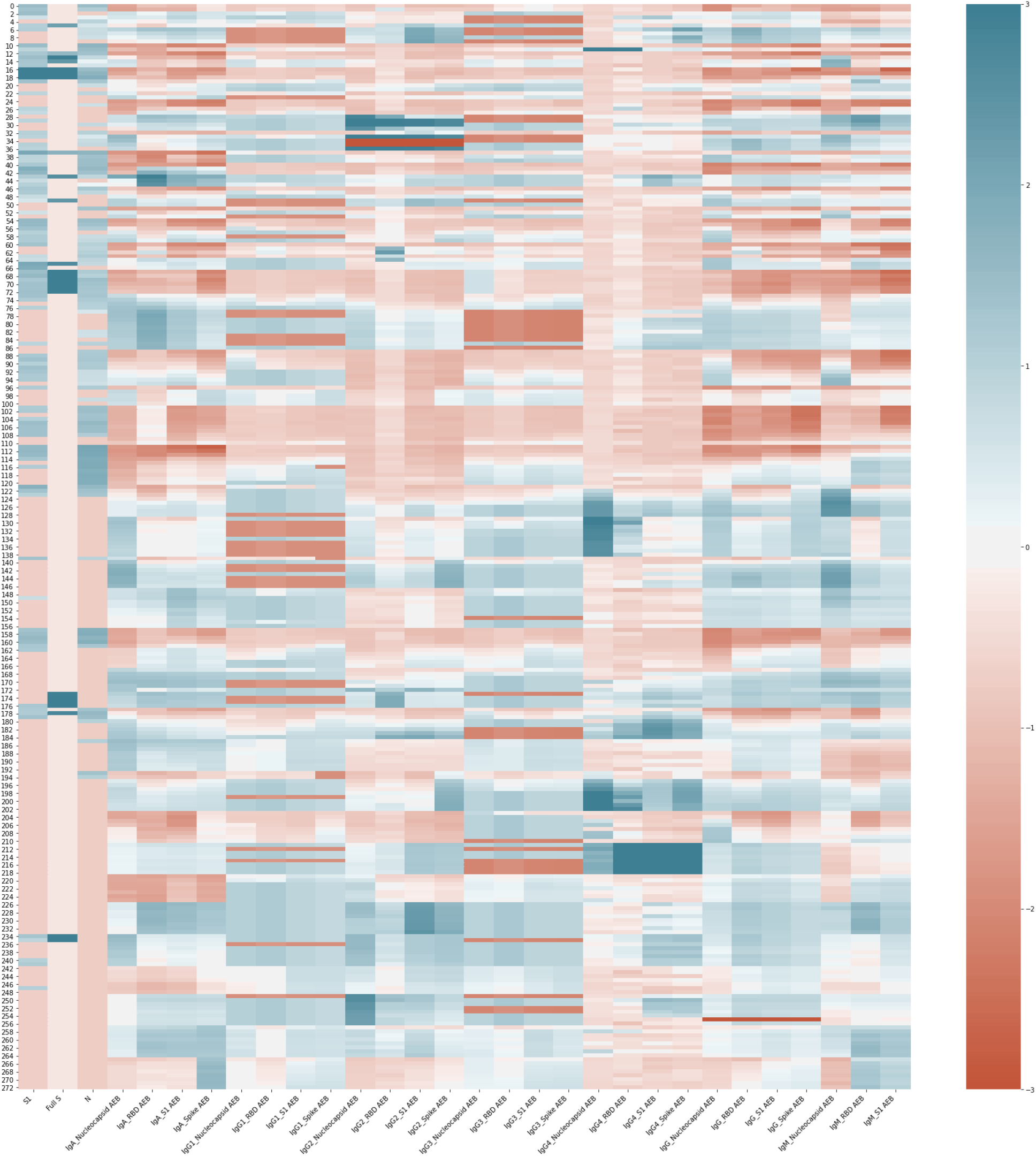
Heat map of SARS-CoV-2 antigen and antibody results for 39 patients (total samples N = 252). Map of standardized but un-clustered data. Scale bar represents the Standardized Biomarker Abundances from values −3 to 3. 31 biomarkers are listed along the x-axis of the heat map. Sample IDs are listed on the y-axis of the heat map.

#### Antigen and Immunoglobulin Data for All Patient Samples

Four plots are shown for each patient: (1) the total IgA, IgM, and IgG against four viral antigens (nucleocapsid, RBD, S1, and spike), (2) nucleocapsid antigen levels with IgA, IgM, and IgG against nucleocapsid, (3) S1 antigen levels with IgA, IgM, and IgG against S1, and (4) S1 antigen levels with IgG1-4 against S1. Six patients had detectable levels of Spike in their plasma. For those patients, a fifth plot of Spike antigen levels with IgA, IgM, and IgG against Spike is also shown. Plots for Patients 21, 22, 26, and 39 can be found in the main text.

**Figure S9.**
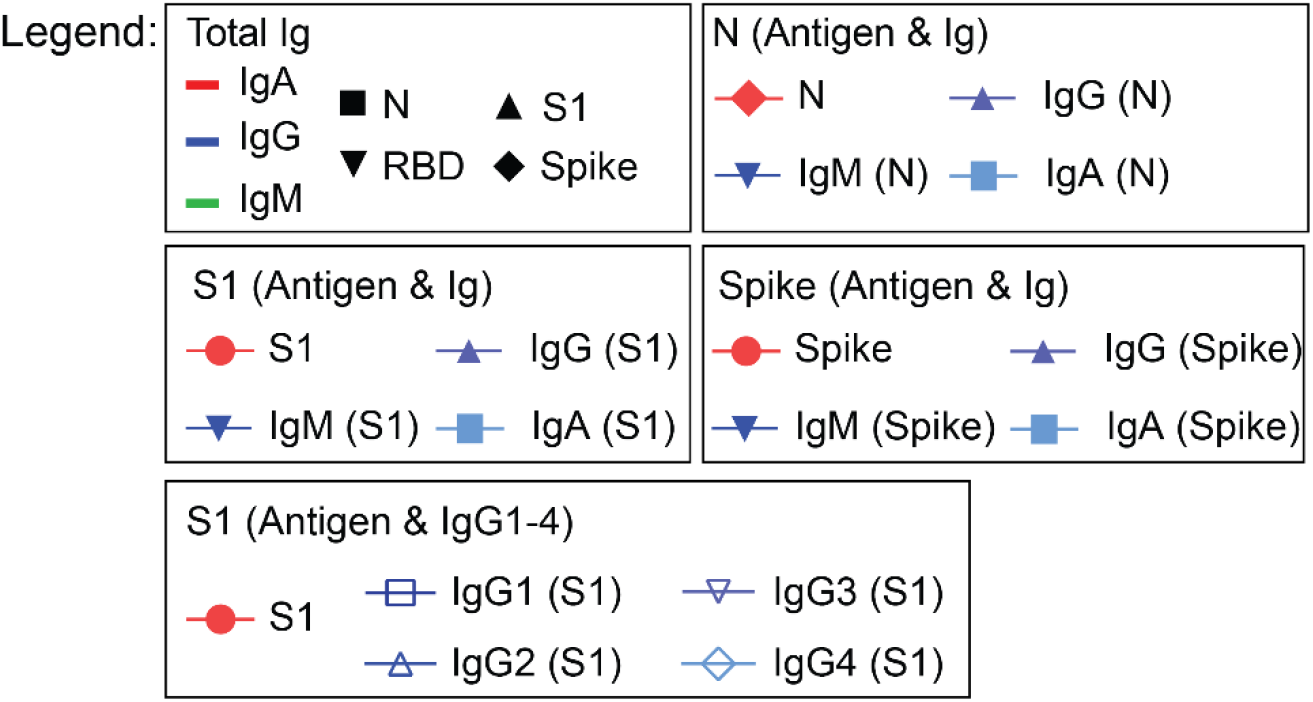
Plot Legend for viral antigen and immunoglobulin data for Patients 1-20, 23-25, and 27-38 in Figures S10 – S40.

**Figure S10.**
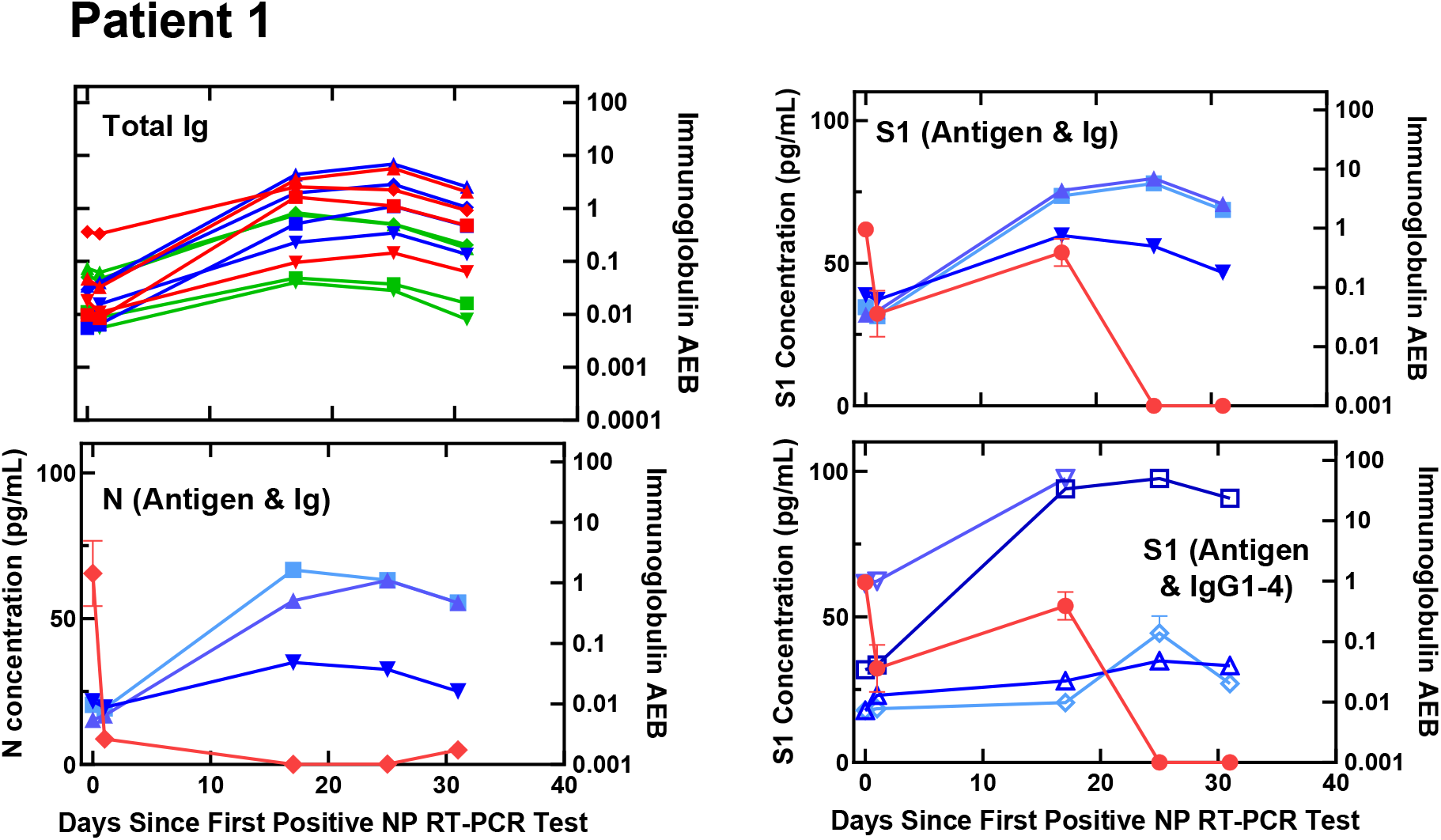
Viral antigen and anti-SARS-CoV-2 immunoglobulin plots for Patient 1.

**Figure S11.**
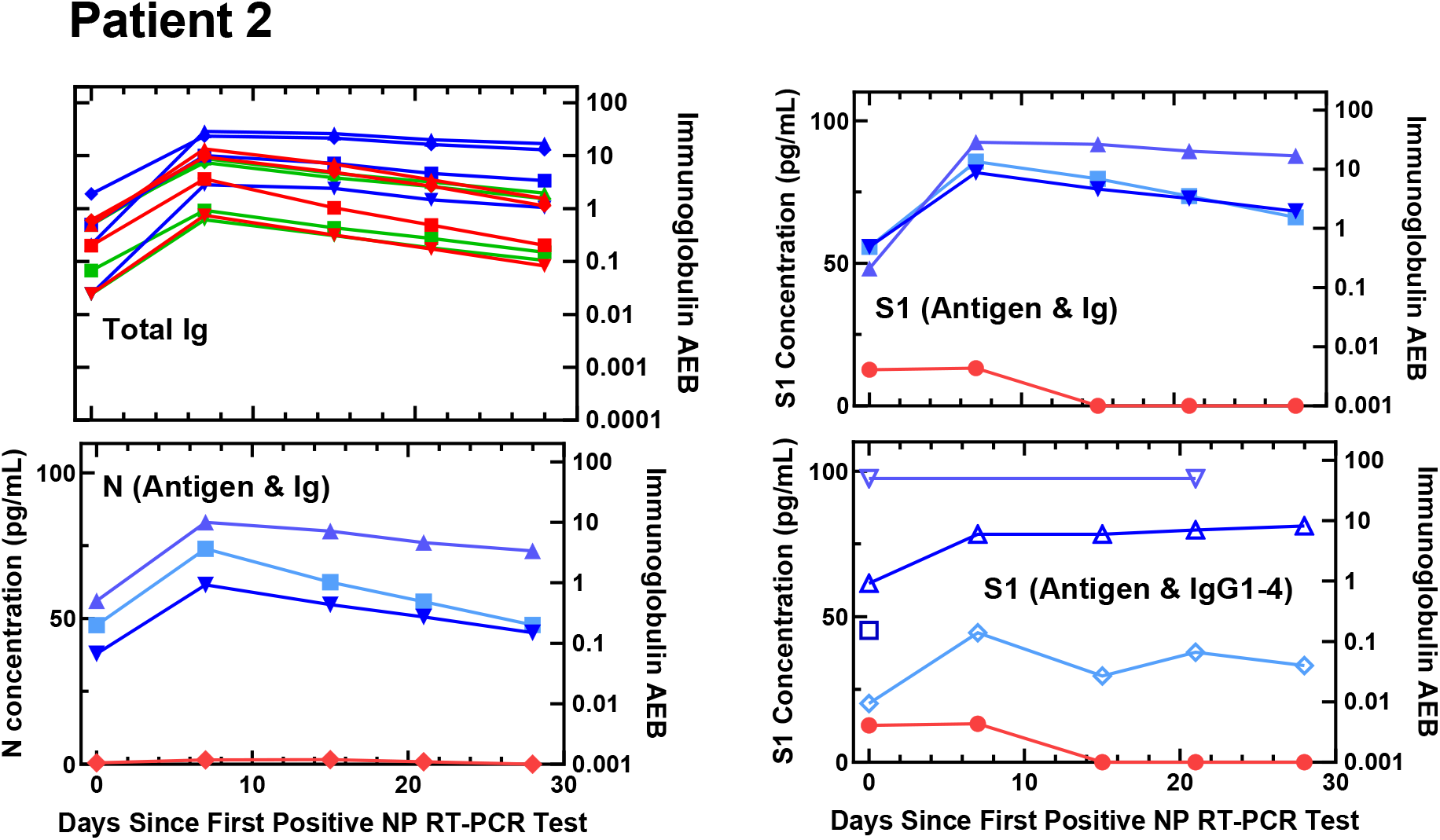
Viral antigen and anti-SARS-CoV-2 immunoglobulin plots for Patient 2.

**Figure S12.**
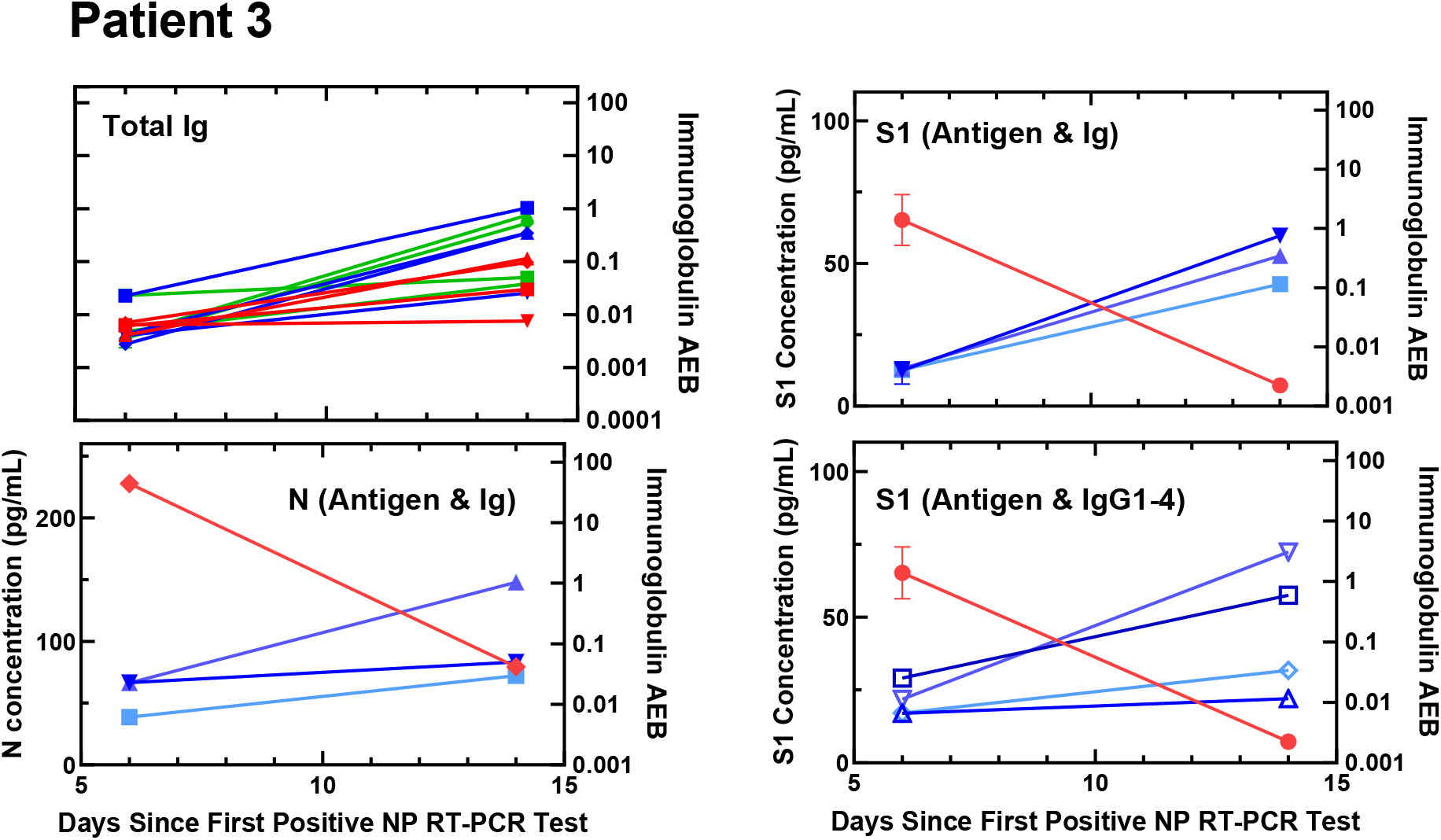
Viral antigen and anti-SARS-CoV-2 immunoglobulin plots for Patient 3.

**Figure S13.**
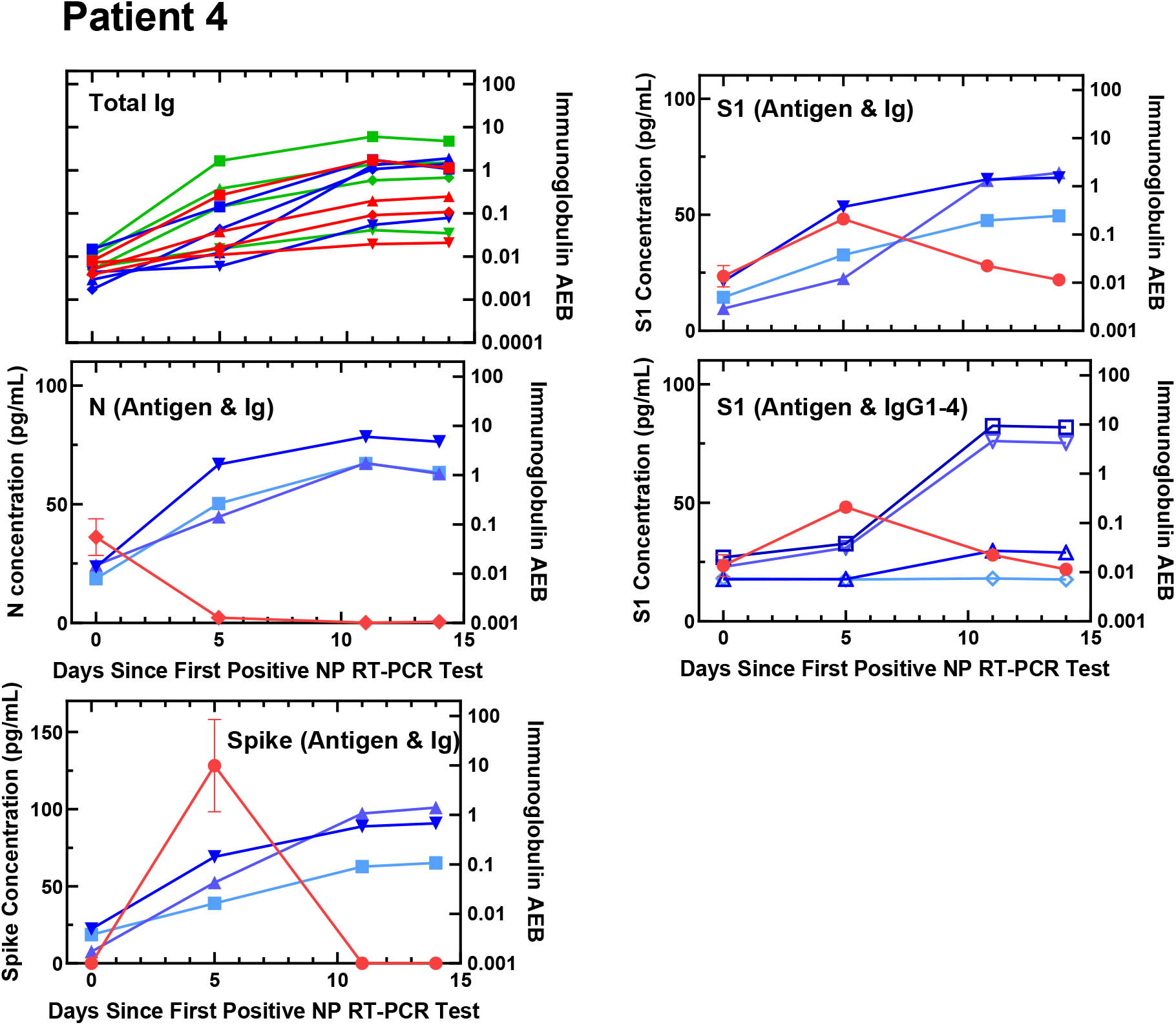
Viral antigen and anti-SARS-CoV-2 immunoglobulin plots for Patient 4.

**Figure S14.**
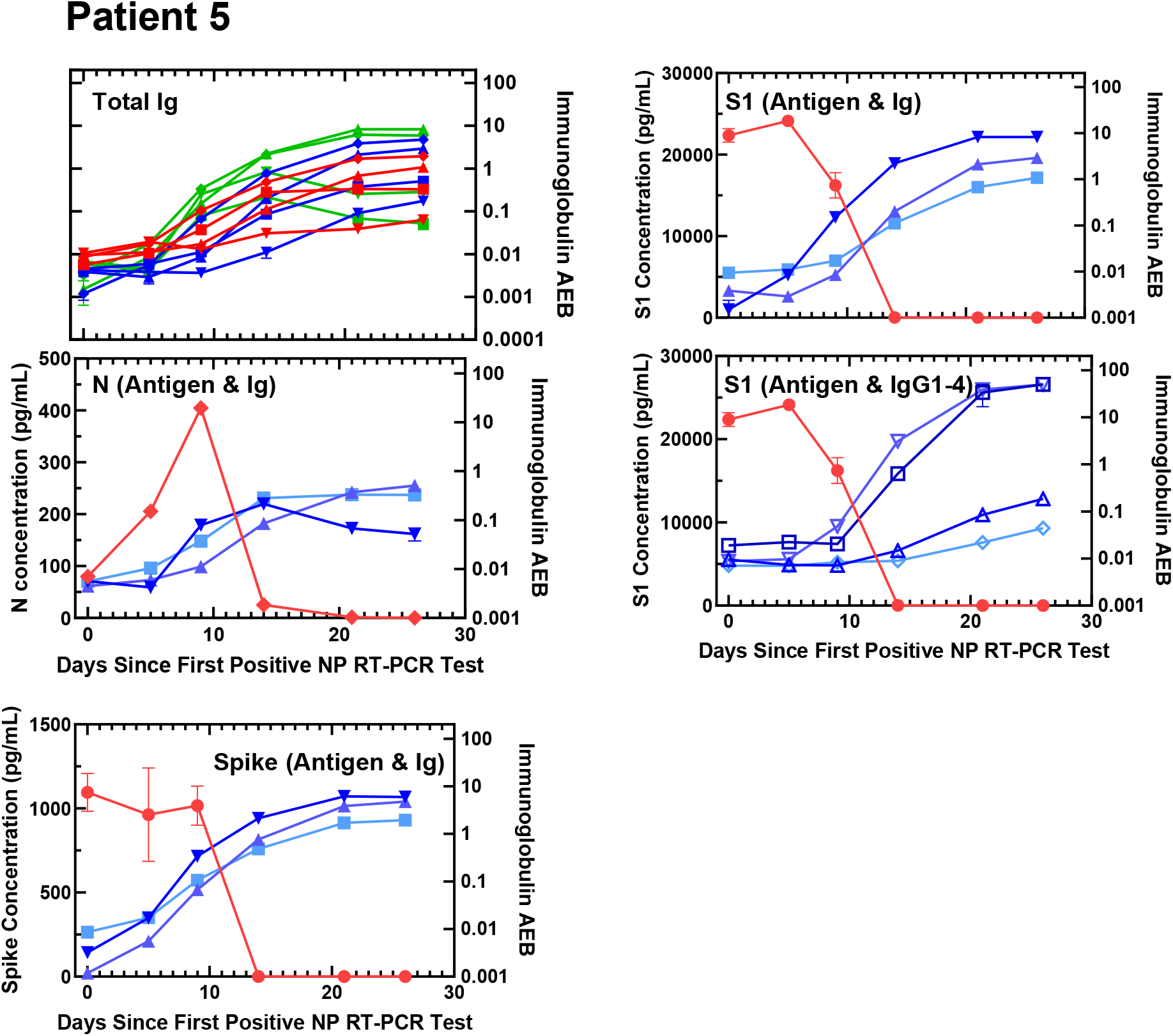
Viral antigen and anti-SARS-CoV-2 immunoglobulin plots for Patient 5.

**Figure S15.**
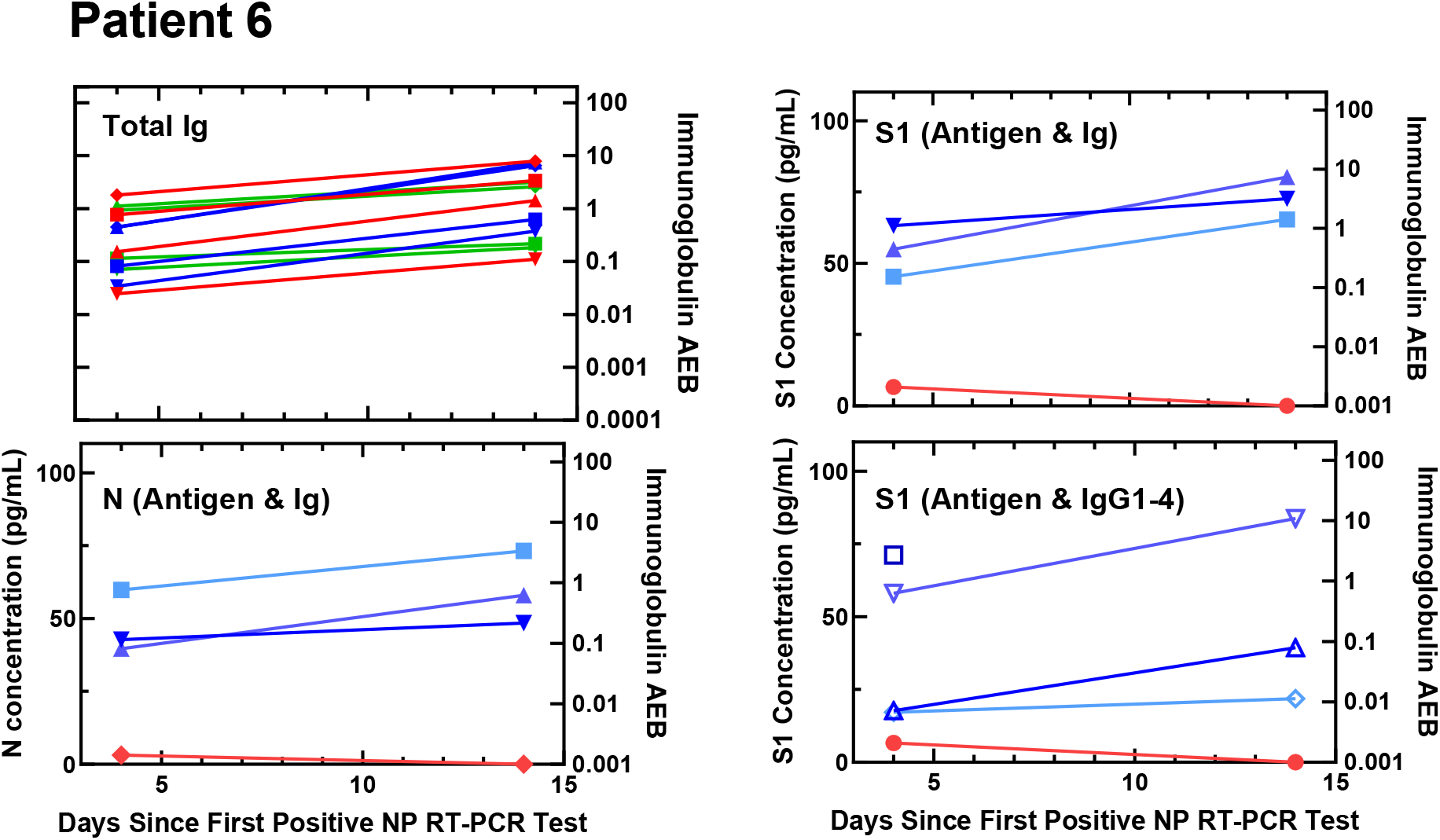
Viral antigen and anti-SARS-CoV-2 immunoglobulin plots for Patient 6.

**Figure S16.**
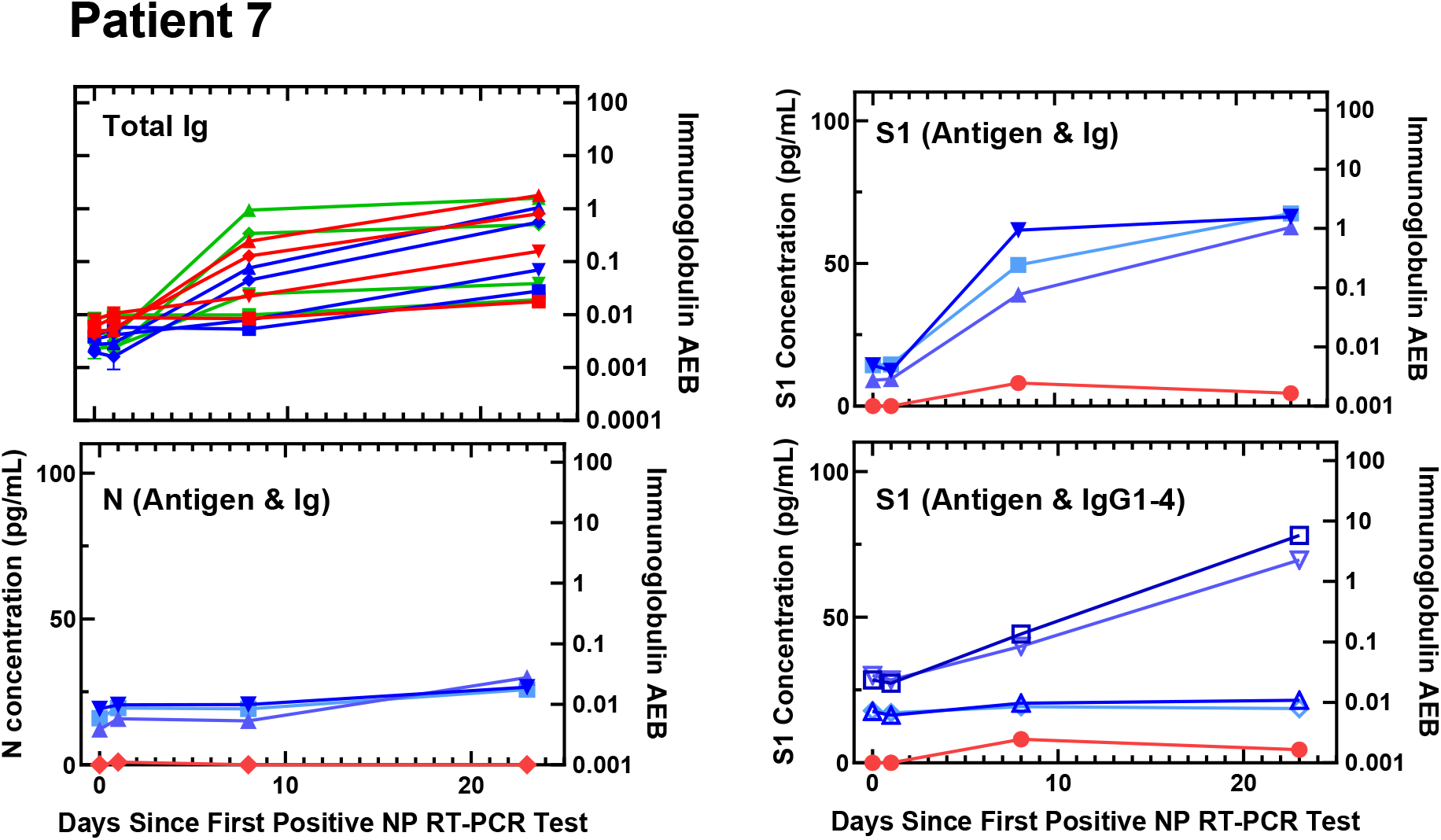
Viral antigen and anti-SARS-CoV-2 immunoglobulin plots for Patient 7.

**Figure S17.**
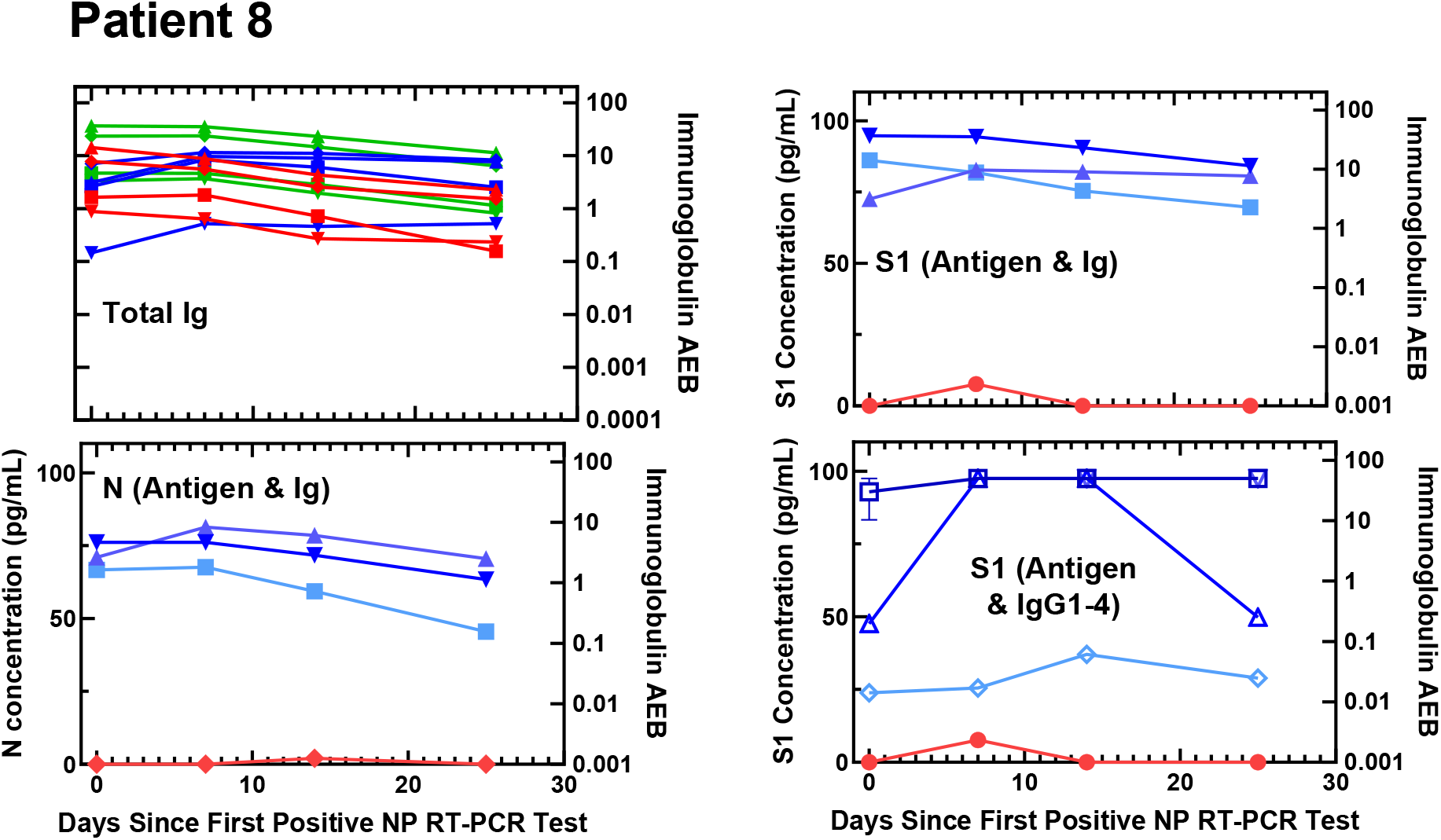
Viral antigen and anti-SARS-CoV-2 immunoglobulin plots for Patient 8.

**Figure S18.**
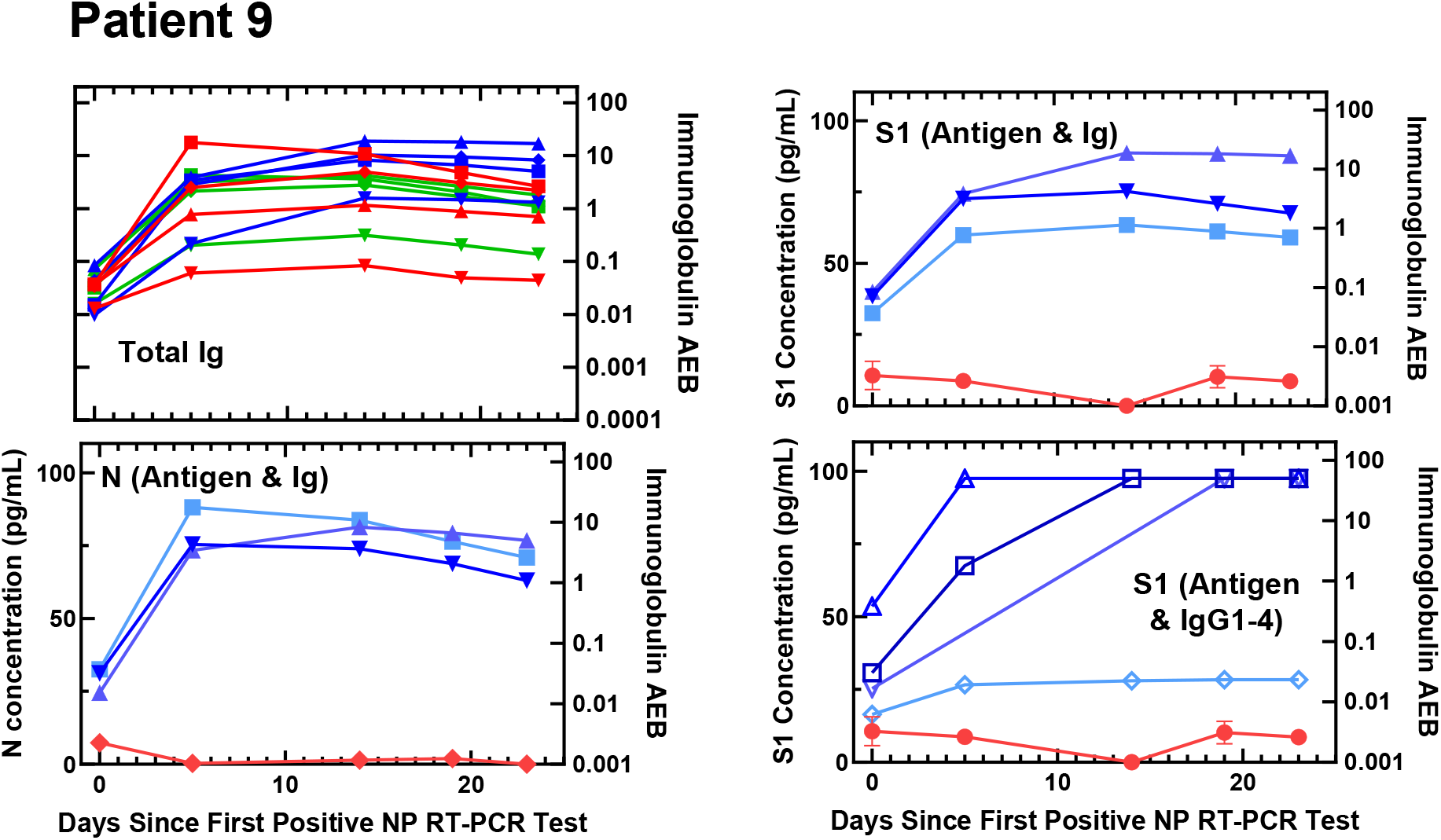
Viral antigen and anti-SARS-CoV-2 immunoglobulin plots for Patient 9.

**Figure S19.**
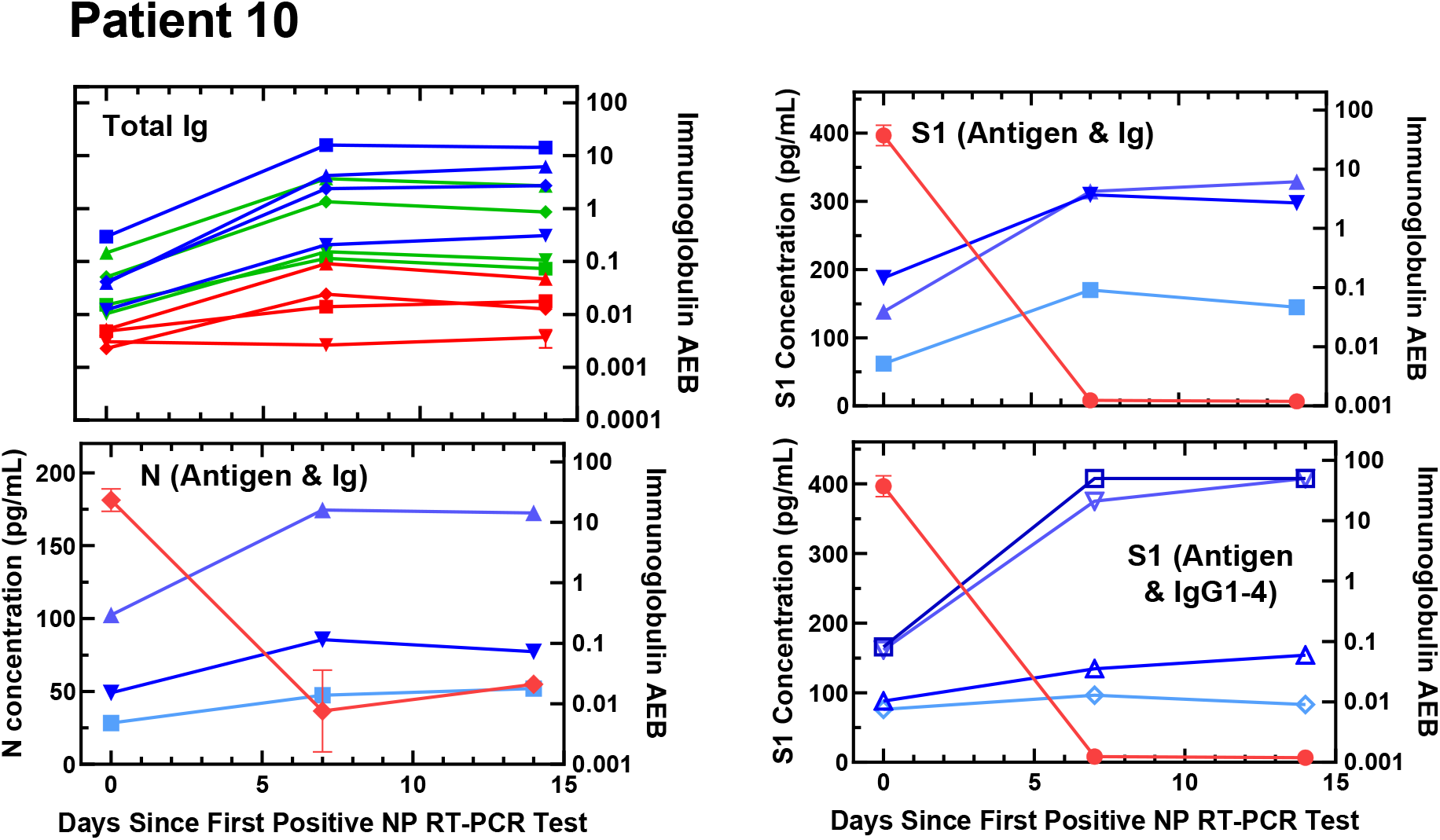
Viral antigen and anti-SARS-CoV-2 immunoglobulin plots for Patient 10.

**Figure S20.**
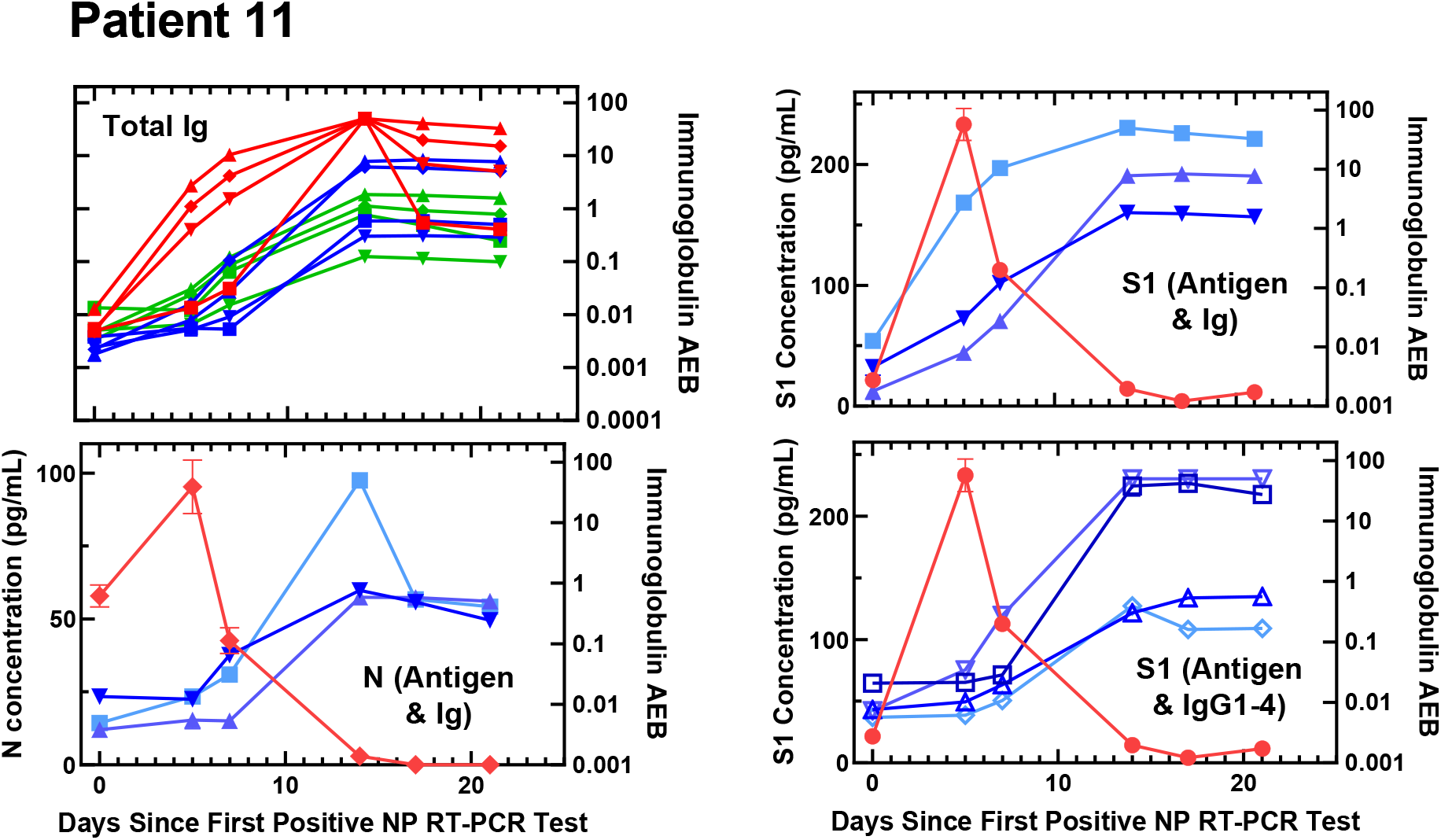
Viral antigen and anti-SARS-CoV-2 immunoglobulin plots for Patient 11.

**Figure S21.**
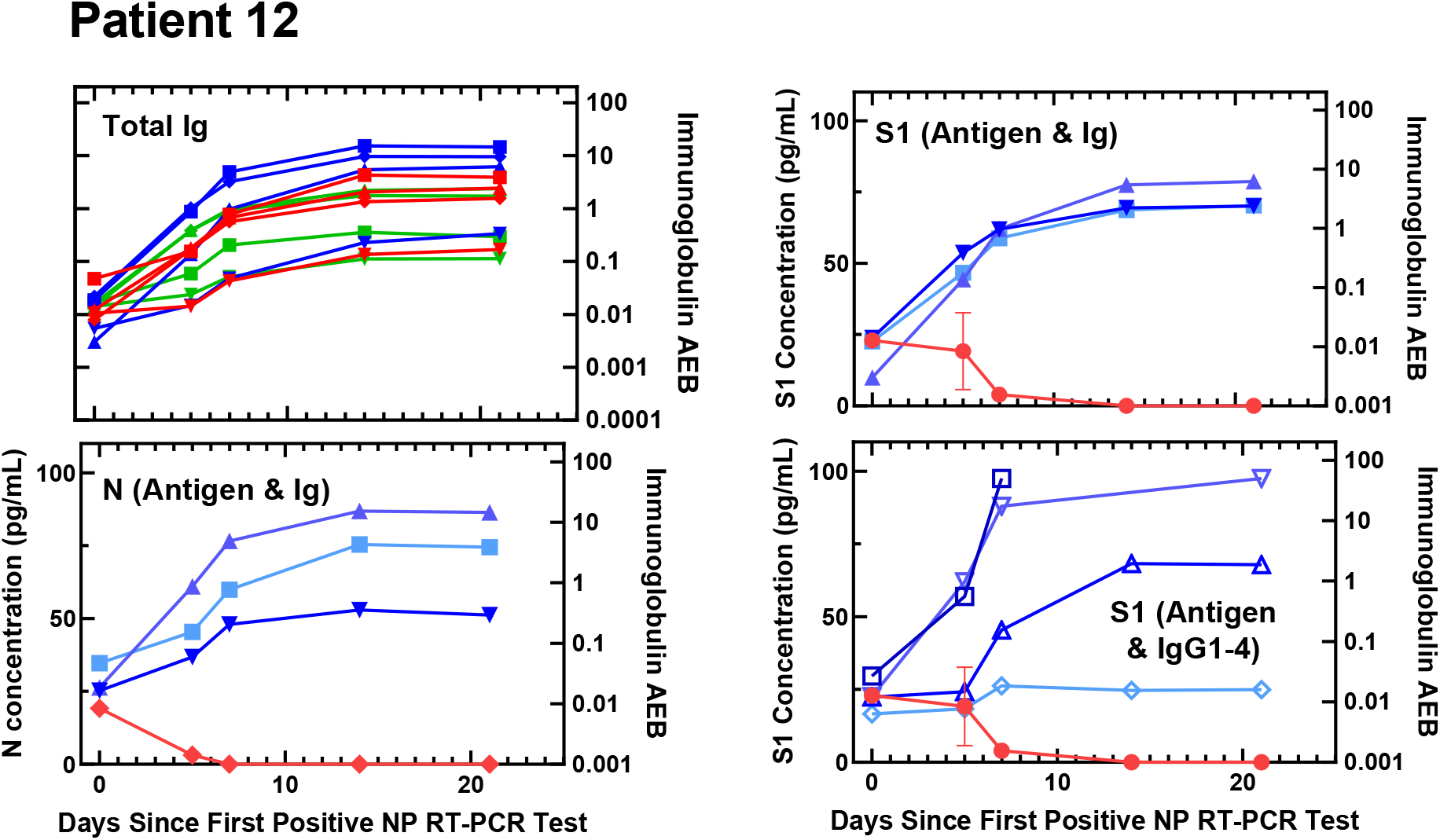
Viral antigen and anti-SARS-CoV-2 immunoglobulin plots for Patient 12.

**Figure S22.**
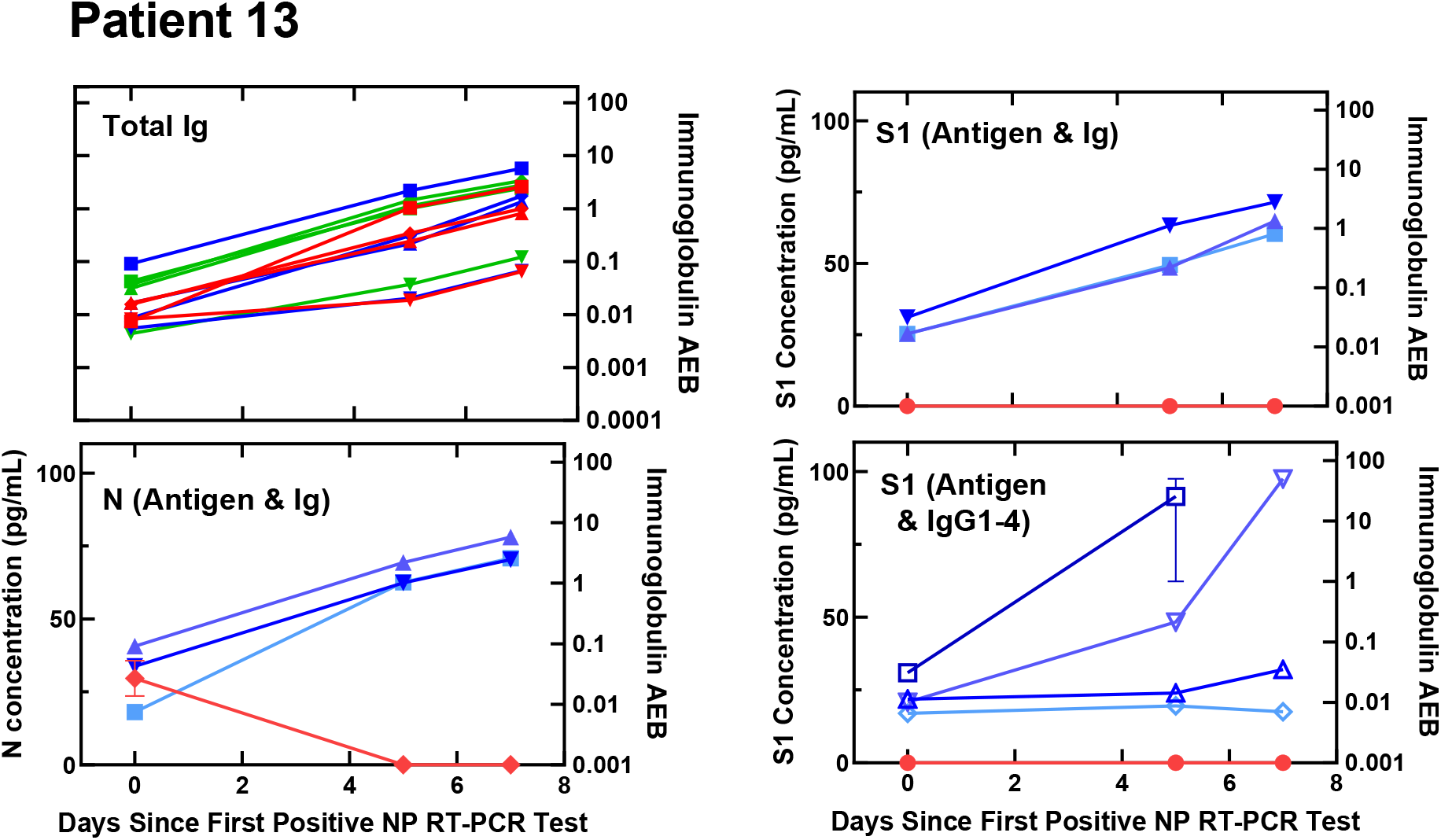
Viral antigen and anti-SARS-CoV-2 immunoglobulin plots for Patient 13.

**Figure S23.**
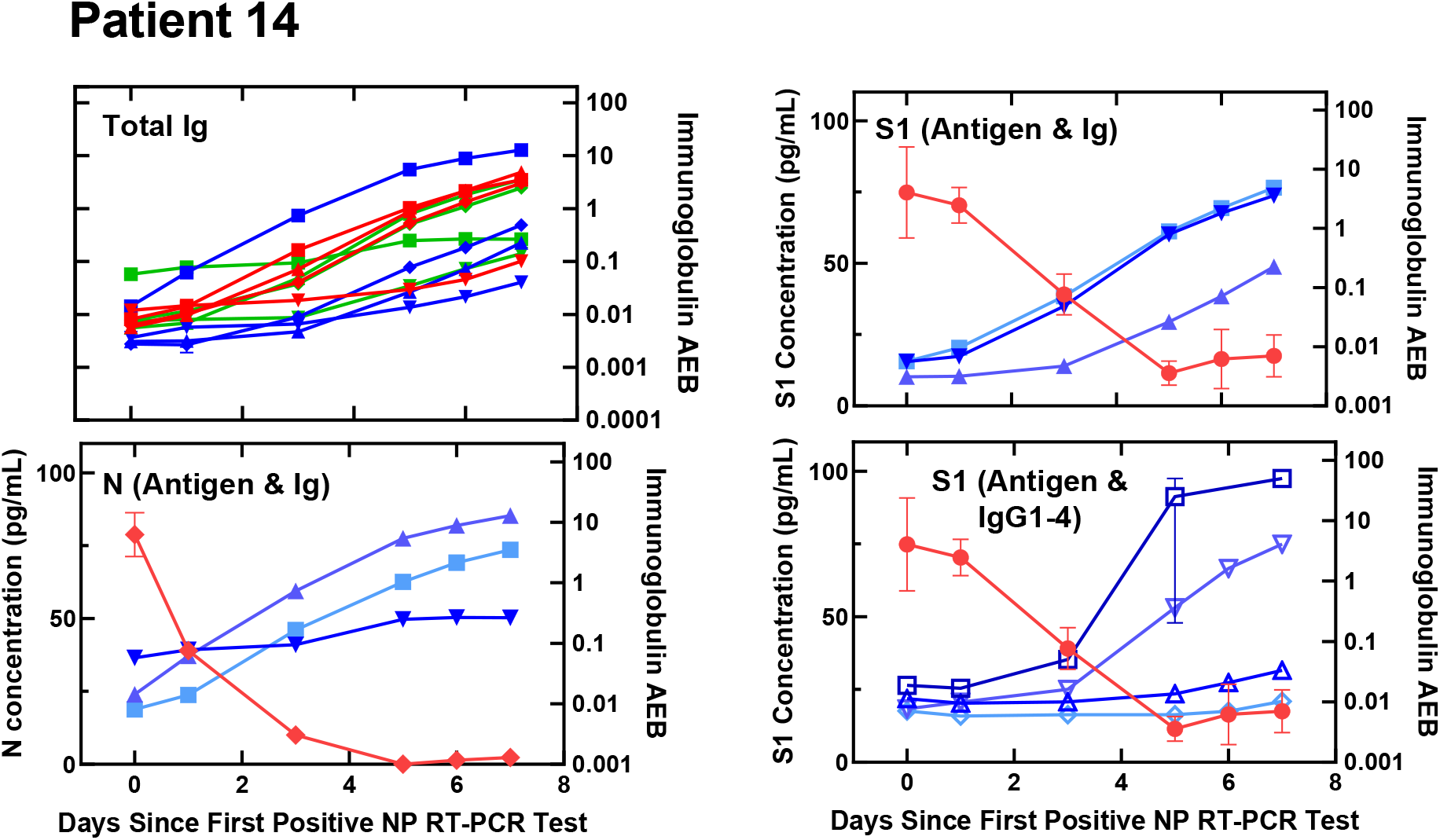
Viral antigen and anti-SARS-CoV-2 immunoglobulin plots for Patient 14.

**Figure S24.**
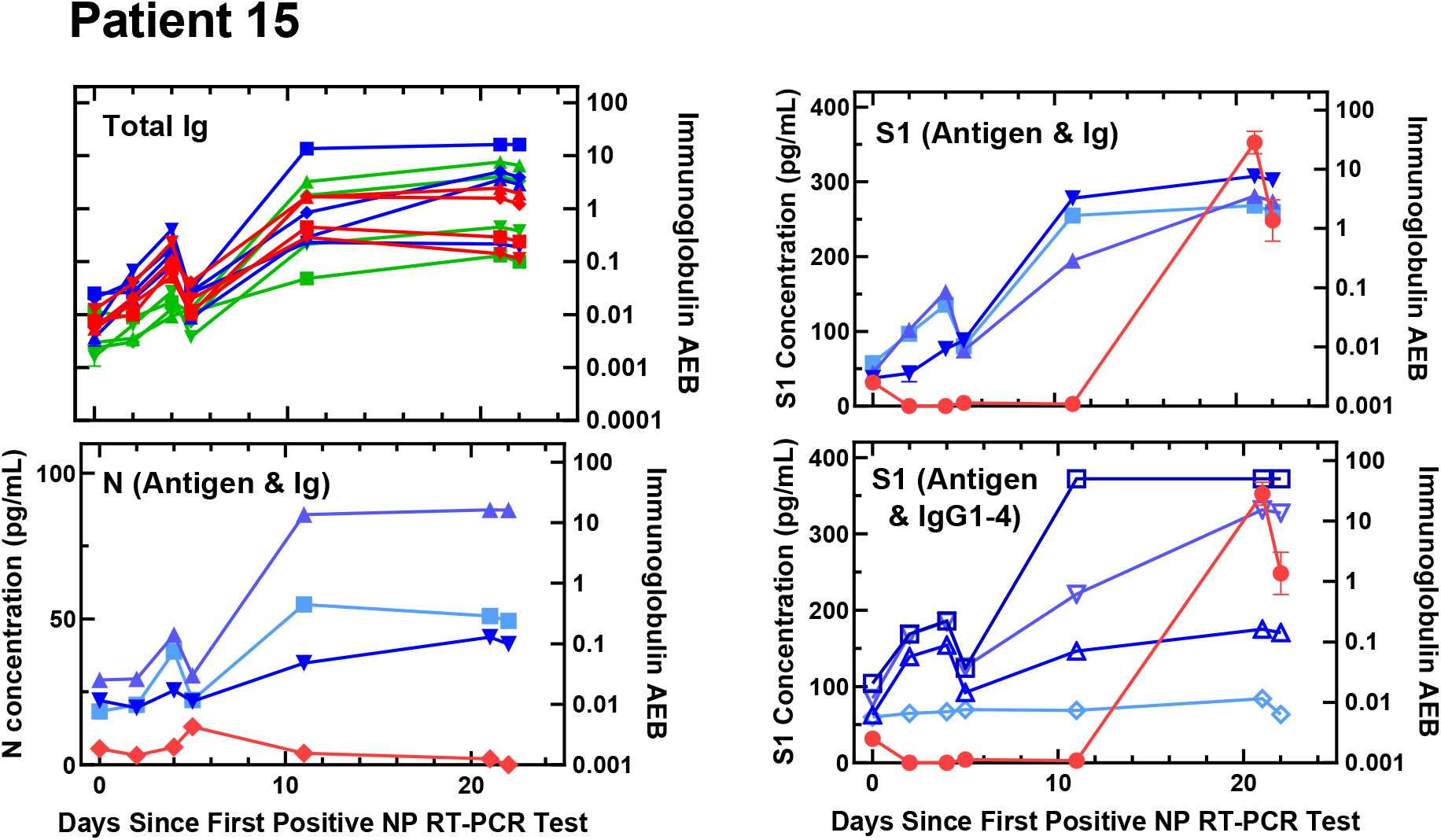
Viral antigen and anti-SARS-CoV-2 immunoglobulin plots for Patient 15.

**Figure S25.**
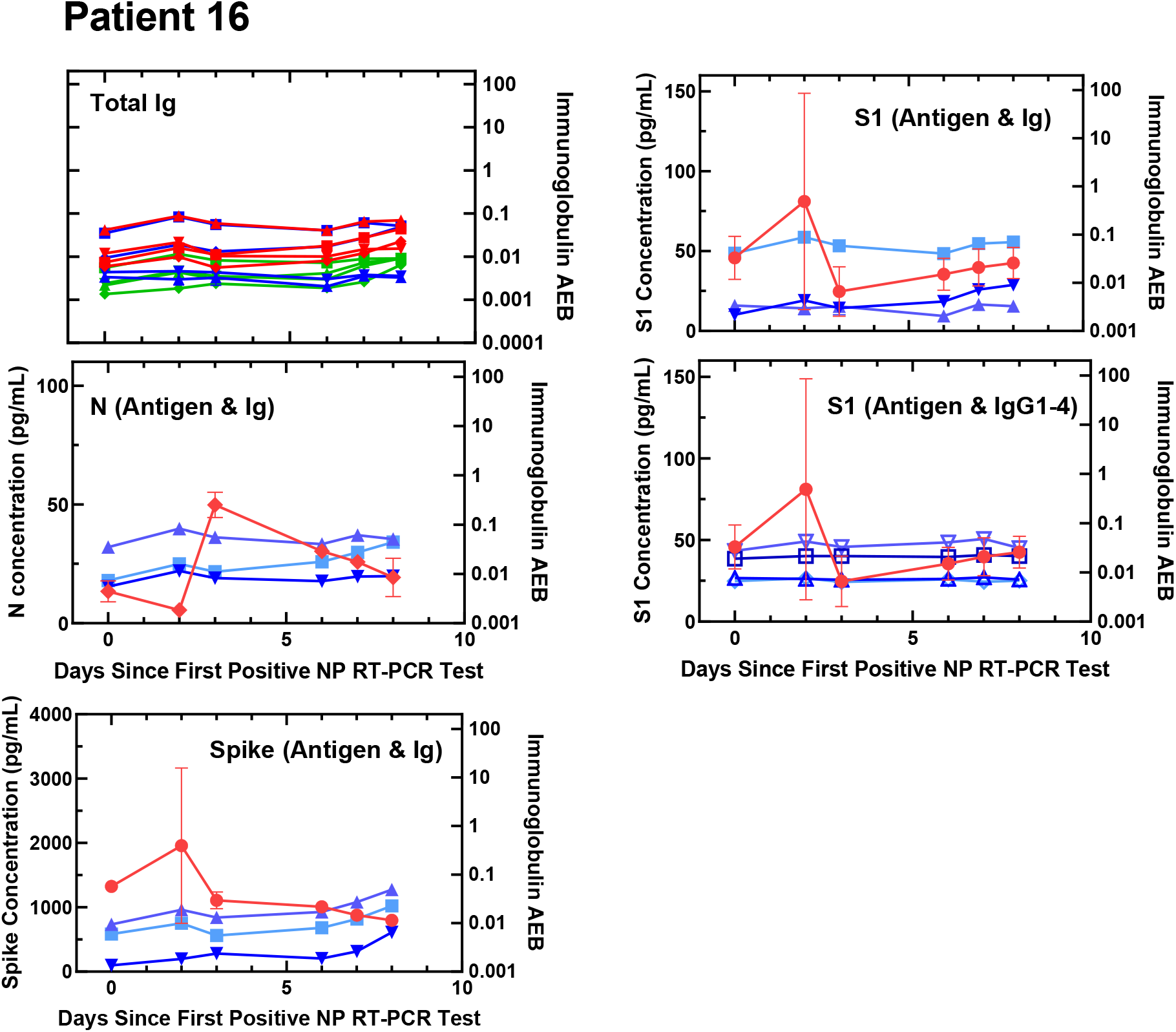
Viral antigen and anti-SARS-CoV-2 immunoglobulin plots for Patient 16.

**Figure S26.**
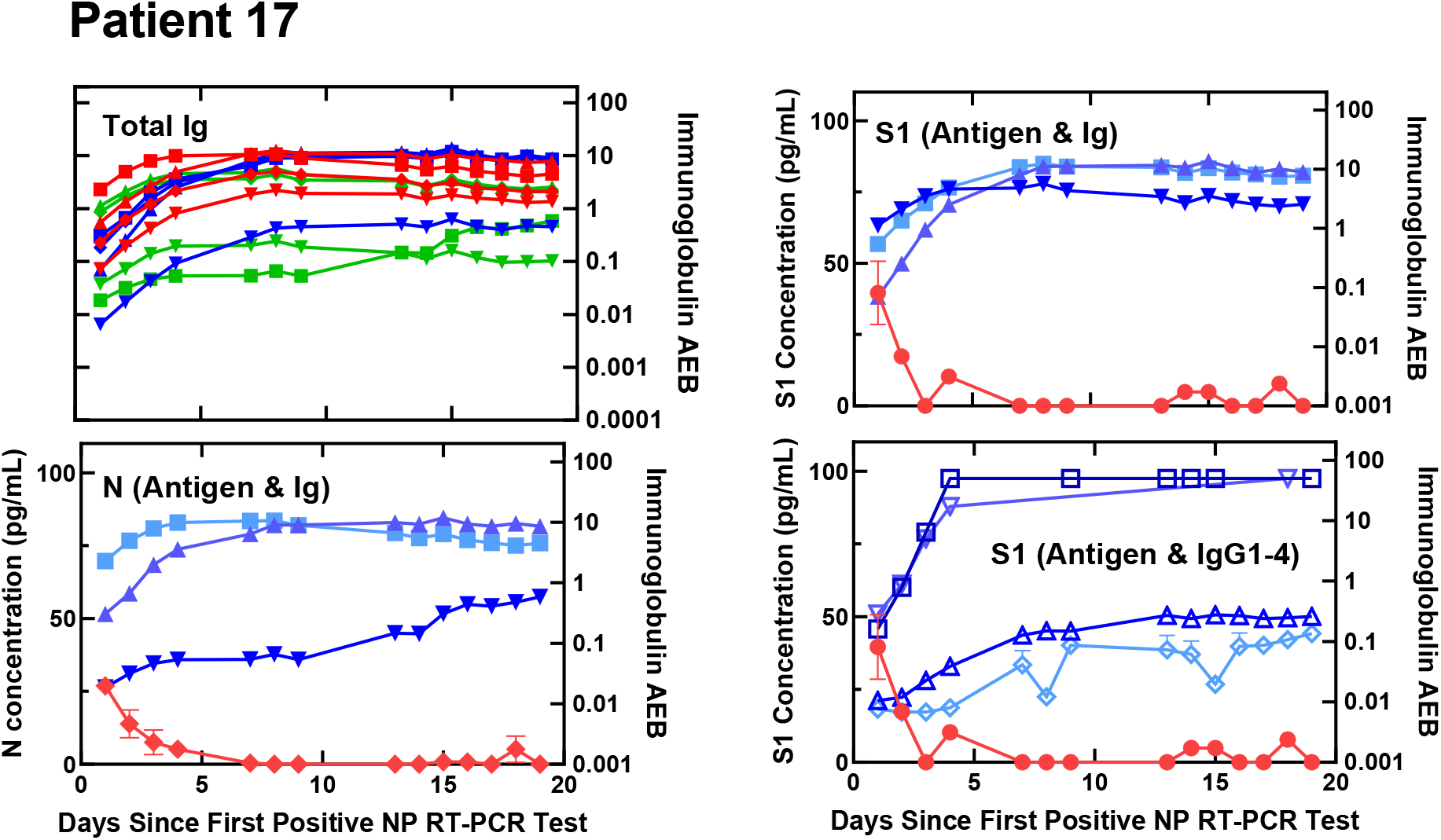
Viral antigen and anti-SARS-CoV-2 immunoglobulin plots for Patient 17.

**Figure S27.**
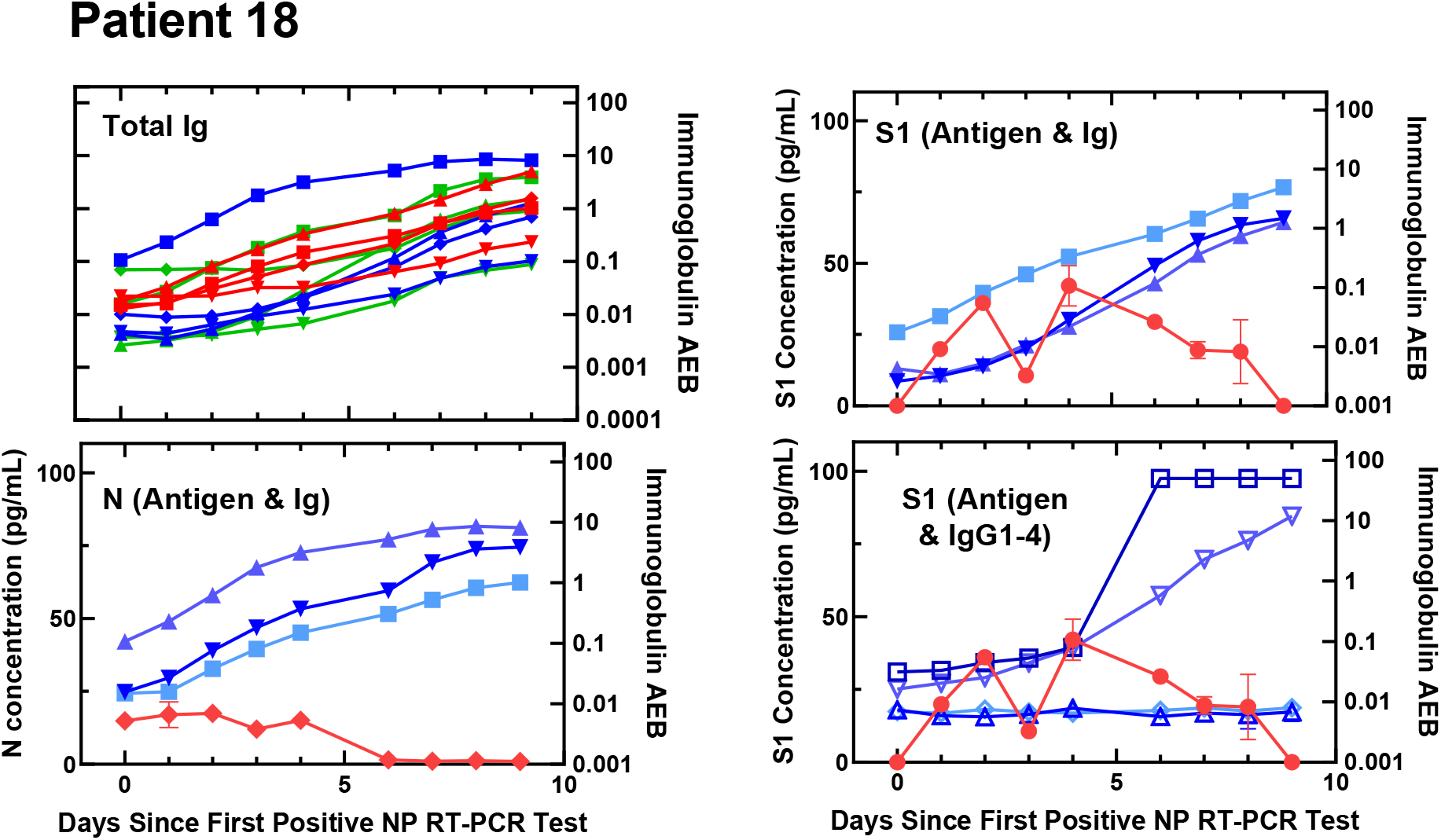
Viral antigen and anti-SARS-CoV-2 immunoglobulin plots for Patient 18.

**Figure S28.**
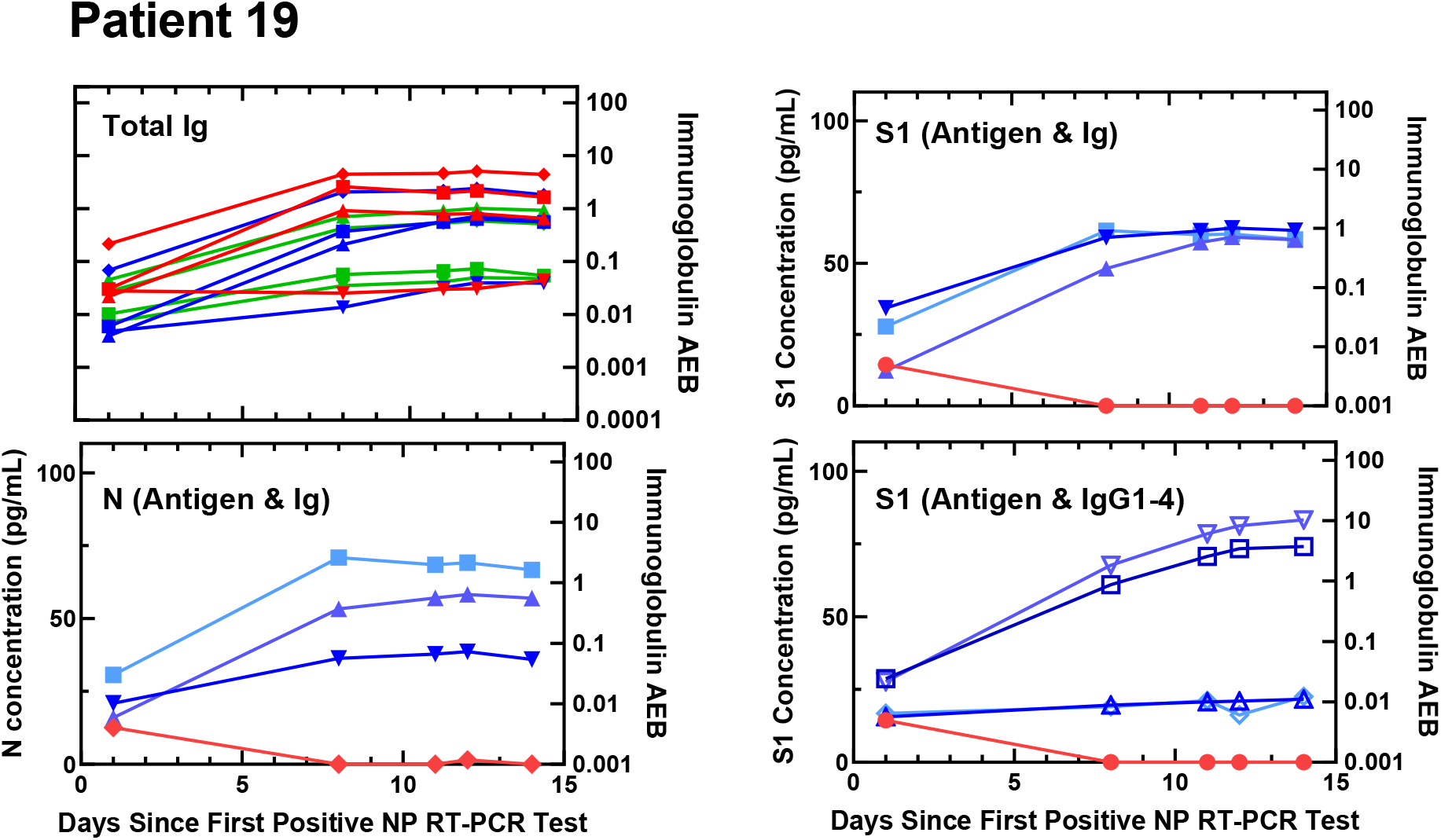
Viral antigen and anti-SARS-CoV-2 immunoglobulin plots for Patient 19.

**Figure S29.**
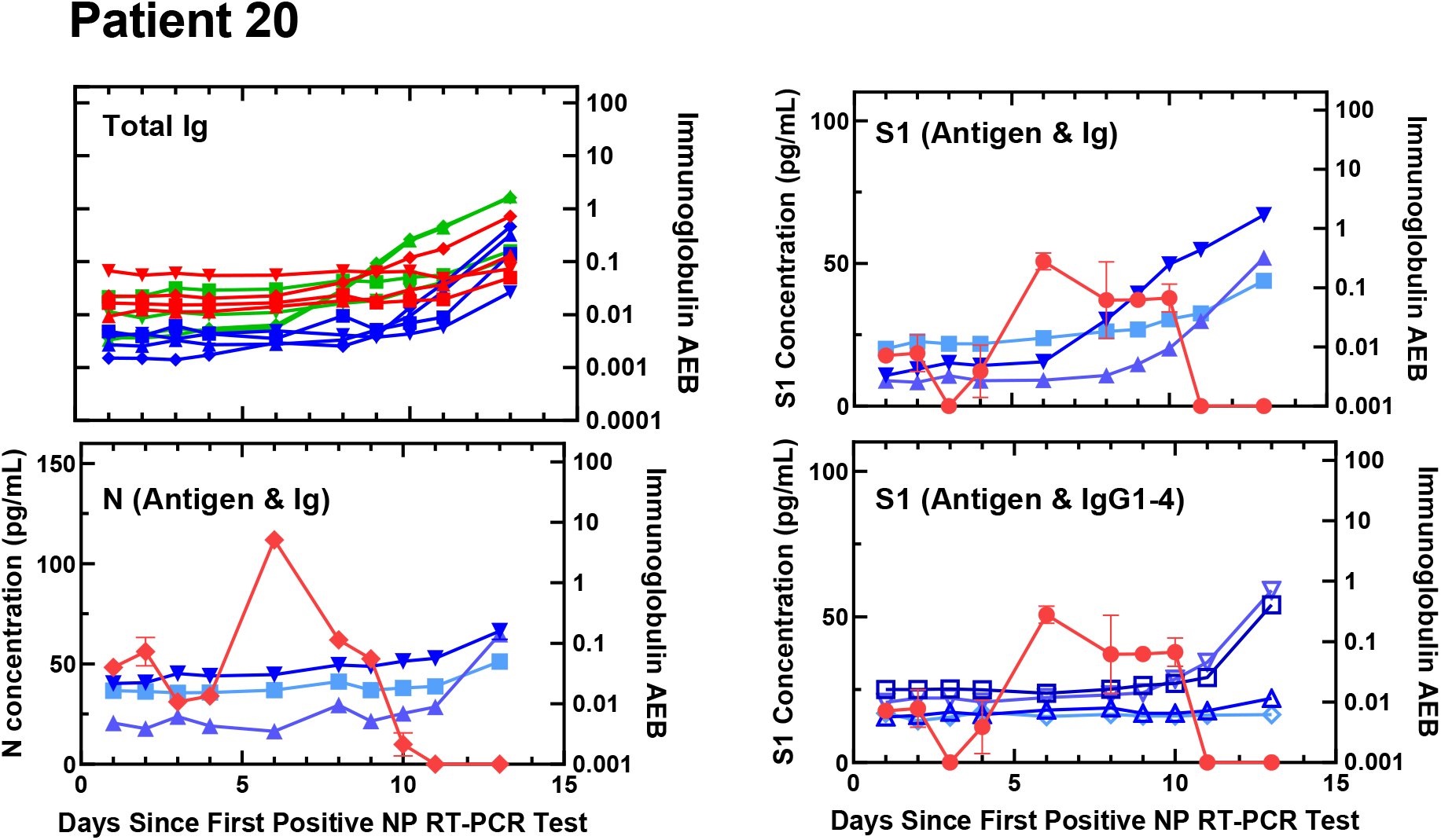
Viral antigen and anti-SARS-CoV-2 immunoglobulin plots for Patient 20.

**Figure S30.**
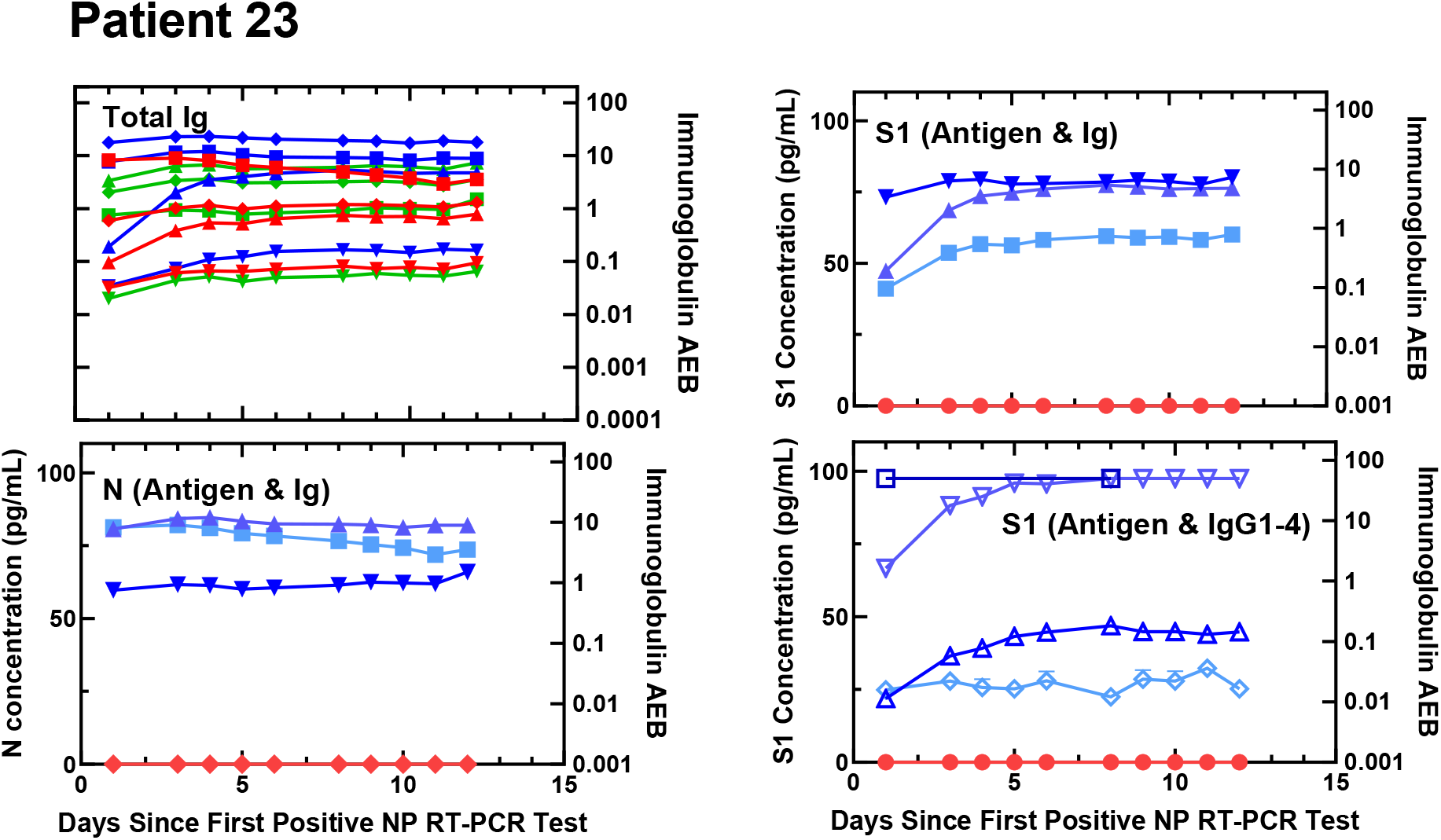
Viral antigen and anti-SARS-CoV-2 immunoglobulin plots for Patient 23.

**Figure S31.**
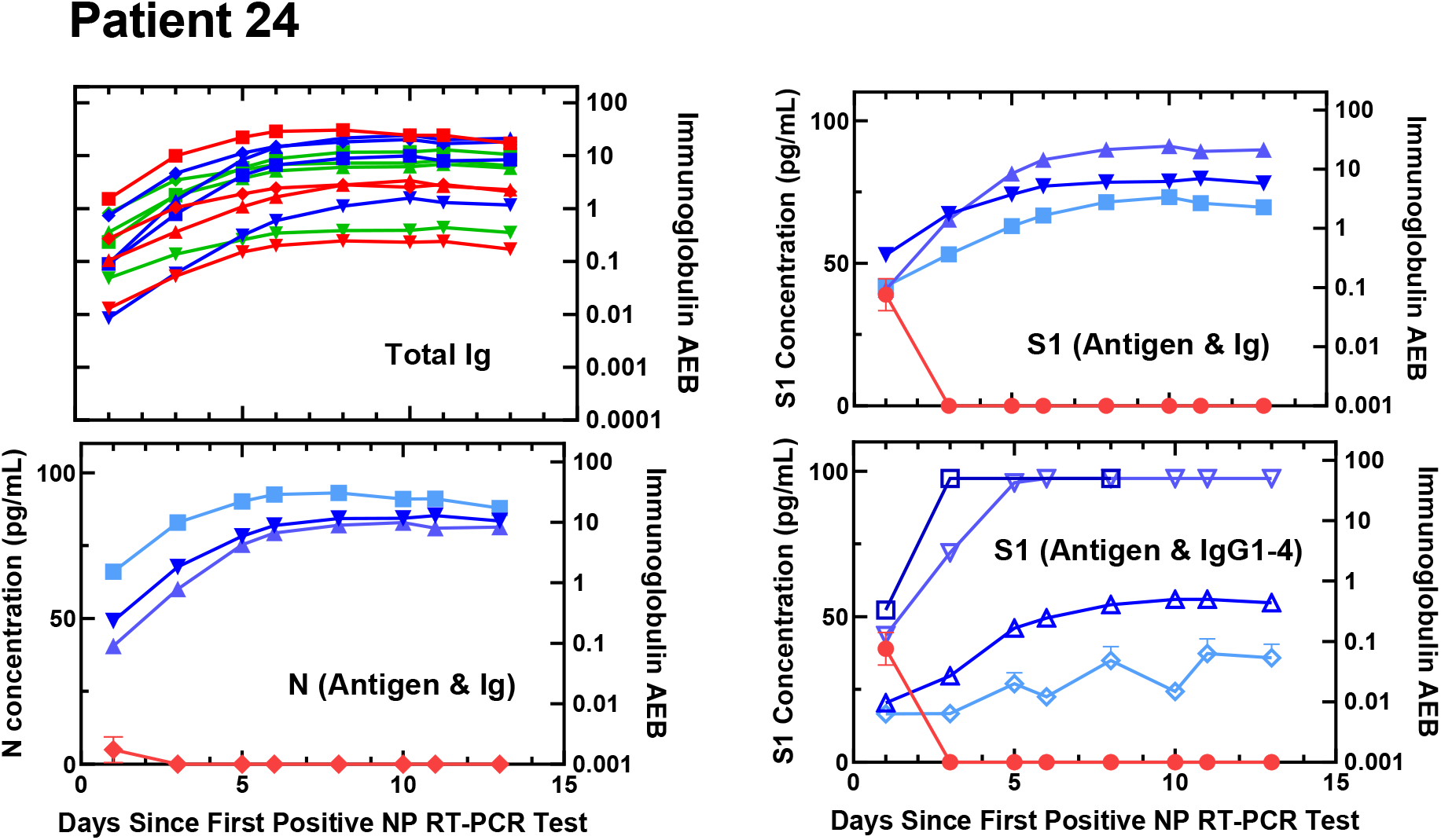
Viral antigen and anti-SARS-CoV-2 immunoglobulin plots for Patient 24.

**Figure S32.**
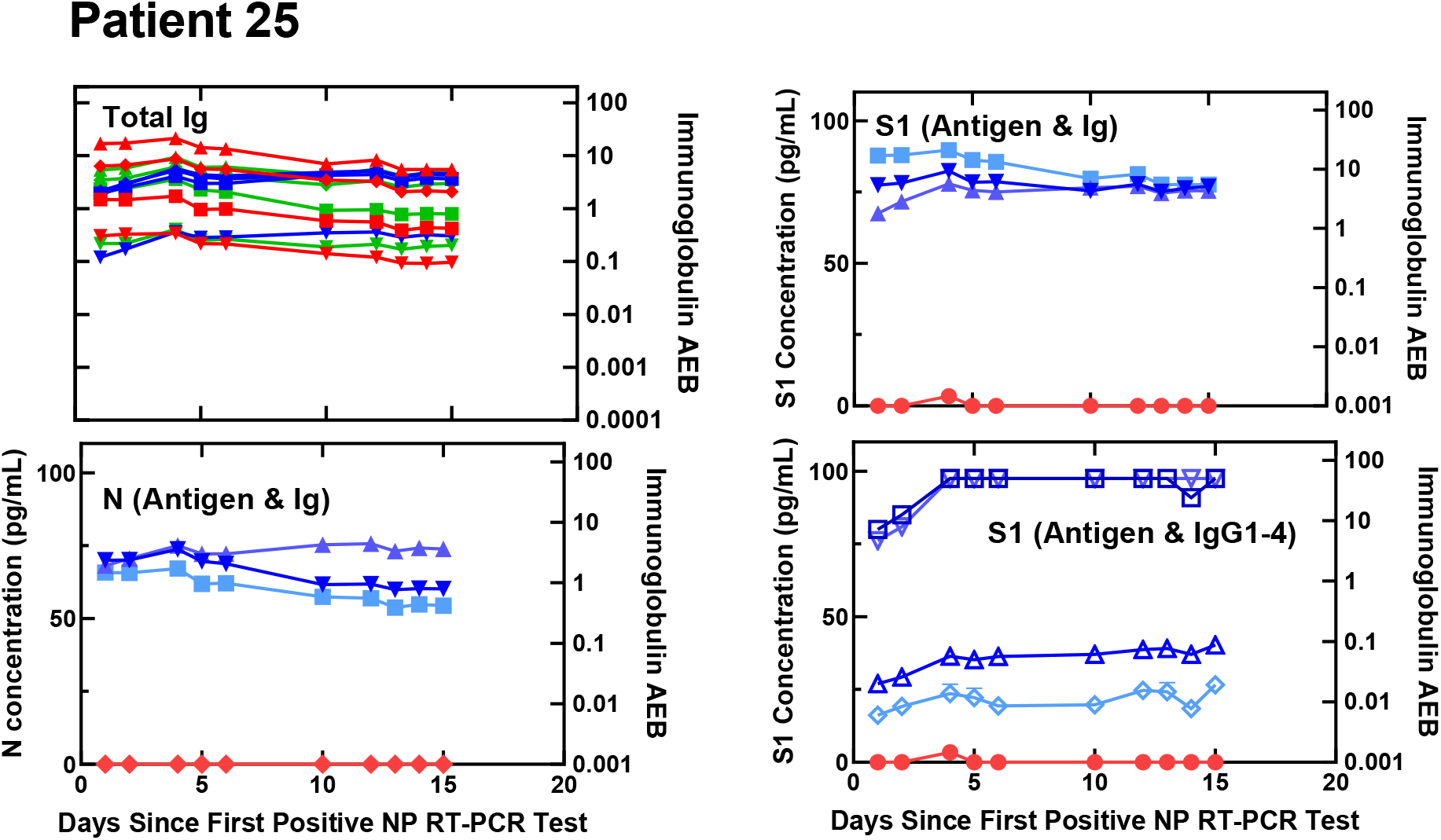
Viral antigen and anti-SARS-CoV-2 immunoglobulin plots for Patient 25.

**Figure S33.**
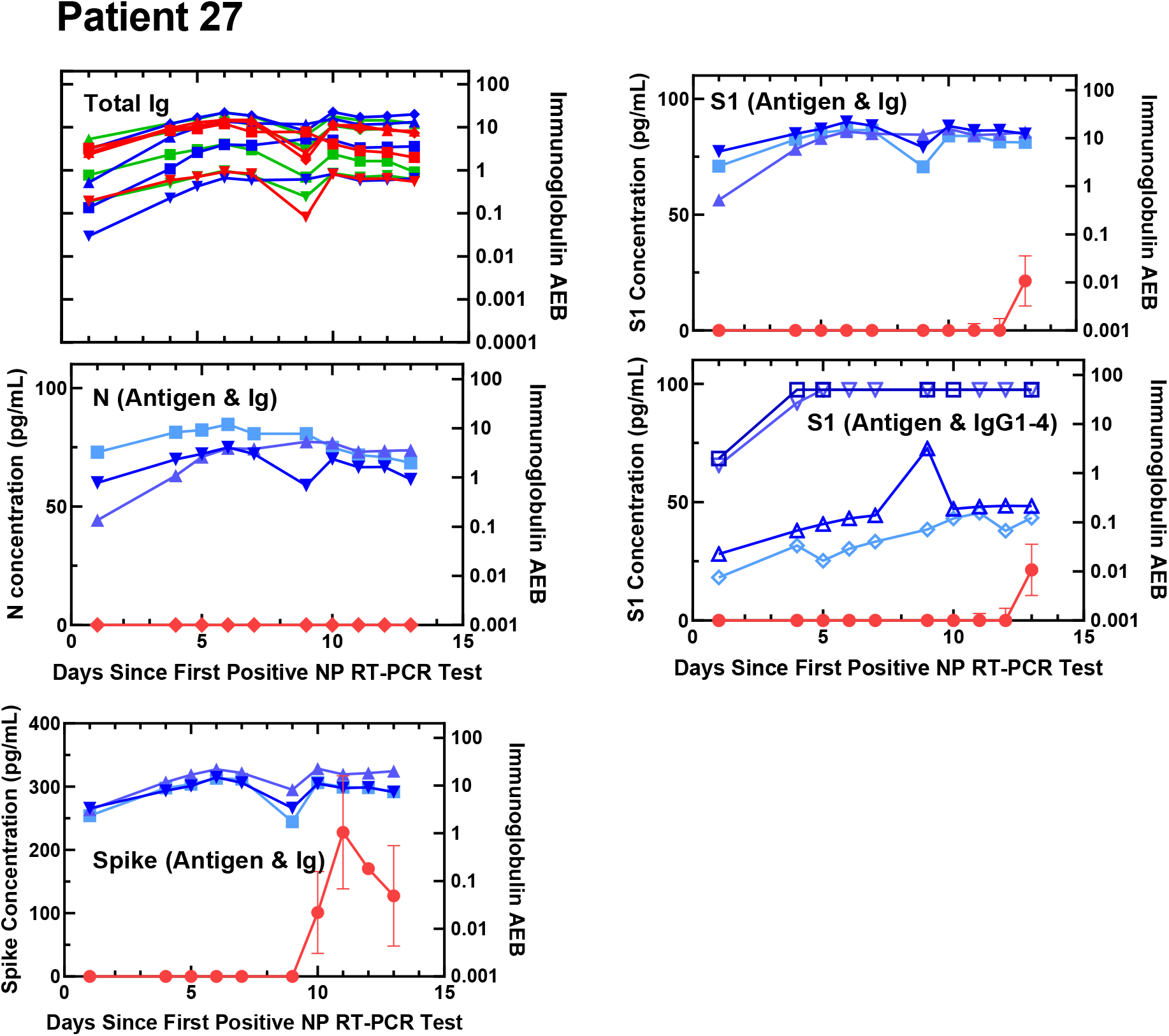
Viral antigen and anti-SARS-CoV-2 immunoglobulin plots for Patient 27.

**Figure S34.**
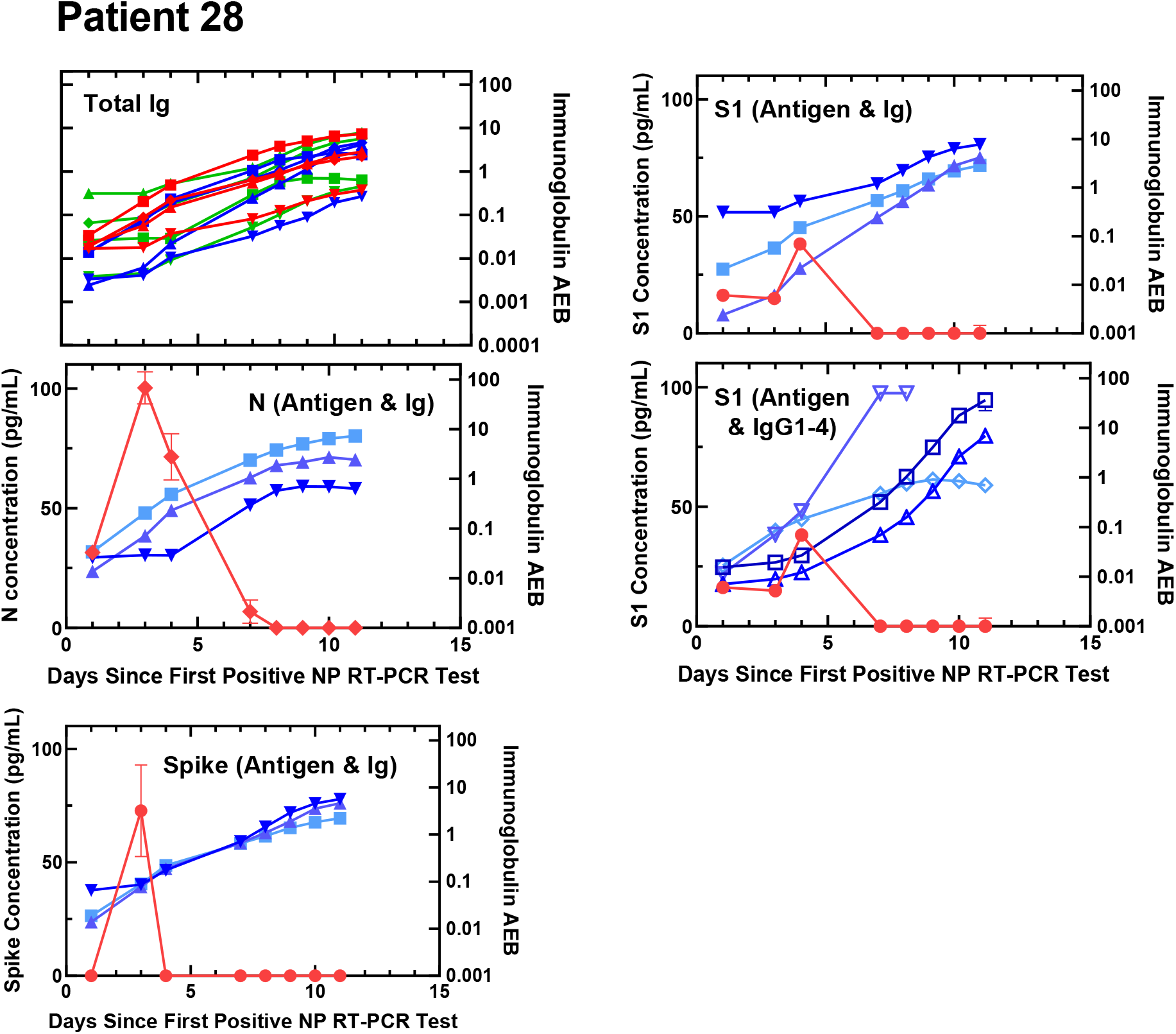
Viral antigen and anti-SARS-CoV-2 immunoglobulin plots for Patient 28.

**Figure S35.**
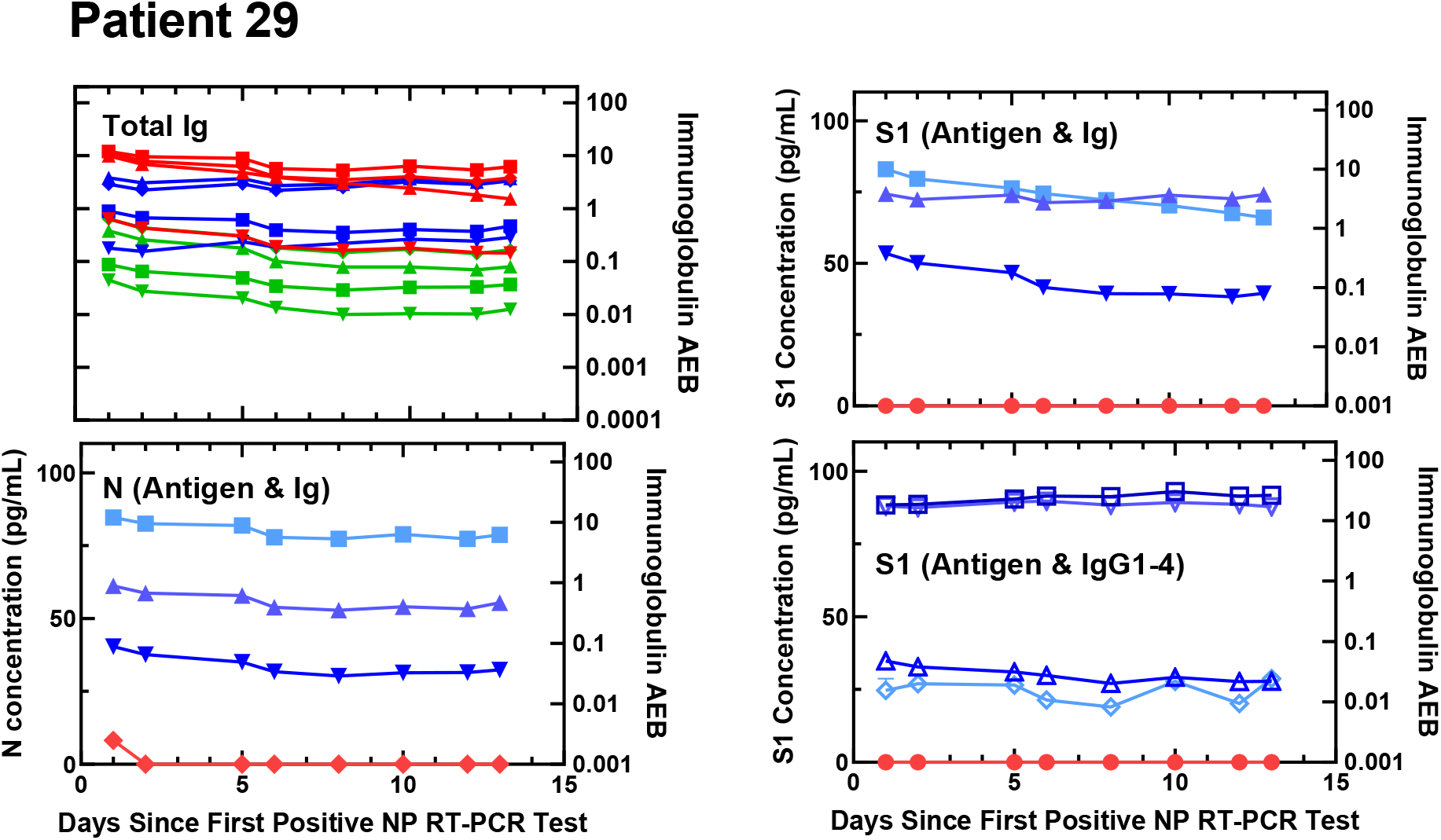
Viral antigen and anti-SARS-CoV-2 immunoglobulin plots for Patient 29.

**Figure S36.**
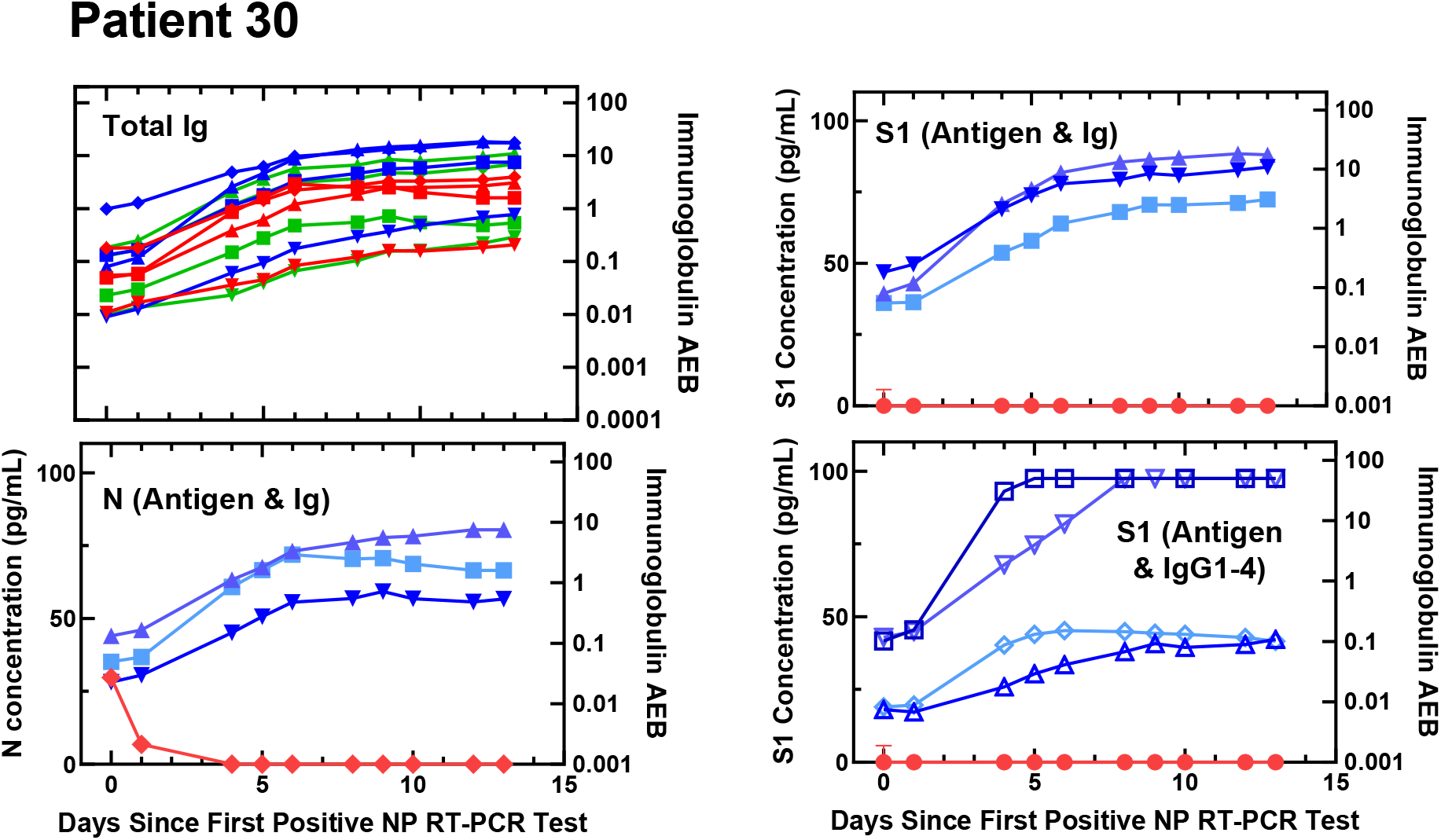
Viral antigen and anti-SARS-CoV-2 immunoglobulin plots for Patient 30.

**Figure S37.**
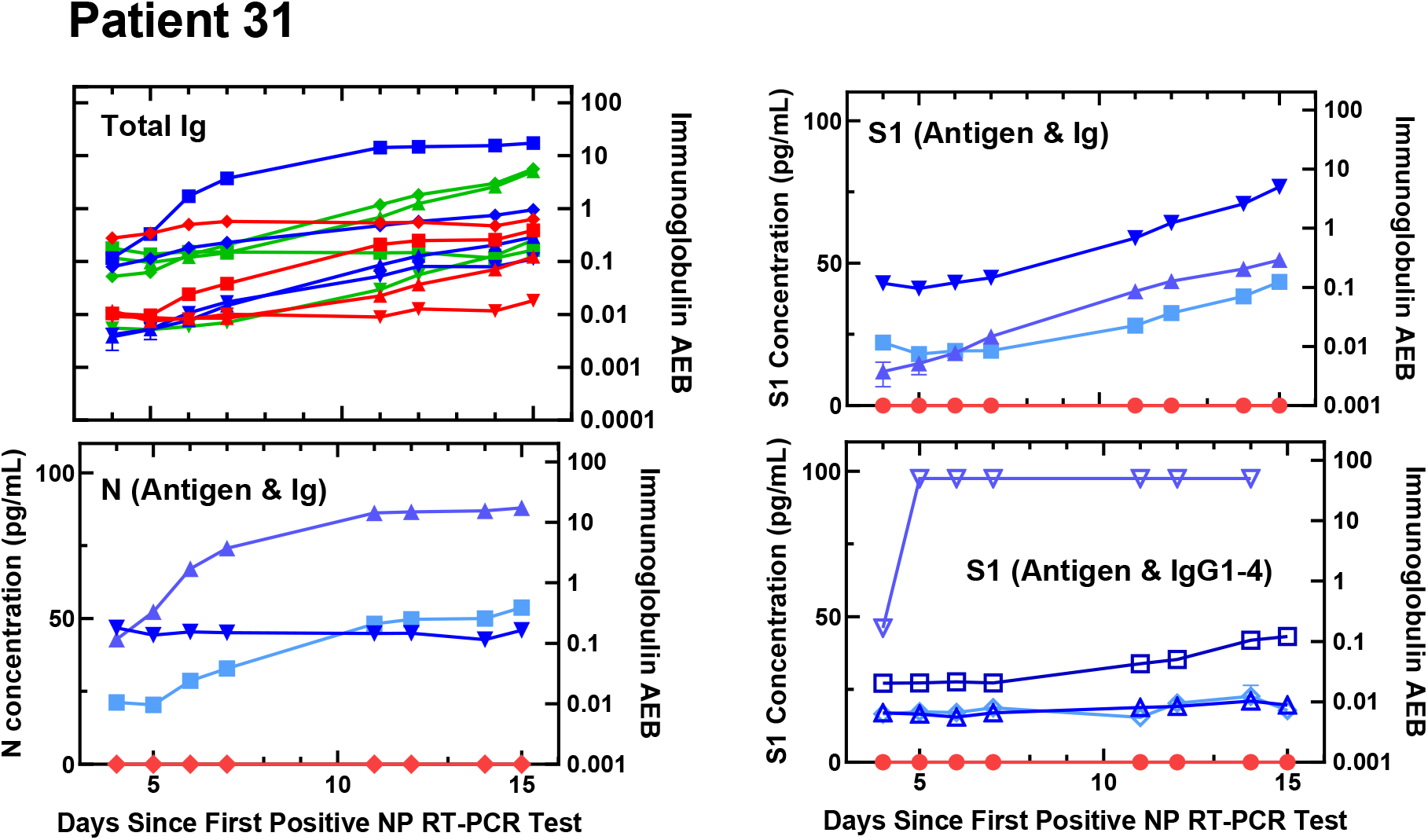
Viral antigen and anti-SARS-CoV-2 immunoglobulin plots for Patient 31.

**Figure S38.**
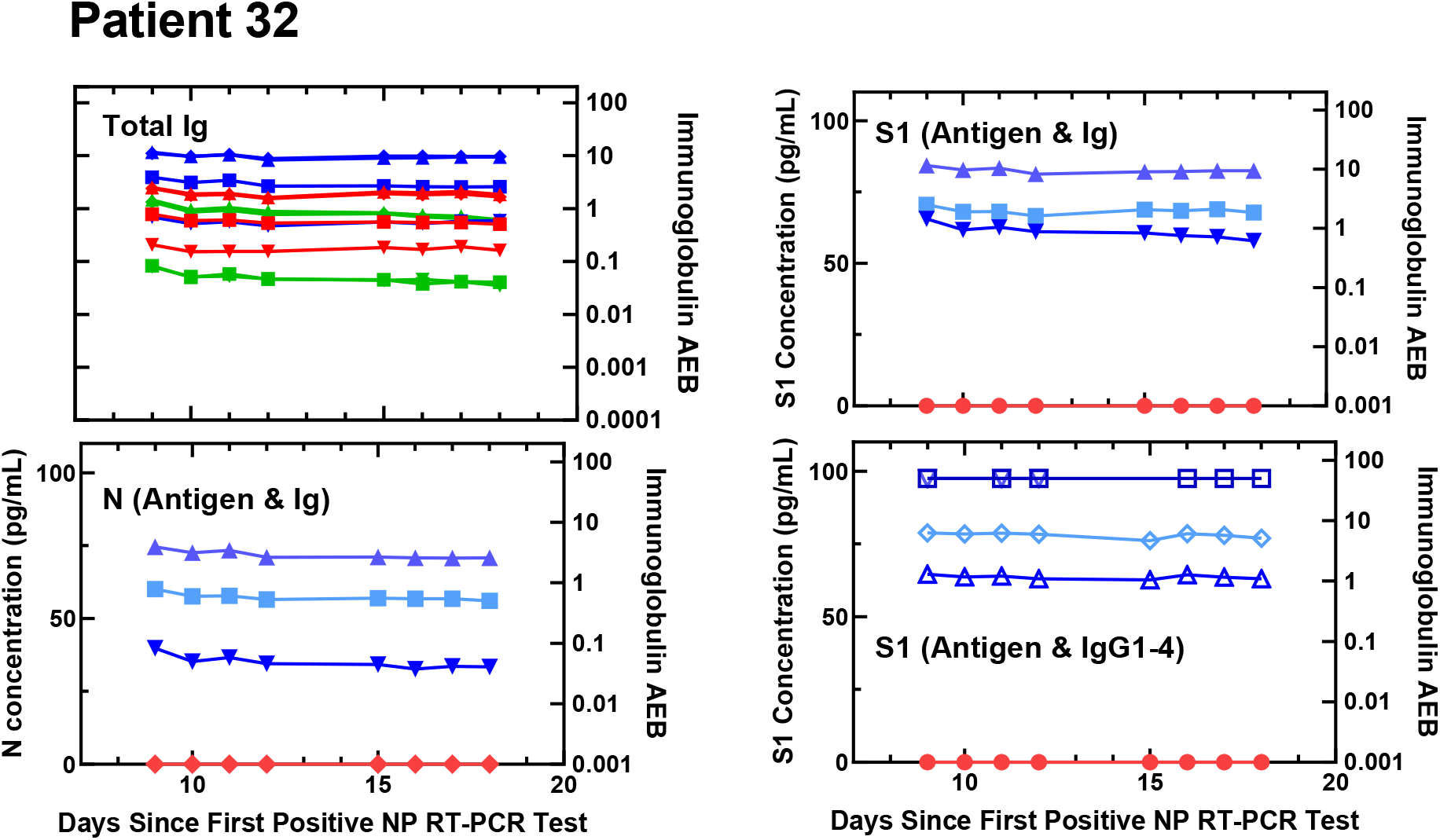
Viral antigen and anti-SARS-CoV-2 immunoglobulin plots for Patient 32.

**Figure S39.**
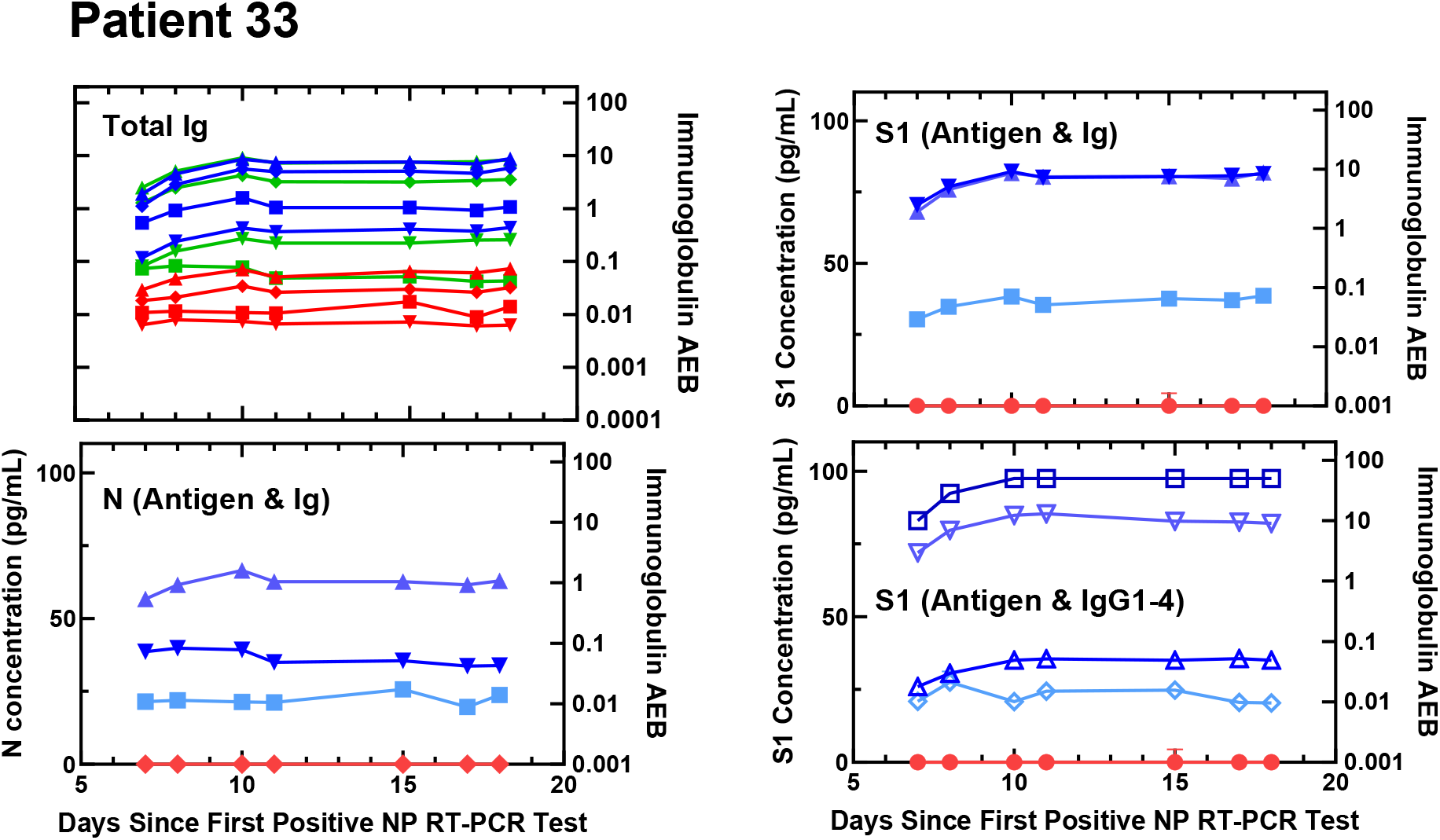
Viral antigen and anti-SARS-CoV-2 immunoglobulin plots for Patient 33.

**Figure S40.**
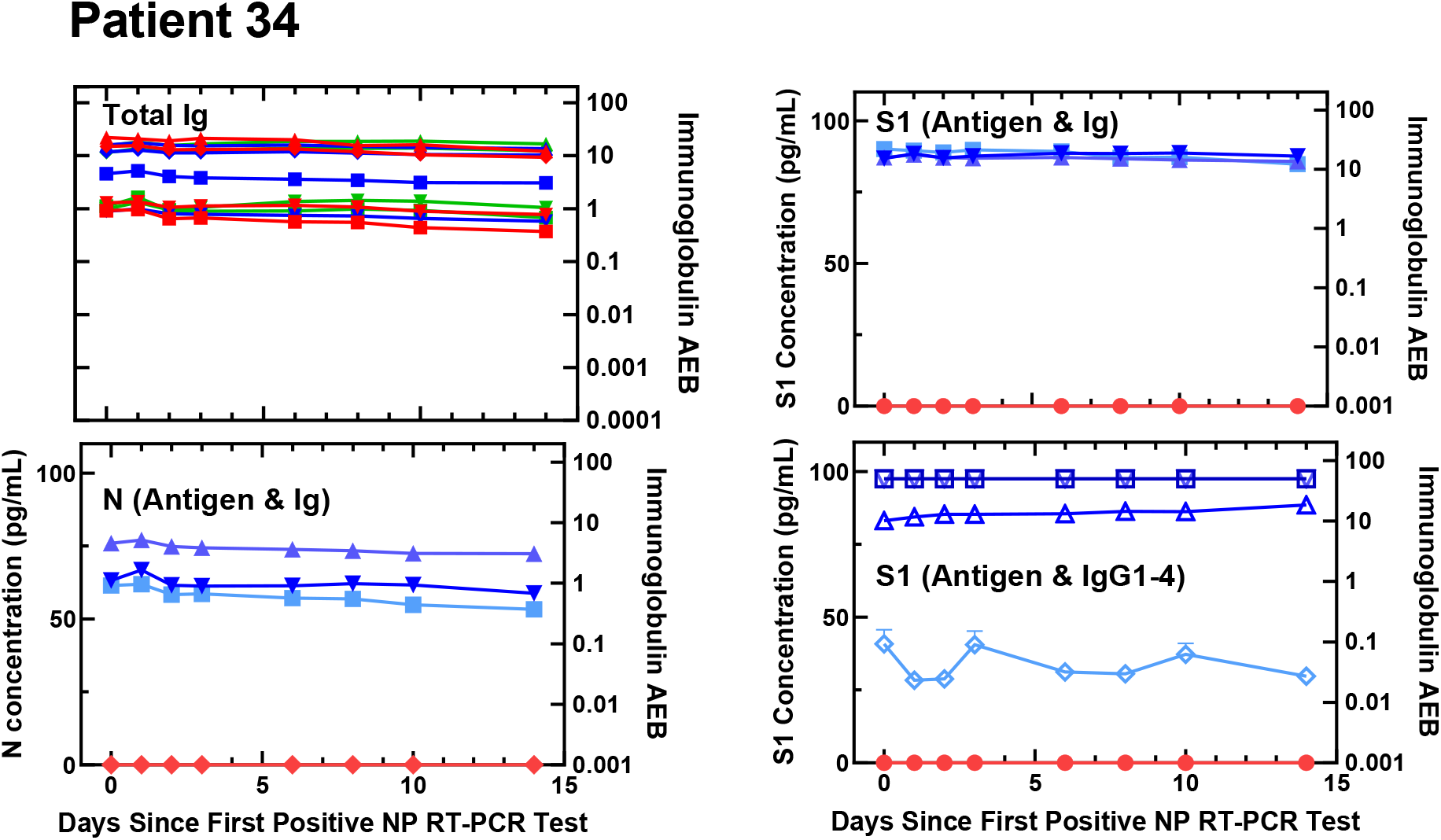
Viral antigen and anti-SARS-CoV-2 immunoglobulin plots for Patient 34.

**Figure S41.**
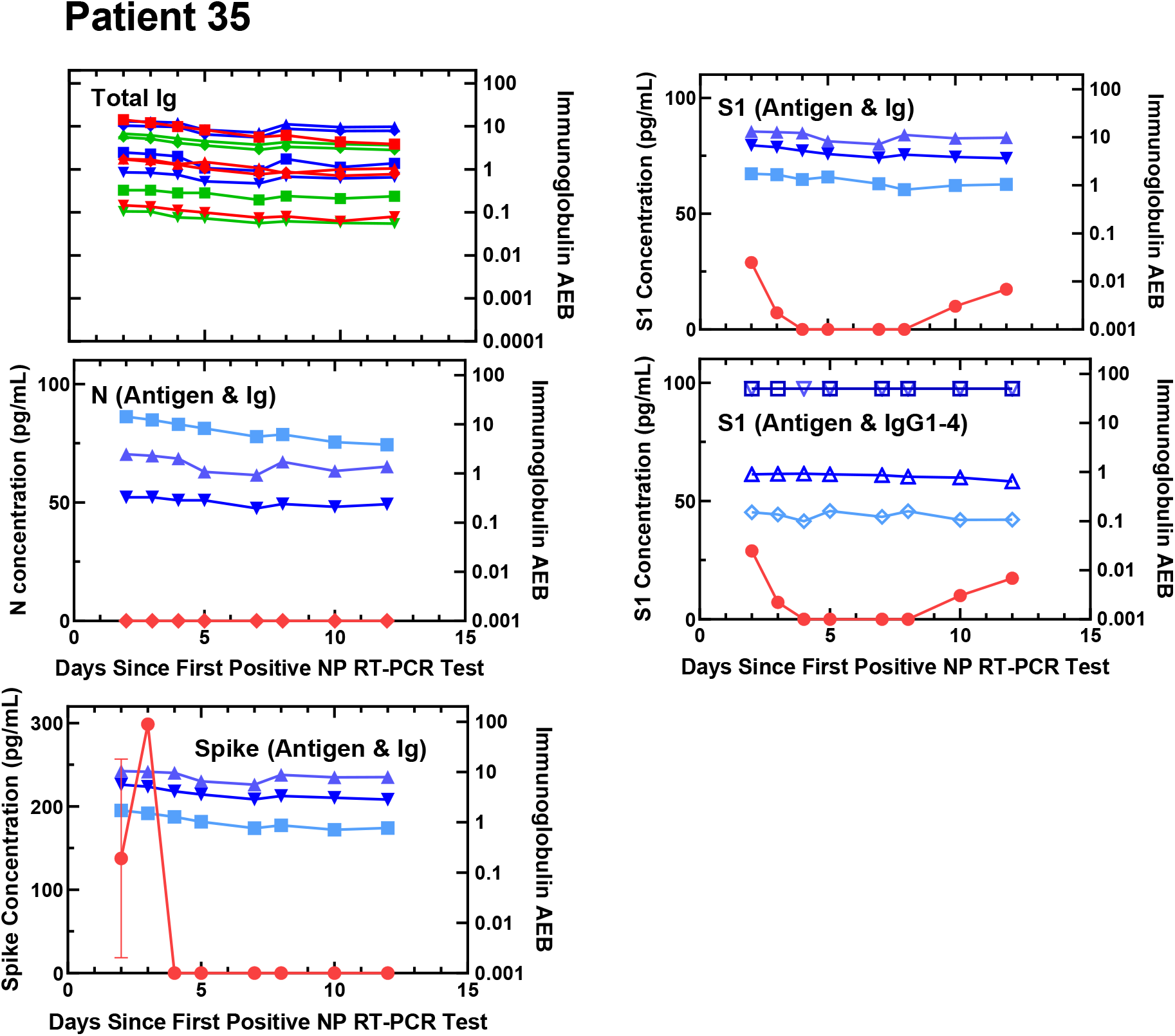
Viral antigen and anti-SARS-CoV-2 immunoglobulin plots for Patient 35.

**Figure S42.**
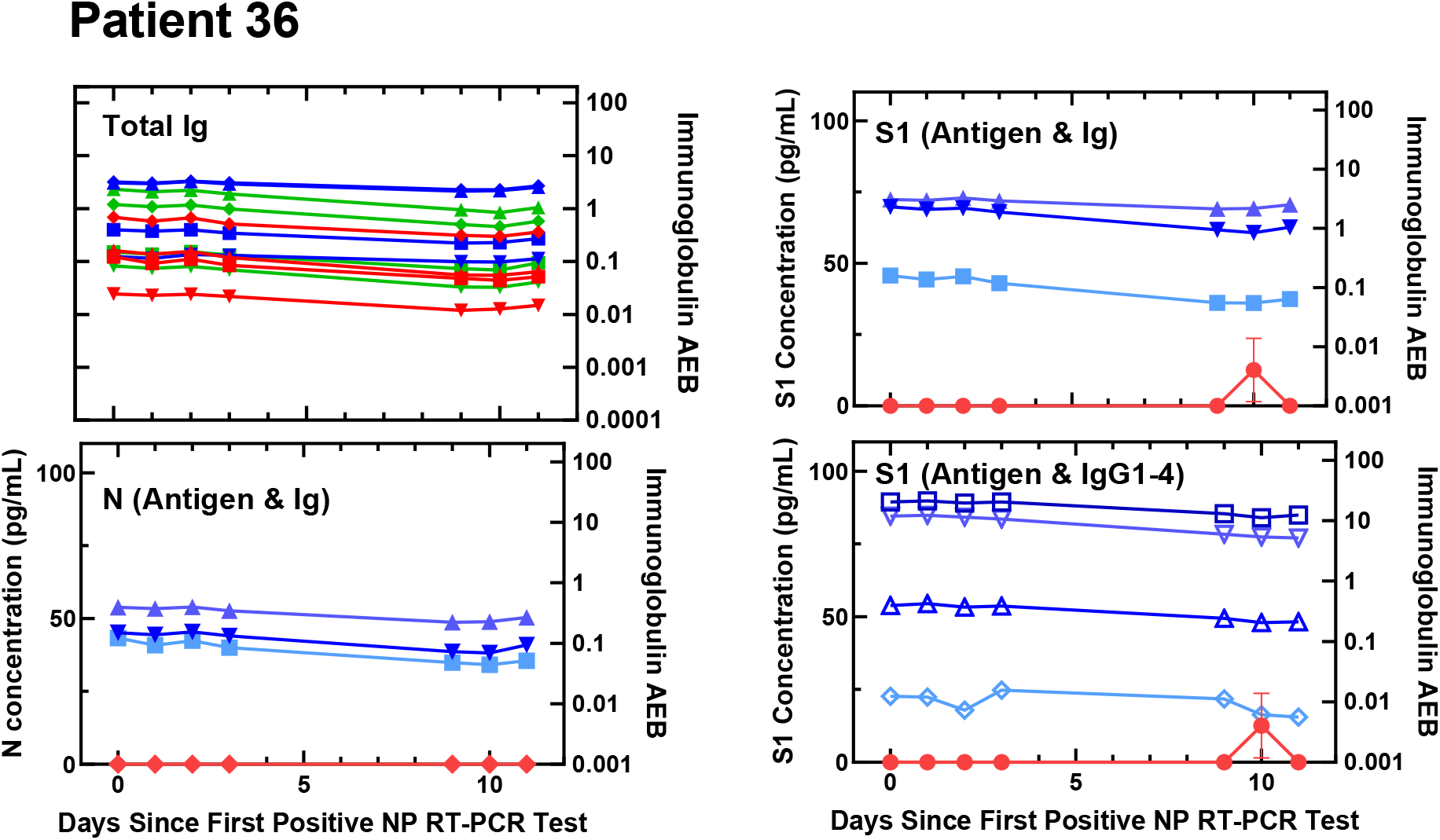
Viral antigen and anti-SARS-CoV-2 immunoglobulin plots for Patient 36.

**Figure S43.**
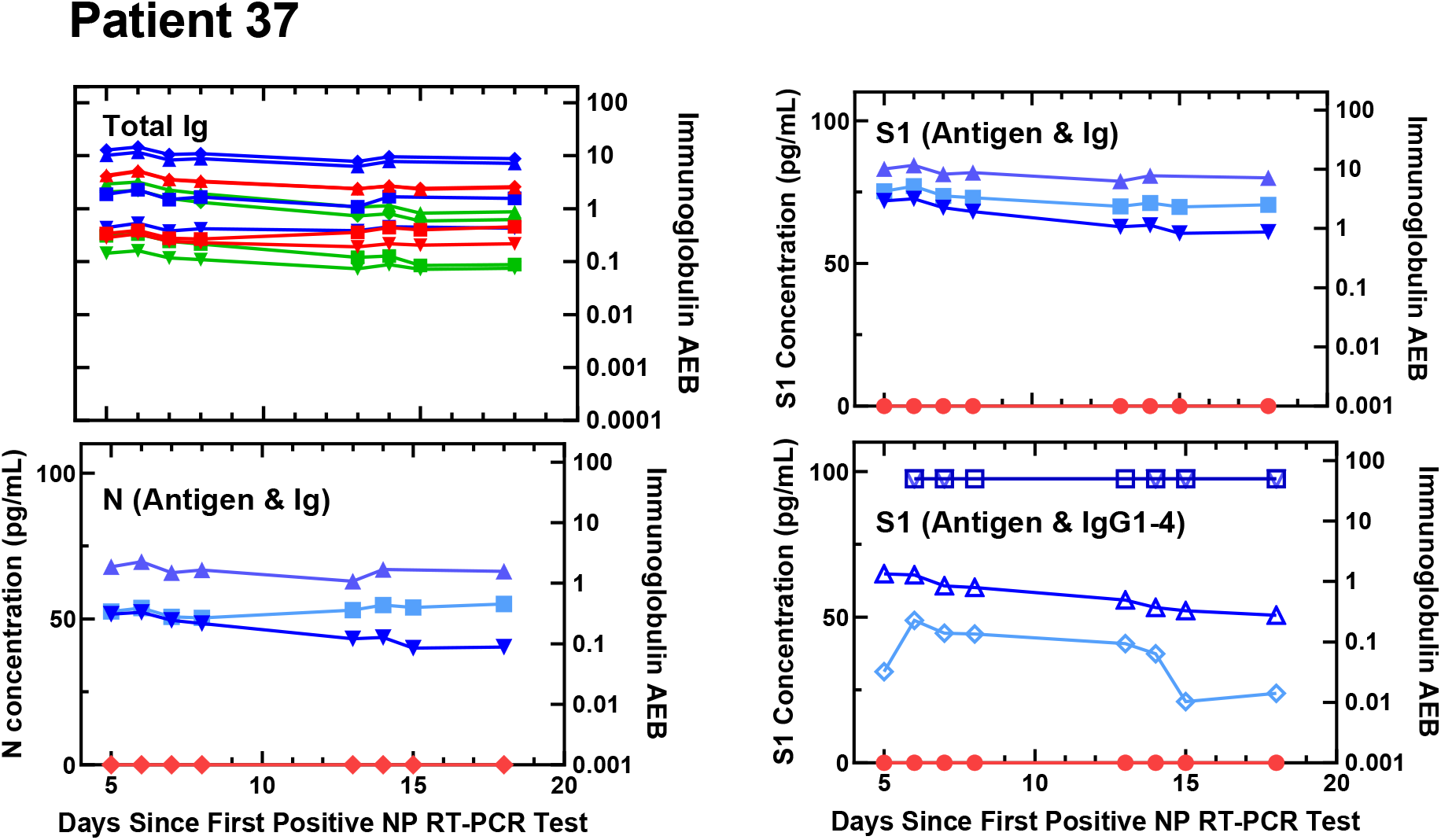
Viral antigen and anti-SARS-CoV-2 immunoglobulin plots for Patient 37.

**Figure S44.**
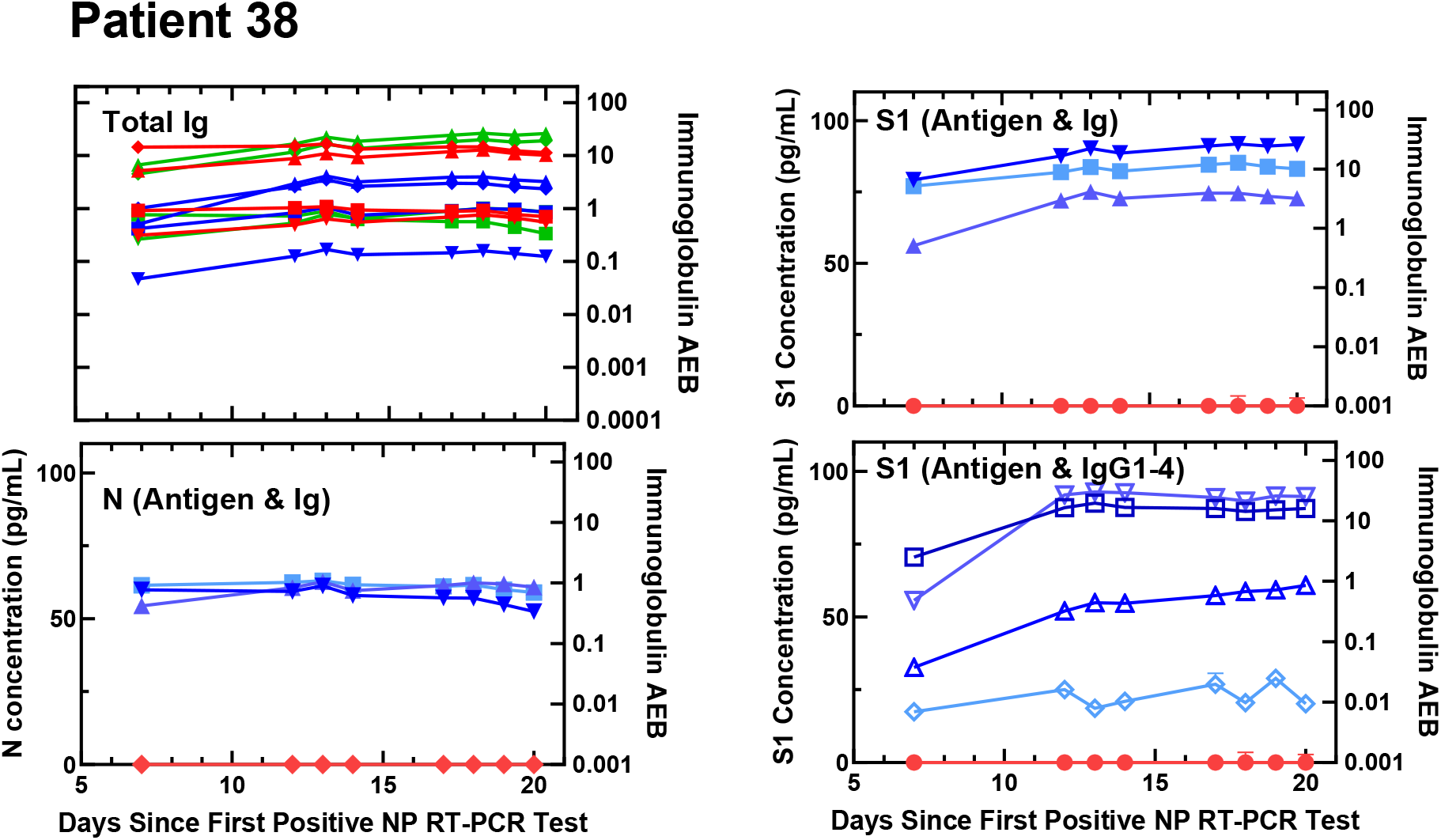
Viral antigen and anti-SARS-CoV-2 immunoglobulin plots for Patient 38.

**Table S4.** Individual patient clinical data.

| **Patient Number** | **Age** | **Gender** | **Patient disposition at time of admission** | **Was Patient Immunosuppressed?** |
| --- | --- | --- | --- | --- |
| 1 | 59 | Male | ICU | No |
| 2 | 40 | Male | ICU | No |
| 3 | 89 | Male | Floor admission | No |
| 4 | 60 | Female | Floor admission | No |
| 5 | 71 | Male | ICU | No |
| 6 | 75 | Male | ICU | No |
| 7 | 33 | Female | Floor admission | Yes |
| 8 | 49 | Male | ICU | No |
| 9 | 32 | Male | ICU | No |
| 10 | 54 | Female | ICU | No |
| 11 | 77 | Female | Floor admission then ICU later in hospitalization | No |
| 12 | 68 | Male | Floor admission then ICU later in hospitalization | No |
| 13 | 50 | Male | Floor admission | No |
| 14 | 64 | Male | Floor admission then ICU later in hospitalization | Yes |
| 15 | 78 | Male | ICU | No |
| 16 | 89 | Male | ICU | No |
| 17 | 50 | Male | ICU | No |
| 18 | 67 | Male | Floor admission | No |
| 19 | 81 | Female | Floor admission | No |
| 20 | 79 | Male | ICU | No |
| 21 | 67 | Female | ICU | No |
| 22 | 66 | Female | ICU | No |
| 23 | 70 | Male | ICU | No |
| 24 | 41 | Female | ICU | No |
| 25 | 54 | Female | ICU | No |
| 26 | 69 | Female | ICU | No |
| 27 | 53 | Male | ICU | No |
| 28 | 69 | Female | ICU | No |
| 29 | 63 | Female | ICU | No |
| 30 | 72 | Male | Floor admission then ICU later in hospitalization | No |
| 31 | 47 | Male | Floor admission then ICU later in hospitalization | Yes |
| 32 | 58 | Male | Floor admission then<br>ICU later in<br>hospitalization | No |
| 33 | 44 | Male | Floor admission | No |
| 34 | 45 | Male | Floor admission then<br>ICU later in<br>hospitalization | No |
| 35 | 78 | Male | Floor admission then<br>ICU later in<br>hospitalization | No |
| 36 | 37 | Female | Floor admission then<br>ICU later in<br>hospitalization | No |
| 37 | 58 | Male | Floor admission then<br>ICU later in<br>hospitalization | No |
| 38 | 43 | Male | Floor admission then<br>ICU later in<br>hospitalization | No |
| 39 | 56 | Female | Floor admission then<br>ICU later in<br>hospitalization | No |

### IV. Statistical Analysis

Statistical analysis was performed in GraphPad Prism version 8.3.0.

**Table S5.** Statistics for S1 as predictors of clinical outcomes (3 groups). Patients were split into three groups based on S1 levels: (1) undetectable antigen levels (below the limit of detection/LOD), (2) low levels of antigen (6-50 pg/mL), and (3) high levels of antigen (>50 pg/mL).

| Clinical Outcome | Test | P value |
| --- | --- | --- |
| Fraction of patients admitted to ICU | Chi-square test | 0.0107 (significant) |
| Fraction of patients intubated | Chi-square test | 0.2752 (not significant) |
| Days between hospitalization and intubation | ANOVA (Kruskal-Wallis test and Dunn's multiple comparisons test) | **Difference among means** (0.0063, significant)<br>**Multiple comparisons:**<br><LOD vs. 6-50 pg/mL (0.7495, not significant)<br><LOD vs. >50 pg/mL (0.0050, significant)<br>6-50 pg/mL vs. >50 pg/mL (0.1225, not significant) |
| Length of intubation | ANOVA (Kruskal-Wallis test and Dunn's multiple comparisons test) | **Difference among means** (0.2297, not significant)<br>**Multiple comparisons:**<br><LOD vs. 6-50 pg/mL (>0.9999, not significant)<br><LOD vs. >50 pg/mL (0.3426, not significant)<br>6-50 pg/mL vs. >50 pg/mL (0.5482, not significant) |

To confirm that splitting the patients into three groups based on S1 levels in plasma did not affect the results of analysis of clinical outcomes (compared to splitting patients into two groups: undetectable S1 levels and detectable S1 levels), we repeated the statistical analysis using two patient groups. The results are shown below in Table S5. When split into two groups, the differences in fraction of patients admitted to the ICU and mean days between hospitalization and intubation are still significant.

**Table S6.** Statistics for S1 as predictors of clinical outcomes (2 groups). Patients were split into two groups based on S1 levels: (1) undetectable N levels (below the limit of detection/LOD) and (2) detectable N levels (above the LOD).

| Clinical Outcome | Test | P value |
| --- | --- | --- |
| Fraction of patients admitted to ICU | Fisher's exact test | 0.0184 (significant) |
| Fraction of patients intubated | Fisher's exact test | 0.4011 (not significant) |
| Days between hospitalization and intubation | Mann-Whitney test | 0.0138 (significant) |
| Length of intubation | Mann-Whitney test | 0.2871 (not significant) |

In addition to S1, we assessed if nucleocapsid levels in plasma are indicators of disease severity in COVID-19. The results are plotted below in Figure S43 and the results of the statistical analysis are shown in Table S6. For nucleocapsid, the difference in fraction of patients admitted to the ICU upon presentation is significantly different between the two groups (Figure S43A). Differences of all other clinical markers of severity are not statistically significant.

**Figure S45.**
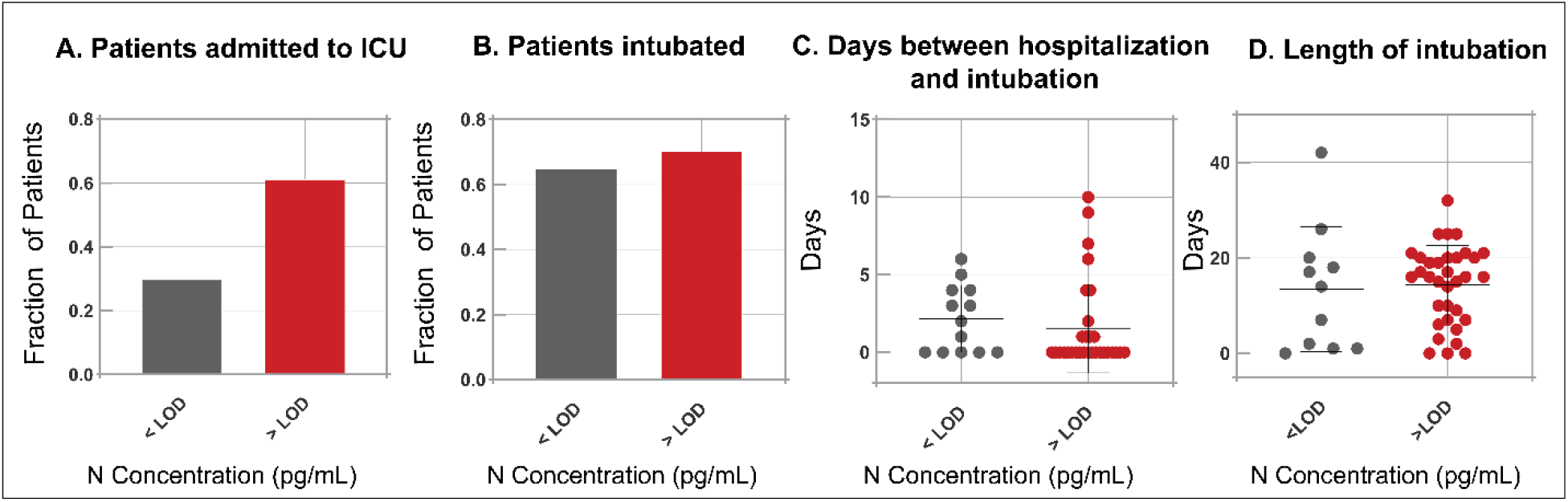
Clinical data for all COVID-19 positive patients plotted based on nucleocapsid levels in plasma.

**Table S7.** Statistics for Nucleocapsid as predictors of clinical outcomes (2 groups). Patients were split into two groups based on nucleocapsid levels: (1) undetectable antigen levels (below the limit of detection/LOD) and (2) detectable antigen levels (above the LOD).

| Clinical Outcome | Test | P value |
| --- | --- | --- |
| Fraction of patients admitted to ICU | Fisher's exact test | 0.0305 (significant) |
| Fraction of patients intubated | Fisher's exact test | 0.7729 (not significant) |
| Days between hospitalization and intubation | Mann-Whitney test | 0.1719 (not significant) |
| Length of intubation | Mann-Whitney test | 0.5610 (not significant) |

**Table S8.** Statistics for death rates.

| Antigen | Test | P value |
| --- | --- | --- |
| S1 (3 groups: <LOD, 6-50 pg/mL, and >50 pg/mL) | Chi-square test | 0.1921 (not significant) |
| S1 (2 groups: <LOD and >LOD) | Fisher's exact test | 0.5636 (not significant) |
| N (2 groups: <LOD and >LOD) | Fisher's exact test | 0.5148 (not significant) |

### V. Saliva Data

**Figure S46.**
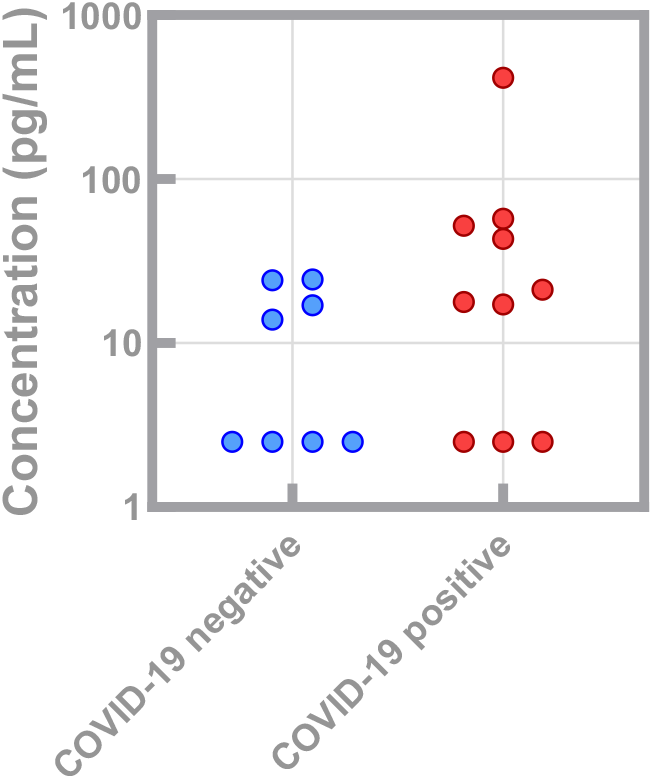
S1 concentrations in saliva. Saliva samples were collected from patients presenting to the Emergency Department at Brigham and Women’s Hospital (Boston, MA, USA). Patients were determined COVID-19 negative or positive by NP RT-PCR.

**Figure S47.**
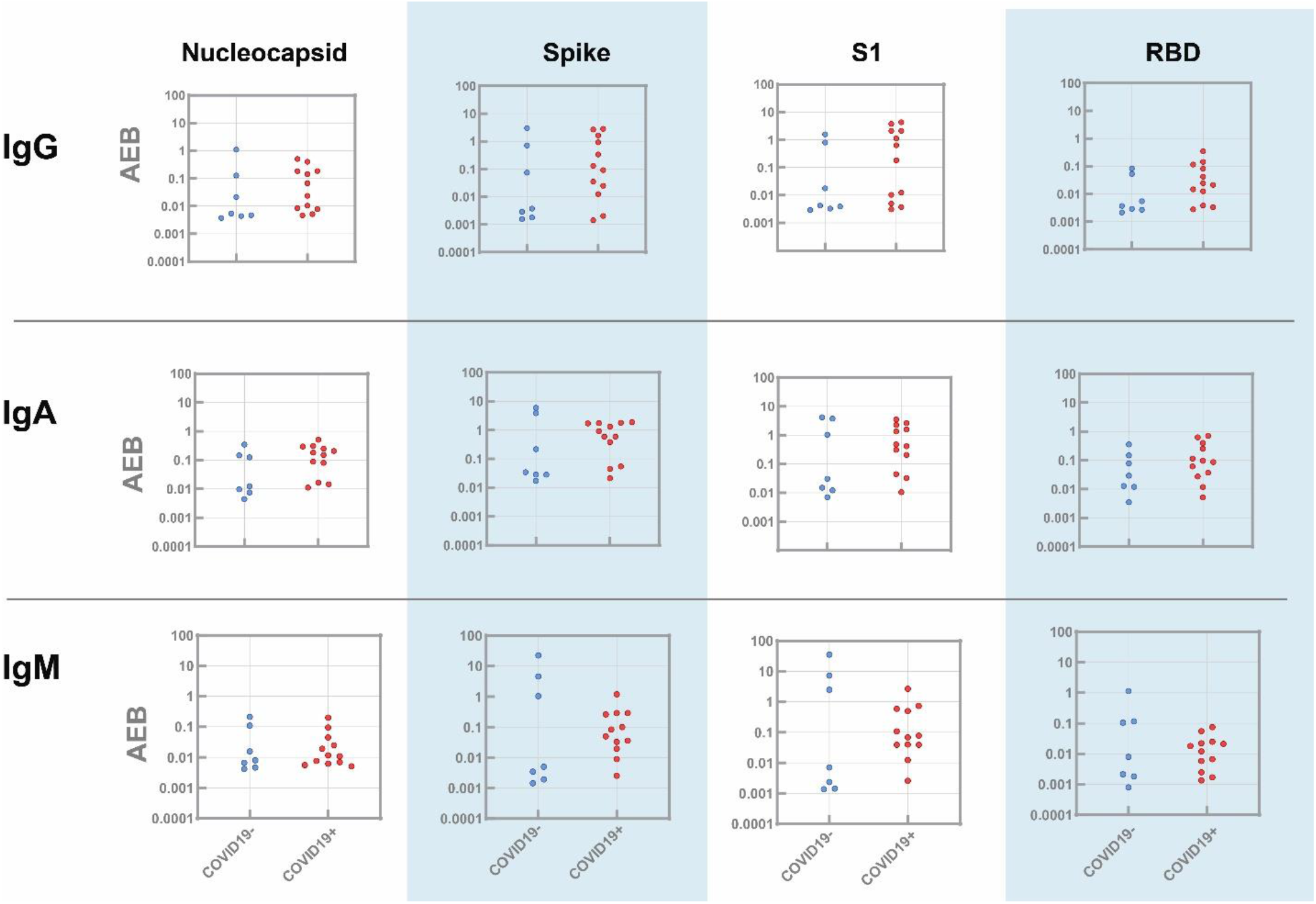
Simoa results for IgG, IgM, and IgG against four viral antigens in saliva. Each marker is the average AEB (average enzyme per bead) value from two replicate measurements.

**Figure S48.**
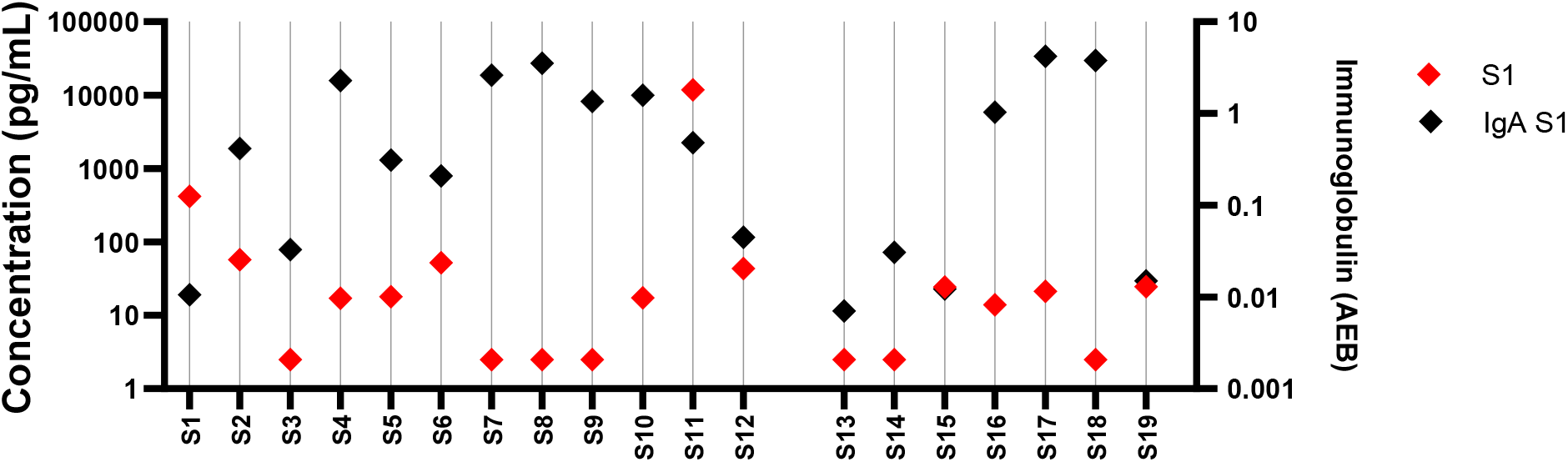
Correlated S1 and IgA (against S1) levels in individual patient saliva samples. S1 levels measured below the LOD (5 pg/mL in saliva) are considered undetectable. IgA levels with AEB< 0.05 are considered background level readings. COVID-19 positive patients are plotted on the left side of the graph (S1-S12) and COVID-19 negative patients are plotted on the right side of the graph (S13-S19).

## References and Notes

1. World Health Organization, Coronavirus disease (COVID-19) Situation Report 178 (2020), (available at https://www.who.int/docs/default-source/coronaviruse/situation-reports/20200716-covid-19-sitrep-178.pdf?sfvrsn=28ee165b_2).

2. N. Sethuraman, S. S. Jeremiah, A. Ryo, Interpreting Diagnostic Tests for SARS-CoV-2. JAMA. 323, 2249–2251 (2020).

3. L. Rodriguez, P. Pekkarinen, L. K. Tadepally, Z. Tan, C. R. Consiglio, C. Pou, Y. Chen, C. H. Mugabo, A. N. Quoc, K. Nowlan, T. Strandin, L. Levanov, J. Mikes, J. Wang, A. Kantele, J. Hepojoki, O. Vapalahti, S. Heinonen, E. Kekalainen, P. Brodin, medRxiv, in press, doi:10.1101/2020.06.03.20121582.

4. L. Ni, F. Ye, M. L. Cheng, Y. Feng, Y. Q. Deng, H. Zhao, P. Wei, J. Ge, M. Gou, X. Li L. Sun, T. Cao, P. Wang, C. Zhou, R. Zhang, P. Liang, H. Guo, X. Wang, C. F. Qin, F. Chen, C. Dong, Detection of SARS-CoV-2-Specific Humoral and Cellular Immunity in COVID-19 Convalescent Individuals. Immunity. 52, 971–977.e3 (2020).

5. F. Amanat, D. Stadlbauer, S. Strohmeier, T. H. O. Nguyen, V. Chromikova, M. McMahon, K. Jiang, G. A. Arunkumar, D. Jurczyszak, J. Polanco, M. Bermudez-Gonzalez, G. Kleiner, T. Aydillo, L. Miorin, D. S. Fierer, L. A. Lugo, E. M. Kojic, J. Stoever, S. T. H. Liu, C. Cunningham-Rundles, P. L. Felgner, T. Moran, A. García-Sastre, D. Caplivski, A. C. Cheng, K. Kedzierska, O. Vapalahti, J. M. Hepojoki, V. Simon, F. Krammer, A serological assay to detect SARS-CoV-2 seroconversion in humans. Nat. Med. 26 (2020), doi:10.1038/s41591-020-0913-5.

6. S. Mahapatra, P. Chandra, Clinically practiced and commercially viable nanobio engineered analytical methods for COVID-19 diagnosis. Biosens. Bioelectron. 165, 112361 (2020).

7. Veritor ™ System Veritor ™ System (2020), (available at https://www.fda.gov/media/139755/download).

8. A. J. Rivnak, D. M. Rissin, C. W. Kan, L. Song, M. W. Fishburn, T. Piech, T. G. Campbell, D. R. DuPont, M. Gardel, S. Sullivan, B. A. Pink, C. G. Cabrera, D. R. Fournier, D. C. Duffy, A fully-automated, six-plex single molecule immunoassay for measuring cytokines in blood. J. Immunol. Methods. 424, 20–27 (2015).

9. M. Norman, T. Gilboa, A. F. Ogata, A. M. Maley, L. Cohen, Y. Cai, J. Zhang, J. E. Feldman, B. M. Hauser, T. M. Caradonna, B. Chen, A. G. Schmidt, G. Alter, R. C. Charles, E. T. Ryan, D. R. Walt, Ultra-Sensitive High-Resolution Profiling of Anti-SARS- CoV-2 Antibodies for Detecting Early Seroconversion in COVID-19 Patients. medRxiv (2020), doi:10.1101/2020.04.28.20083691.

10. K. Steven, Woloshin, Neeraj Patel, Aaron S, False Negative Tests for SARS-CoV-2 Infection — Challenges and Implications. N. Engl. J. Med., 1–2 (2020).

11. Q. X. Long, B. Z. Liu, H. J. Deng, G. C. Wu, K. Deng, Y. K. Chen, P. Liao, J. F. Qiu, Y. Lin, X. F. Cai, D. Q. Wang, Y. Hu, J. H. Ren, N. Tang, Y. Y. Xu, L. H. Yu, Z. Mo, F. Gong, X. L. Zhang, W. G. Tian, L. Hu, X. X. Zhang, J. L. Xiang, H. X. Du, H. W. Liu, C. H. Lang, X. H. Luo, S. B. Wu, X. P. Cui, Z. Zhou, M. M. Zhu, J. Wang, C. J. Xue, X. F. Li, L. Wang, Z. J. Li, K. Wang, C. C. Niu, Q. J. Yang, X. J. Tang, Y. Zhang, X. M. Liu, J. J. Li, D. C. Zhang, F. Zhang, P. Liu, J. Yuan, Q. Li, J. L. Hu, J. Chen, A. L. Huang, Antibody responses to SARS-CoV-2 in patients with COVID-19. Nat. Med. 26 (2020), doi:10.1038/s41591-020-0897-1.

12. F. Amanat, T. H. O. Nguyen, V. Chromikova, S. Strohmeier, D. Stadlbauer, A serological assay to detect SARS-CoV-2 seroconversion in humans (2020).

13. A. T. Huang, B. Garcia-Carreras, M. D. T. Hitchings, B. Yang, L. Katzelnick, S. M. Rattigan, B. Borgert, C. Moreno, B. D. Solomon, I. Rodriguez-Barraquer, J. Lessler, H. Salje, D. S. Burke, A. Wesolowski, D. A. T. Cummings, medRxiv, in press, doi:10.1101/2020.04.14.20065771.

14. D. F. Robbiani, C. Gaebler, F. Muecksch, J. C. C. Lorenzi, Z. Wang, A. Cho, M. Agudelo, C. O. Barnes, S. Finkin, T. Hagglof, T. Y. Oliveira, C. Viant, A. Hurley, K. G. Millard, R. G. Kost, M. Cipolla, A. Gazumyan, K. Gordon, F. Bianchini, S. T. Chen, V. Ramos, R. Patel, J. Dizon, I. Shimeliovich, P. Mendoza, H. Hartweger, L. Nogueira, M. Pack, J. Horowitz, F. Schmidt, Y. Weisblum, H.-H. Hoffmann, E. Michailidis, A. W. Ashbrook, E. Waltari, J. E. Pak, K. E. Huey-Tubman, N. Koranda, P. R. Hoffman, A. P. West, C. M. Rice, T. Hatziioannou, P. J. Bjorkman, P. D. Bieniasz, M. Caskey, M. C. Nussenzweig, bioRxiv, in press, doi:10.1101/2020.05.13.092619.

15. G. Vidarsson, G. Dekkers, T. Rispens, IgG subclasses and allotypes: From structure to effector functions. Front. Immunol. 5, 1–17 (2014).

16. L. P. Daley, M. A. Kutzler, B. W. Bennett, M. C. Smith, A. L. Glaser, J. A. Appleton, Effector functions of camelid heavy-chain antibodies in immunity to West Nile virus. Clin. Vaccine Immunol. 17, 239–246 (2010).

17. P. Koraka, C. Suharti, T. E. Setiati, A. T. A. Mairuhu, E. Van Gorp, C. E. Hach, M. Juffrie, J. Sutaryo, G. M. Van Der Meer, J. Groen, A. D. M. E. Osterhaus, Kinetics of dengue virus-specific serum immunoglobulin classes and subclasses correlate with clinical outcome of infection. J. Clin. Microbiol. 39, 4332–4338 (2001).

18. O. Scharf, H. Golding, L. R. King, N. Eller, D. Frazier, B. Golding, D. E. Scott, Immunoglobulin G3 from Polyclonal Human Immunodeficiency Virus (HIV) Immune Globulin Is More Potent than Other Subclasses in Neutralizing HIV Type 1. J. Virol. 75, 6558–6565 (2001).

19. L. I. Spinsanti, A. A. Farías, J. J. Aguilar, M. P. del Díaz, M. S. Contigiani, Immunoglobulin G subclasses in antibody responses to St. Louis encephalitis virus infections. Arch. Virol. 156, 1861–1864 (2011).

20. R. Wölfel, V. M. Corman, W. Guggemos, M. Seilmaier, S. Zange, M. A. Müller, D. Niemeyer, T. C. Jones, P. Vollmar, C. Rothe, M. Hoelscher, T. Bleicker, S. Brünink, J. Schneider, R. Ehmann, K. Zwirglmaier, C. Drosten, C. Wendtner, Virological assessment of hospitalized patients with COVID-2019. Nature. 581, 465–469 (2020).

21. W. Wang, Y. Xu, R. Gao, R. Lu, K. Han, G. Wu, W. Tan, Detection of SARS-CoV-2 in Different Types of Clinical Specimens. Jama, 3–4 (2020).

22. C. Magro, J. J. Mulvey, D. Berlin, G. Nuovo, S. Salvatore, J. Harp, A. Baxter-Stoltzfus, J. Laurence, Complement associated microvascular injury and thrombosis in the pathogenesis of severe COVID-19 infection: a report of five cases. Transl. Res. 220, 1–13 (2020).

23. H. Amirfakhryan, F. safari, Outbreak of SARS-CoV2: Pathogenesis of infection and cardiovascular involvement. Hell. J. Cardiol. (2020), doi:10.1016/j.hjc.2020.05.007.

24. R. Channappanavar, A. R. Fehr, R. Vijay, M. Mack, J. Zhao, D. K. Meyerholz, S. Perlman, Dysregulated Type I Interferon and Inflammatory Monocyte-Macrophage Responses Cause Lethal Pneumonia in SARS-CoV-Infected Mice. Cell Host Microbe. 19, 181–193 (2016).

25. L. A. Teuwen, V. Geldhof, A. Pasut, P. Carmeliet, COVID-19: the vasculature unleashed. Nat. Rev. Immunol., 19–21 (2020).

## References

24. A. J. Rivnak, D. M. Rissin, C. W. Kan, L. Song, M. W. Fishburn, T. Piech, T. G. Campbell, D. R. DuPont, M. Gardel, S. Sullivan, B. A. Pink, C. G. Cabrera, D. R. Fournier, D. C. Duffy, A y-automated, six-plex single molecule immunoassay for measuring cytokines in blood. J. Immunol. Methods. 424, 20–27 (2015).

